# Psychiatric comorbidity in dissociative identity disorder: A meta-analysis

**DOI:** 10.64898/2026.10.06.26364737

**Authors:** Emily Mitchell, Anisha Kazi, Andrew Watson, Susannah Pick, Devin B. Terhune

## Abstract

**Background:** Dissociative identity disorder (DID) is associated with frequent psychiatric comorbidity. Its complex and overlapping symptom profile can complicate recognition of both DID and co-occurring psychiatric conditions, contributing to diagnostic uncertainty. Evidence regarding psychiatric comorbidity is limited and prevalence estimates vary across studies.

**Aims:** This meta-analysis aimed to examine the prevalence of psychiatric comorbidities in DID and factors associated with variation in prevalence estimates.

**Methods:** Eligible studies were identified through PubMed, Scopus, Embase, PsycINFO, and MEDLINE from 1980 to 25 April 2026 (PROSPERO, no. CRD420261335819). Studies reporting psychiatric comorbidity in adults with DID were independently screened by two reviewers. Risk of bias was assessed. Random-effects meta-analyses using logit-transformed prevalence estimates were conducted. Meta-regressions examined moderators, whilst influence and sensitivity analyses assessed robustness. Certainty was evaluated using the GRADE approach.

**Results:** Twenty-four studies (*N* = 858) contributed to the meta-analysis across 23 psychiatric comorbidities. Among individuals with DID, posttraumatic stress disorder had the highest pooled prevalence (87.44%, 95% CI 71.79–94.96), followed by major depressive disorder (62.64%, 95% CI 33.45–84.84), and borderline personality disorder (47.05%, 95% CI 34.21–60.32). Substantial heterogeneity was noted across most estimates. Meta-regressions identified significant associations with participant characteristics, recruitment, study quality, and diagnostic method. Risk of bias and certainty of evidence were heterogenous.

**Conclusions:** Psychiatric comorbidity in DID is substantial and spans multiple diagnostic categories, although considerable heterogeneity and methodological variation limit confidence in prevalence estimates. Standardised diagnostic assessment, more representative sampling, and longitudinal research are needed to clarify the extent and clinical significance of comorbidity in DID.

---

Dissociative identity disorder (DID) is a form of pathological dissociation characterised by pronounced disruptions in the experience of selfhood and presentation of two or more quasi-distinct personality states. Epidemiological studies estimate the prevalence of DID to be 1-1.5% in the general population (1–3), with prevalence increasing to approximately 5% in inpatient settings (4). Individuals with DID are often subject to restrictive levels of care, including compulsory inpatient treatment, placing considerable demand and subsequent cost upon healthcare systems (5).

Individuals with DID often present with complex and heterogeneous symptom profiles that complicate clinical assessment and treatment. The predominance of overlapping symptoms, such as auditory hallucinations, emotional dysregulation, and somatisation can obscure the clinical presentation of DID (6, 7). On average, individuals with DID receive three to four misdiagnoses and spend between five and twelve years in healthcare services before receiving a diagnosis of DID (5). Consequently, individuals present to treatment services multiple times, receive misdirected and often ineffective, costly treatment, and are subject to increased risk and worsening symptoms (8, 9).

The clinical picture of DID is also marked by its frequent co-occurrence with other psychiatric disorders. A nationwide observational study of over two thousand individuals with DID in Germany reported 86.4% had at least one psychiatric comorbidity, and 23.9% had five or more (10). Such high rates of comorbidity are frequently observed in trauma-related populations, with previous literature suggesting that people with DID typically present with multiple non-dissociative psychiatric comorbidities (11). Despite this, there is little evidence directly evaluating the impact of psychiatric comorbidity on prognosis in DID. However, across psychiatric populations, including trauma-related disorders, comorbidity is strongly related to severity (12).

There is limited evidence regarding psychiatric comorbidity in DID. Many major epidemiological studies of DID do not systematically assess for comorbidity, and where comorbidities are reported, prevalence estimates vary across studies or are restricted to particular comorbid conditions. To date, no known study has systematically synthesised prevalence rates of individual psychiatric comorbidities in DID. Given the severity and marked functional impairment associated with DID, a greater understanding of its comorbidity profile may help to clarify factors that influence prognosis and risk and add to understanding of the relationship between DID and broader psychiatric symptomatology. In turn, this may further inform treatment planning which, when appropriate and directed, shows improvements in risk, global functioning, comorbid psychopathology, and cost to mental health systems (13–15). The primary aim of this meta-analysis was to quantitatively synthesise prevalence rates of comorbid psychiatric disorders in adults with DID. The secondary aim was to examine study-and sample-level factors as potential sources of heterogeneity in prevalence estimates.

## Methods

The present meta-analysis was conducted in accordance with the Preferred Reporting Items for Systematic Reviews and Meta-Analyses (PRISMA) guidelines (16). The PRISMA checklist is available in Supplementary Table 1. A protocol for this study was established *a priori* and pre-registered on PROSPERO (CRD420261335819).

### Search Strategy and Selection Criteria

A systematic search of the literature was conducted in PubMed (via National Library of Medicine), Scopus (via Elsevier), Embase, PsycINFO, and MEDLINE (via Ovid) on 13/10/2025, subsequently updated on 25/04/2026. Searches were restricted to articles published from 1980 onwards, the year the Diagnostic and Statistical Manual of Mental Disorders, third edition (DSM-III) (17), was published, formally recognising DID (then, multiple personality disorder) as a psychiatric diagnosis. The search strategy incorporated current and historical terms for all psychiatric disorders recognised in DSM and International Statistical Classification of Diseases and Related Health Problems (ICD) (18, 19) from DSM-III onwards. The full search strategy is presented in the supplementary materials. To ensure no eligible study was missed from the review, the search was supplemented by a manual search of the reference lists of included studies. References were imported into Covidence (20) review software.

Supplementary Table 2 presents the full selection criteria according to the PECOS framework (21). Two independent reviewers (EM, AK) screened all studies at every stage for eligibility. Where discrepancies arose between reviewers, a third reviewer (DBT) arbitrated. Authors were contacted where eligibility could not be established from available information. Studies that listed rates of psychiatric disorders as “previous diagnoses” were excluded due to ambiguity relating to whether psychiatric disorders represented true comorbidities or exemplified the pattern of misdiagnosis that is characteristic of individuals with DID (5). In the event of overlapping samples, the largest sample (or earliest paper if sample sizes were identical) was extracted.

### Data Extraction

Data were extracted independently and in parallel with reviewer blinding. Discrepancies were resolved through discussion with the third reviewer. The primary outcome was current reported prevalence estimates of psychiatric comorbidities in individuals with DID. Psychiatric comorbidities were harmonised according to DSM-5 classification to facilitate synthesis across studies. Where several historic diagnoses corresponded to a single DSM-5 diagnosis, the most frequently reported diagnosis across studies was extracted to minimise overlapping data. If a disorder was no longer recognised in the DSM-5, terminology from the most recent diagnostic classification system in which the disorder appeared was retained. Broad diagnostic categories combining several disorders (e.g., anxiety disorders), were not included.

Agreement between the two primary raters was assessed for both data extraction and risk-of-bias assessment prior to resolution of discrepancies by the third reviewer. Inter-rater percentage agreement was calculated for data extraction as outcomes were not binary and therefore Cohen’s κ was not appropriate. Percentage agreement across extracted variables was 81.09%. For assessment of risk of bias, agreement was assessed using a linearly weighted Cohen’s κ, reflecting substantial inter-rater agreement (κ = 0.65) (22).

Risk of bias was assessed by the same two reviewers using The Joanna Briggs Institute (JBI) Critical Appraisal Checklist for Studies Reporting Prevalence Data (23), with consultation regarding discrepancies with the third reviewer. To maintain relevance and consistency, the items of the checklist were operationalised according to their application to studies of individuals with DID. The nine checklist items were rated as “yes” (1), “no” (0), “unclear” (0) or “not applicable” (0). For descriptive purposes, a total score of 0-3 was considered to be low quality/high risk of bias, 4-6 to be moderate quality, and 7-9 to be high quality/low risk of bias.

### Data Analysis

Data were synthesised using a random-effects meta-analysis using the DerSimonian Laird estimator to estimate pooled prevalence and between-study variance (τ²) of psychiatric comorbidities among individuals with DID (24). Meta-analyses were conducted for individual psychiatric comorbidities reported by at least three independent studies. Logit transformation was conducted on prevalence estimates prior to analysis to stabilise variances and account for the bounded nature of proportion data. Pooled estimates and corresponding 95% confidence intervals (CIs) were subsequently back-transformed to the original proportion scale and presented as prevalence estimates (%). Meta-analyses were conducted using Jamovi (version 2.3.26.0) with the MAJOR module (version 1.2.4), with additional analyses conducted using MATLAB (version 2024a). The threshold for statistical significance was set to *p* < 0.05. Between-study heterogeneity was quantified using Cochran’s *Q*, *I*², and τ² statistics.

A forest plot was generated for each meta-analysis to show individual study logit-transformed prevalence estimates with 95% CIs, study weights, and the pooled estimate (see Supplementary Figures 1-23). Assessments of reporting bias were calculated using a funnel plot for each meta-analysis (see Supplementary Figures 24-46). Given the methodological limitations of funnel plots in prevalence meta-analyses (25), evidence of funnel plot asymmetry was reported descriptively and interpreted with caution.

To contextualise the severity of dissociative symptoms in participants with DID, an additional random-effects meta-analysis was conducted to estimate the pooled mean Experiences Scale (DES) (26) score and corresponding 95% CIs following Lyssenko, Schmahl (27).

Random-effects meta-regression analyses were conducted to identify potential sources of heterogeneity in prevalence rates. Analyses were undertaken only for psychiatric comorbidities with prevalence rates from at least 10 independent studies to improve reliability of regression estimates. Continuous moderators included publication year, mean age, percentage of female participants, and overall quality (JBI) score. Binary moderators included study setting, continent, recruitment source, DID diagnostic system, DID diagnostic method, and psychiatric comorbidity diagnostic method. Predicted differences in prevalence between categories were calculated from the fitted meta-regression models to aid interpretation.

Influence diagnostics were conducted to detect studies that exerted a disproportionate influence on pooled prevalence estimates. Diagnostics were conducted for meta-analyses with at least five independent studies (*k* ≥ 5). Possible outliers were identified using studentised residuals exceeding |3|. Potentially influential studies were identified using DFBETAS exceeding |1| or Cook’s distance exceeding 4/k. For each of these meta-analyses, leave-one-out analyses were also conducted by sequentially removing each study and recalculating the pooled prevalence, with the resulting range of prevalence estimates recorded. Where outliers or influential cases were identified, separate sensitivity analyses were conducted excluding the identified outlier(s) or influential study/studies, respectively, and the relevant meta-analyses were repeated.

Additional sensitivity analyses were conducted to assess the robustness of the pooled estimates to methodological restrictions. The potential influence of sample size, methodological quality, and diagnostic rigour on prevalence estimates were examined by individually restricting the analyses to studies with a sample size of ≥20, a JBI quality score of ≥7, and use of a validated, structured interview to diagnose DID or the psychiatric comorbidity. Sensitivity analyses were not conducted where the restriction resulted in fewer than three studies.

### Certainty of Evidence

The Grading of Recommendations, Assessment, Development and Evaluations (GRADE) (28) approach was used to determine the certainty of the body of evidence. In the absence of an established procedure to apply this to prevalence estimates, a framework developed for applying GRADE to prognostic studies was applied (29). In line with these recommendations, cross-sectional studies and baseline assessments from cohort studies were deemed the most appropriate evidence, so initially ranked as high certainty. Subsequently, certainty could be downgraded based upon concerns relating to risk of bias, inconsistency, indirectness, imprecision, and publication bias. Overall certainty for each prevalence estimate was classified as high, moderate, low, or very low.

## Results

### Search Results

As shown in Figure 1, 3,905 records were screened at the title and abstract stage, with 851 full-text articles screened for eligibility. Of these, 24 studies were identified for inclusion (see Supplementary Table 3). Prior to final inclusion, additional information was sought from four author groups relating to six individual studies; information sufficient to meet inclusion was provided for two studies. Two studies (30, 31) had overlapping samples with different psychiatric comorbidity outcomes, so were merged and treated as a single study at analysis.

**Figure 1.**
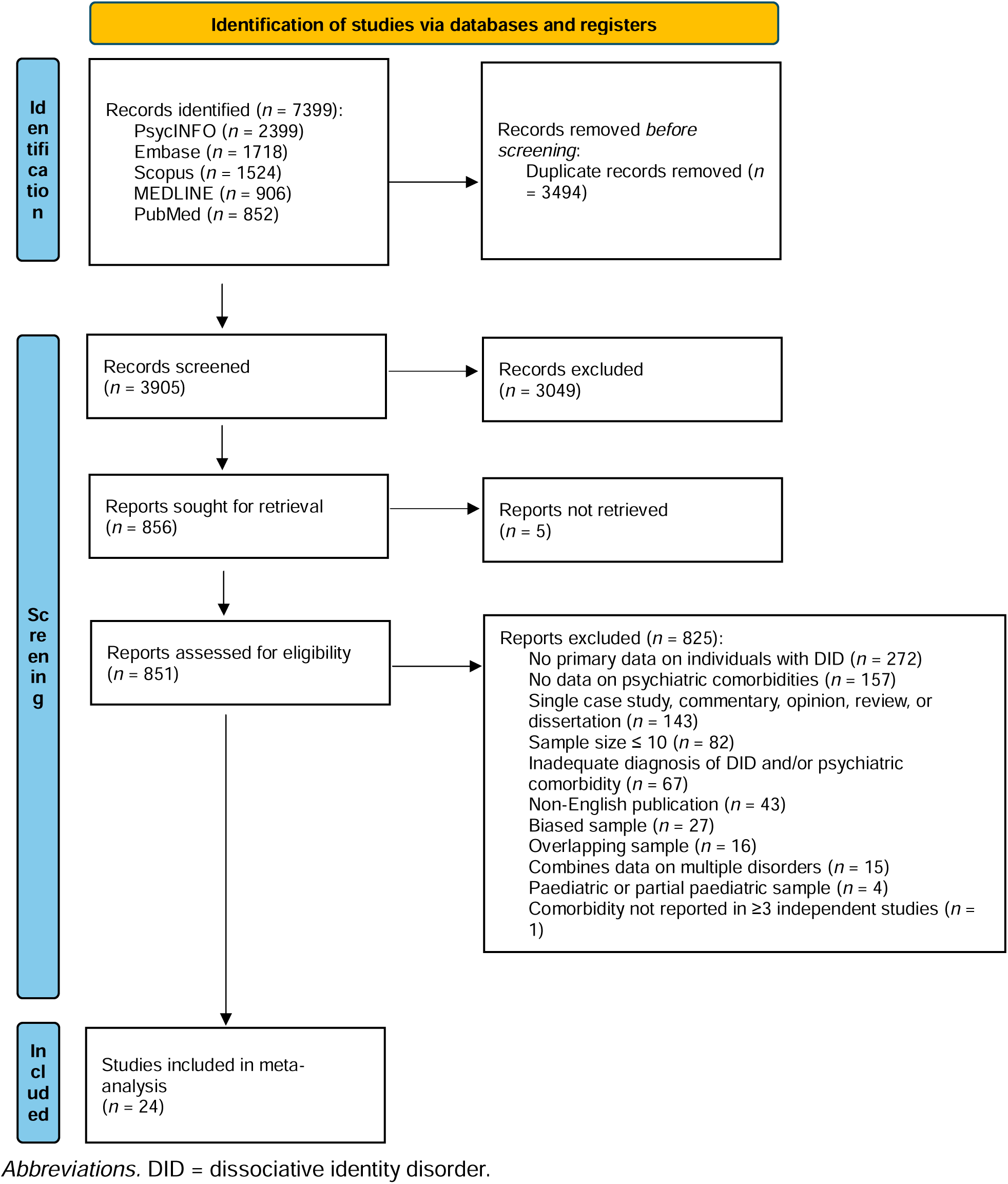
Preferred Reporting Items for Systematic reviews and Meta-Analyses flow diagram.

### Characteristics of Included Studies

Included studies were published between 1989 – 2025. The studies were mostly conducted in North America (*n* = 12), with additional studies from Europe (*n* = 7) and Asia (*n* = 6). Nine studies had small sample sizes (*n* ≤ 20), whilst the largest study (32) had a sample size of 166. Most studies (*n* = 22) recruited from treatment clinics.

The availability of demographic data for study participants varied. Mean age ranged from 21 – 44 years. Studies reporting mean age of 25 years and under (*n* = 4) were all conducted in Asia. The remaining studies (*n* = 12) reported average ages between 30 – 45 years. Across studies reporting gender (*n* = 17), most participants were female (92.0%). Only five studies reported ethnicity and largely comprised White participants (70.3%). Where averages were reported (*n* = 10), the number of years of completed education ranged from 7 – 16 years. A complete breakdown of characteristics of the included studies is presented in Table 1.

**Table 1.**
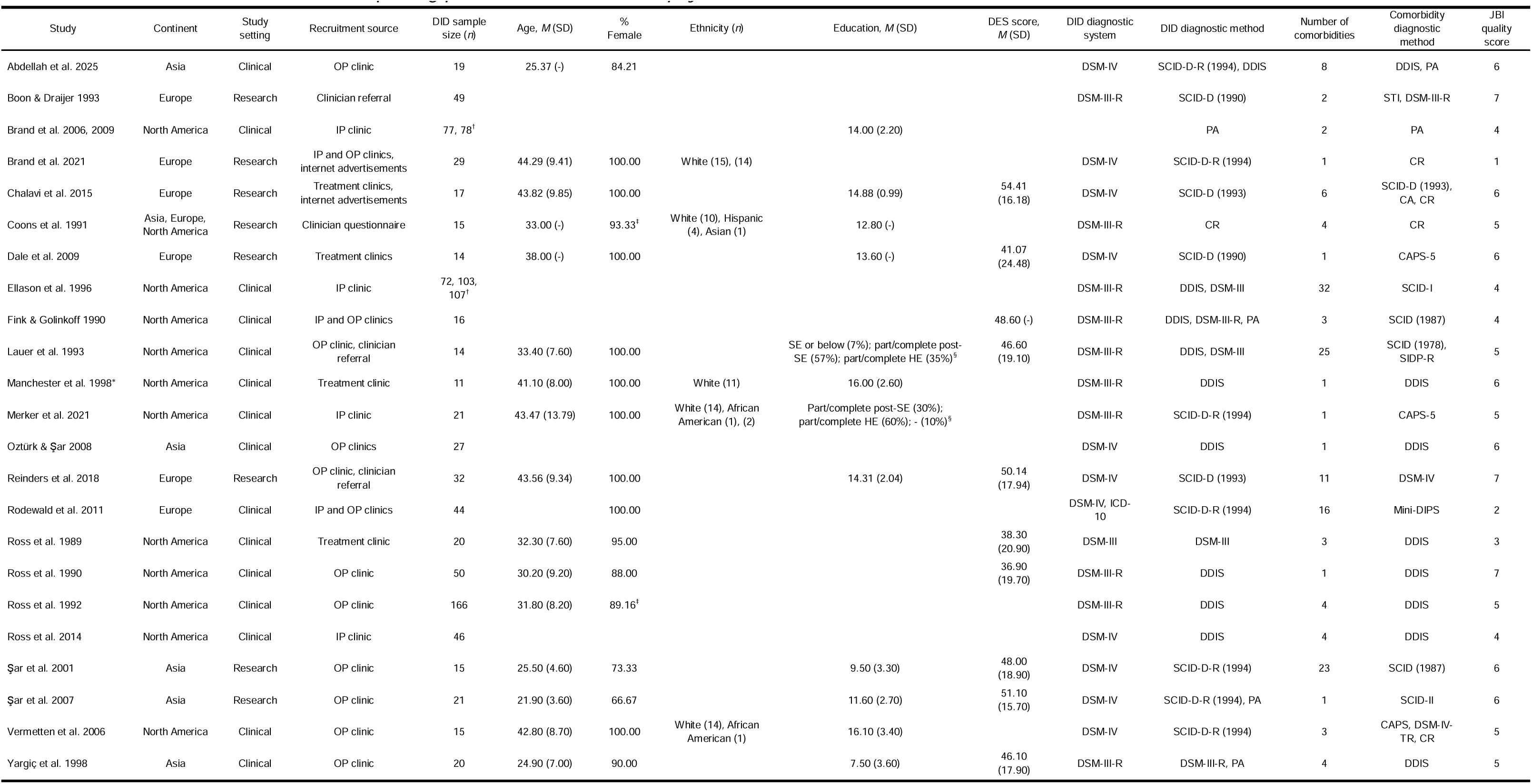
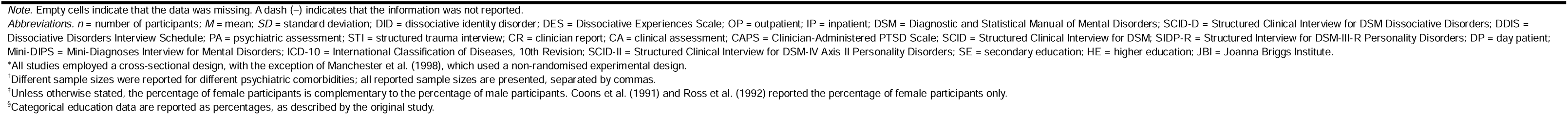
Characteristics of the included studies reporting prevalence estimates of psychiatric comorbidities in DID.

| Study | Continent | Study setting | Recruitment source | DID sample size (*n*) | Age, *M* (SD) | % Female | Ethnicity (*n*) | Education, *M* (SD) | DES score, *M* (SD) | DID diagnostic system | DID diagnostic method | Number of comorbidities | Comorbidity diagnostic method | JB1 quality score |
| --- | --- | --- | --- | --- | --- | --- | --- | --- | --- | --- | --- | --- | --- | --- |
| Abdellah et al. 2025 | Asia | Clinical | OP clinic | 19 | 25.37 (-) | 84.21 |  |  |  | DSM-IV | SCID-D-R (1994), DDIS | 8 | DDIS, PA | 6 |
| Boon & Draijer 1993 | Europe | Research | Clinician referral | 49 |  |  |  |  |  | DSM-III-R | SCID-D (1990) | 2 | STI, DSM-III-R | 7 |
| Brand et al. 2006, 2009 | North America | Clinical | IP clinic | 77, 78<sup>‡</sup> |  |  |  | 14.00 (2.20) |  |  | PA | 2 | PA | 4 |
| Brand et al. 2021 | Europe | Research | IP and OP clinics, internet advertisements | 29 | 44.29 (9.41) | 100.00 | White (15), (14) |  |  | DSM-IV | SCID-D-R (1994) | 1 | CR | 1 |
| Chalavi et al. 2015 | Europe | Research | Treatment clinics, internet advertisements | 17 | 43.82 (9.85) | 100.00 |  | 14.88 (0.99) | 54.41 (16.18) | DSM-IV | SCID-D (1993) | 6 | SCID-D (1993), CA, CR | 6 |
| Coons et al. 1991 | Asia, Europe, North America | Research | Clinician questionnaire | 15 | 33.00 (-) | 93.33<sup>‡</sup> | White (10), Hispanic (4), Asian (1) | 12.80 (-) |  | DSM-III-R | CR | 4 | CR | 5 |
| Dale et al. 2009 | Europe | Research | Treatment clinics | 14 | 38.00 (-) | 100.00 |  | 13.60 (-) | 41.07 (24.48) | DSM-IV | SCID-D (1990) | 1 | CAPS-5 | 6 |
| Ellason et al. 1996 | North America | Clinical | IP clinic | 72, 103, 107<sup>†</sup> |  |  |  |  |  | DSM-III-R | DDIS, DSM-III | 32 | SCID-I | 4 |
| Fink & Golinkoff 1990 | North America | Clinical | IP and OP clinics | 16 |  |  |  |  | 48.60 (-) | DSM-III-R | DDIS, DSM-III-R, PA | 3 | SCID (1987) | 4 |
| Lauer et al. 1993 | North America | Clinical | OP clinic, clinician referral | 14 | 33.40 (7.60) | 100.00 |  | SE or below (7%); part/complete post-SE (57%); part/complete HE (35%)<sup>§</sup> | 46.60 (19.10) | DSM-III-R | DDIS, DSM-III | 25 | SCID (1978), SIDP-R | 5 |
| Manchester et al. 1998* | North America | Clinical | Treatment clinic | 11 | 41.10 (8.00) | 100.00 | White (11) | 16.00 (2.60) |  | DSM-III-R | DDIS | 1 | DDIS | 6 |
| Merker et al. 2021 | North America | Clinical | IP clinic | 21 | 43.47 (13.79) | 100.00 | White (14), African American (1), (2) | Part/complete post-SE (30%); part/complete HE (60%); - (10%)<sup>§</sup> |  | DSM-III-R | SCID-D-R (1994) | 1 | CAPS-5 | 5 |
| Oztürk & Şar 2008 | Asia | Clinical | OP clinics | 27 |  |  |  |  |  | DSM-IV | DDIS | 1 | DDIS | 6 |
| Reinders et al. 2018 | Europe | Research | OP clinic, clinician referral | 32 | 43.56 (9.34) | 100.00 |  | 14.31 (2.04) | 50.14 (17.94) | DSM-IV | SCID-D (1993) | 11 | DSM-IV | 7 |
| Rodewald et al. 2011 | Europe | Clinical | IP and OP clinics | 44 |  | 100.00 |  |  |  | DSM-IV, ICD-10 | SCID-D-R (1994) | 16 | Mini-DIPS | 2 |
| Ross et al. 1989 | North America | Clinical | Treatment clinic | 20 | 32.30 (7.60) | 95.00 |  |  | 38.30 (20.90) | DSM-III | DSM-III | 3 | DDIS | 3 |
| Ross et al. 1990 | North America | Clinical | OP clinic | 50 | 30.20 (9.20) | 88.00 |  |  | 36.90 (19.70) | DSM-III-R | DDIS | 1 | DDIS | 7 |
| Ross et al. 1992 | North America | Clinical | OP clinic | 166 | 31.80 (8.20) | 89.16<sup>‡</sup> |  |  |  | DSM-III-R | DDIS | 4 | DDIS | 5 |
| Ross et al. 2014 | North America | Clinical | IP clinic | 46 |  |  |  |  |  | DSM-IV | DDIS | 4 | DDIS | 4 |
| Şar et al. 2001 | Asia | Research | OP clinic | 15 | 25.50 (4.60) | 73.33 |  | 9.50 (3.30) | 48.00 (18.90) | DSM-IV | SCID-D-R (1994) | 23 | SCID (1987) | 6 |
| Şar et al. 2007 | Asia | Research | OP clinic | 21 | 21.90 (3.60) | 66.67 |  | 11.60 (2.70) | 51.10 (15.70) | DSM-IV | SCID-D-R (1994), PA | 1 | SCID-II | 6 |
| Vermetten et al. 2006 | North America | Clinical | OP clinic | 15 | 42.80 (8.70) | 100.00 | White (14), African American (1) | 16.10 (3.40) |  | DSM-IV | SCID-D-R (1994) | 3 | CAPS, DSM-IV-TR, CR | 5 |
| Yargıç et al. 1998 | Asia | Clinical | OP clinic | 20 | 24.90 (7.00) | 90.00 |  | 7.50 (3.60) | 46.10 (17.90) | DSM-III-R | DSM-III-R, PA | 4 | DDIS | 5 |

*Note.* Empty cells indicate that the data was missing. A dash (–) indicates that the information was not reported.
*Abbreviations.* *n* = number of participants; *M* = mean; *SD* = standard deviation; DID = dissociative identity disorder; DES = Dissociative Experiences Scale; OP = outpatient; IP = inpatient; DSM = Diagnostic and Statistical Manual of Mental Disorders; SCID-D = Structured Clinical Interview for DSM Dissociative Disorders; DDIS = Dissociative Disorders Interview Schedule; PA = psychiatric assessment; STI = structured trauma interview; CR = clinician report; CA = clinical assessment; CAPS = Clinician-Administered PTSD Scale; SCID = Structured Clinical Interview for DSM; SIDP-R = Structured Interview for DSM-III-R Personality Disorders; DP = day patient; Mini-DIPS = Mini-Diagnoses Interview for Mental Disorders; ICD-10 = International Classification of Diseases, 10th Revision; SCID-II = Structured Clinical Interview for DSM-IV Axis II Personality Disorders; SE = secondary education; HE = higher education; JBI = Joanna Briggs Institute.
\*All studies employed a cross-sectional design, with the exception of Manchester et al. (1998), which used a non-randomised experimental design.
<sup>†</sup>Different sample sizes were reported for different psychiatric comorbidities; all reported sample sizes are presented, separated by commas.
<sup>‡</sup>Unless otherwise stated, the percentage of female participants is complementary to the percentage of male participants. Coons et al. (1991) and Ross et al. (1992) reported the percentage of female participants only.
<sup>§</sup>Categorical education data are reported as percentages, as described by the original study.

Across 556 participants (*k* = 9), the pooled mean DES score was 46.48 (95% CI 42.88, 50.07), closely replicating the estimate reported by Lyssenko et al. (27) (48.7 (95% CI 46.4, 50.9).

### Risk of Bias Assessment

JBI quality scores ranged from 1 – 7, with three studies of high quality, three of low quality, and the remaining 18 studies rated as moderate quality. The most common methodological limitations related to appropriateness of the sample frame, adequacy and management of response rates, and descriptive detail of the study participants. The results of the risk of bias assessment are presented in Supplementary Table 4.

### Psychiatric Comorbidities

Thirty-six distinct psychiatric comorbidities were reported across the included studies. The most frequently reported comorbidities were somatic symptom disorder (*n* = 14), PTSD (*n* = 13), borderline personality disorder (*n* = 13), and major depressive disorder (*n* = 12). Three comorbidities – attention-deficit/hyperactivity disorder, body dysmorphic disorder, and sadistic personality disorder – were only reported in a single study. All reported psychiatric comorbidities and their associated prevalence count are reported in Supplementary Table 5.

### Meta-Analysis

Meta-analyses were conducted for 23 psychiatric comorbidities reported by at least three independent studies. Across the included studies, 858 individuals with DID contributed to the prevalence analyses. Pooled prevalence estimates are presented in Table 2 and graphically represented in Figure 2, with corresponding forest plots provided in Supplementary Figures 1-23. Moderate to substantial between-study heterogeneity was observed for most comorbidities, although heterogeneity was low or absent for several disorders.

**Figure 2.**
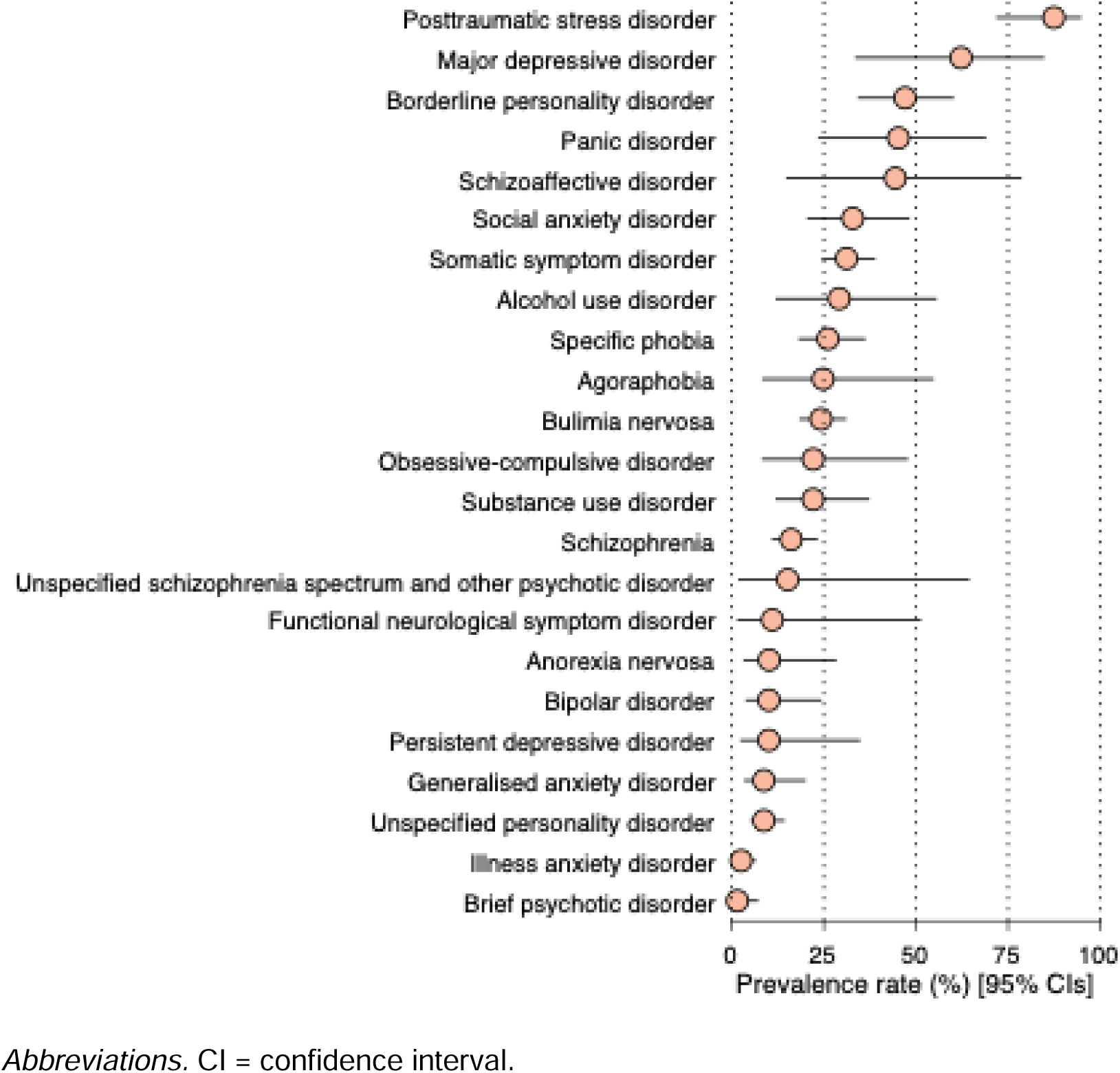
Pooled prevalence estimates of each psychiatric disorder among individuals with dissociative identity disorder (DID).

**Table 2.** Pooled prevalence of psychiatric comorbidities among individuals with DID.

| Disorder | *k* | *N* | Back-transformed<br>pooled prevalence,<br>% (95% CI) | $\tau^2$ (SE) | $I^2$ (%) | $Q$ (*p*) | Funnel plot<br>asymmetry (*p*) |
| --- | --- | --- | --- | --- | --- | --- | --- |
| Posttraumatic stress disorder | 13 | 420 | 87.44 (71.79, 94.96) | 2.34 (1.61) | 82.37 | 68.05<br>($<.001^{***}$) | .085 |
| Major depressive disorder | 12 | 523 | 62.64 (33.45, 84.84) | 3.90 (2.38) | 94.08 | 185.70<br>($<.001^{***}$) | .17 |
| Borderline personality<br>disorder | 13 | 593 | 47.05 (34.21, 60.32) | 0.78 (0.49) | 86.40 | 88.22<br>($<.001^{***}$) | .52 |
| Panic disorder | 5 | 212 | 45.24 (23.42, 69.08) | 1.08 (1.02) | 87.37 | 31.67<br>($<.001^{***}$) | .004** |
| Schizoaffective disorder | 3 | 136 | 44.40 (14.72, 78.70) | 1.41 (1.94) | 80.93 | 10.49 (.005**) | .47 |
| Social anxiety disorder | 4 | 180 | 32.85 (20.41, 48.28) | 0.25 (0.36) | 63.09 | 8.13 (.043*) | .046* |
| Somatic symptom disorder | 14 | 577 | 31.17 (24.34, 38.96) | 0.20 (0.16) | 59.97 | 32.48 (.002**) | $<.001^{***}$ |
| Alcohol use disorder | 4 | 151 | 29.15 (11.91, 55.63) | 0.83 (1.08) | 73.17 | 11.18 (.011*) | .002** |
| Specific phobia | 6 | 229 | 26.11 (18.09, 36.33) | 0.14 (0.22) | 44.92 | 9.08 (.11) | .22 |
| Agoraphobia | 3 | 166 | 24.79 (8.20, 54.86) | 1.12 (1.46) | 89.29 | 18.67<br>($<.001^{***}$) | .58 |
| Bulimia nervosa | 4 | 180 | 24.23 (18.33, 31.09) | 0.000 (0.16) | 0.00 | 2.98 (.40) | .12 |
| Obsessive-compulsive disorder | 6 | 231 | 22.10 (8.12, 47.88) | 1.71 (1.61) | 88.56 | 43.70<br>($<.001^{***}$) | $<.001^{***}$ |
| Substance use disorder | 9 | 443 | 22.10 (11.88, 37.40) | 0.81 (0.77) | 85.02 | 53.39<br>($<.001^{***}$) | $<.001^{***}$ |
| Schizophrenia | 4 | 157 | 16.11 (10.83, 23.34) | 0.01 (0.39) | 1.47 | 3.05 (.39) | .092 |
| Unspecified schizophrenia spectrum and other psychotic disorder | 3 | 136 | 15.19 (1.73, 64.52) | 3.81 (4.28) | 91.03 | 22.30<br>($<.001^{***}$) | .75 |
| Functional neurological symptom disorder | 3 | 95 | 11.01 (1.41, 51.43) | 2.94 (3.76) | 83.28 | 11.97 (.003**) | $<.001^{***}$ |
| Anorexia nervosa | 4 | 180 | 10.16 (3.14, 28.46) | 0.98 (1.37) | 65.02 | 8.58 (.035*) | .007** |
| Bipolar disorder | 4 | 180 | 10.07 (3.75, 24.36) | 0.51 (0.99) | 44.75 | 5.43 (.14) | .039* |
| Persistent depressive disorder | 6 | 229 | 10.07 (2.28, 34.89) | 3.01 (2.82) | 85.37 | 34.17<br>($<.001^{***}$) | .040* |
| Generalised anxiety disorder | 4 | 185 | 8.71 (3.32, 20.10) | 0.61 (0.89) | 57.09 | 6.99 (.072) | .10 |
| Unspecified personality disorder | 3 | 152 | 8.71 (5.10, 14.37) | 0.000 (0.36) | 0.00 | 0.44 (.80) | .99 |
| Illness anxiety disorder | 3 | 166 | 2.53 (0.95, 6.55) | 0.000 (1.26) | 0.00 | 0.40 (.82) | .72 |
| Brief psychotic disorder | 3 | 141 | 1.52 (0.31, 7.25) | 0.000 (2.04) | 0.00 | 1.11 (.58) | .30 |

*Note.* $p < .05^*$ , $p < .01^{**}$ , $p < .001^{***}$ .
*Abbreviations.* $k$ = number of studies; $N$ = total sample size.

#### Trauma-and stressor-related disorders

PTSD prevalence was reported in 13 studies (*N* = 420) with a pooled prevalence of 87.44% (95% CI 71.79, 94.96), the highest estimate of all analysed comorbidities. Substantial between-study heterogeneity was observed. The 95% CIs did not overlap with those of most other comorbidities (except major depressive disorder and schizoaffective disorder), suggesting a descriptive marker of difference between the prevalence estimates of PTSD and other psychiatric comorbidities.

#### Depressive disorders

Major depressive disorder (MDD) was reported in 12 studies (*N*=523), with a pooled prevalence of 62.64% (95% CI 33.45, 84.84), albeit with the greatest heterogeneity of all comorbidities. Persistent depressive disorder (PDD) was reported in six studies (*N* = 229) with a pooled prevalence of 10.07% (95% CI 2.28, 34.89) and substantial heterogeneity.

#### Personality disorders

BPD was reported in 13 studies (*N* = 593). A pooled prevalence of 47.05% (95% CI 34.21, 60.32) was estimated, with substantial heterogeneity. Unspecified personality disorder was reported in three studies (*N* = 152), with a pooled prevalence of 8.71% (95% CI 5.10, 14.37). No between-study heterogeneity was observed.

#### Anxiety disorders

Anxiety disorders were the most frequently represented diagnostic category across the meta-analyses. Pooled prevalence estimates for anxiety disorders ranged from 8.71% for generalised anxiety disorder (GAD; 95% CI 3.32, 20.10; *N* = 185), 24.79% for agoraphobia (95% CI 8.20, 54.86; *N* = 166), 26.11% for specific phobia (95% CI 18.09, 36.33; *N* = 229), 32.85% for social anxiety disorder (95% CI 20.41, 48.28; *N* = 180), to 45.24% for panic disorder (95% CI 23.42, 69.08; *N* = 212). Panic disorder showed substantial heterogeneity, as did agoraphobia. Moderate heterogeneity was observed for social anxiety disorder (SAD) and generalised anxiety disorder, whilst heterogeneity was low for specific phobia.

#### Schizophrenia spectrum and other psychotic disorders

Pooled prevalence estimates varied across psychotic disorders, from 1.52% for brief psychotic disorder (95% CI 0.31, 7.25; *N* = 141) to 44.40% for schizoaffective disorder (95% CI 14.72, 78.70; *N* = 136), with unspecified schizophrenia spectrum and other psychotic disorder at 15.19% (95% CI 1.73, 64.52; *N* = 136) and schizophrenia at 16.11% (95% CI 10.83, 23.34; *N* = 157). Substantial heterogeneity was observed for unspecified schizophrenia spectrum, other psychotic disorder, and schizoaffective disorder but not for schizophrenia or brief psychotic disorder.

#### Somatic symptom and related disorders

The pooled prevalence for somatic symptom disorder was 31.17% (95% CI 24.34, 38.96, *N* = 577), with moderate between-study heterogeneity. Functional neurological symptom disorder was reported in three studies (*N* = 95), with a pooled prevalence of 11.01% (95% CI 1.41, 51.43) and substantial heterogeneity. Illness anxiety disorder had a pooled prevalence of 2.53% (95% CI 0.95, 6.55), with no observed heterogeneity.

#### Substance-related and addictive disorders

Alcohol use disorder was reported in four studies (*N* = 151), with a pooled prevalence of 29.15% (95% CI 11.91, 55.63) and substantial heterogeneity. Similarly, substance use disorder was reported in nine studies (*N* = 443), with a pooled prevalence of 22.10% (95% CI 11.88, 37.40) and substantial heterogeneity.

#### Feeding and eating disorders

Among four studies (*N* = 180), the pooled prevalence estimates were 24.23% for bulimia nervosa (95% CI 18.33, 31.09) and 10.16% for anorexia nervosa (95% CI 3.14, 28.46). The studies displayed no and moderate heterogeneity, respectively.

#### Obsessive-compulsive and related disorders

Obsessive-compulsive disorder (OCD) was reported in six studies (*N* = 231), with a pooled prevalence of 22.10% (95% CI 8.12, 47.88). Substantial between-study heterogeneity was observed.

#### Bipolar and related disorders

Bipolar disorder was reported in four studies (*N* = 180), with a pooled prevalence of 10.07% (95% CI 3.75, 24.36). Moderate heterogeneity was observed.

### Funnel Plot Asymmetry

Funnel plots (see Supplementary Figures 24-46) and tests of funnel plot asymmetry were used to assess potential small-study effects, including possible publication bias. Significant funnel plot asymmetry was observed for ten comorbidities. Visual inspection suggested that smaller studies tended to report lower prevalence estimates, most clearly for substance use disorder, bipolar disorder, PDD, anorexia nervosa, and OCD. In contrast, the direction of asymmetry was less consistent for panic disorder, SAD, SSD, alcohol use disorder, and functional neurological symptom disorder. These findings suggest that multiple prevalence estimates are conservative but do not establish publication bias, particularly given the small number of studies and substantial heterogeneity.

### Meta-Regression

Given the substantial between-study heterogeneity, potential moderators were evaluated using random-effects meta-regressions in analyses of four disorders (*k* ≥ 10). Ten significant associations were identified across the four comorbidities, with full results presented in Table 3.

**Table 3.**
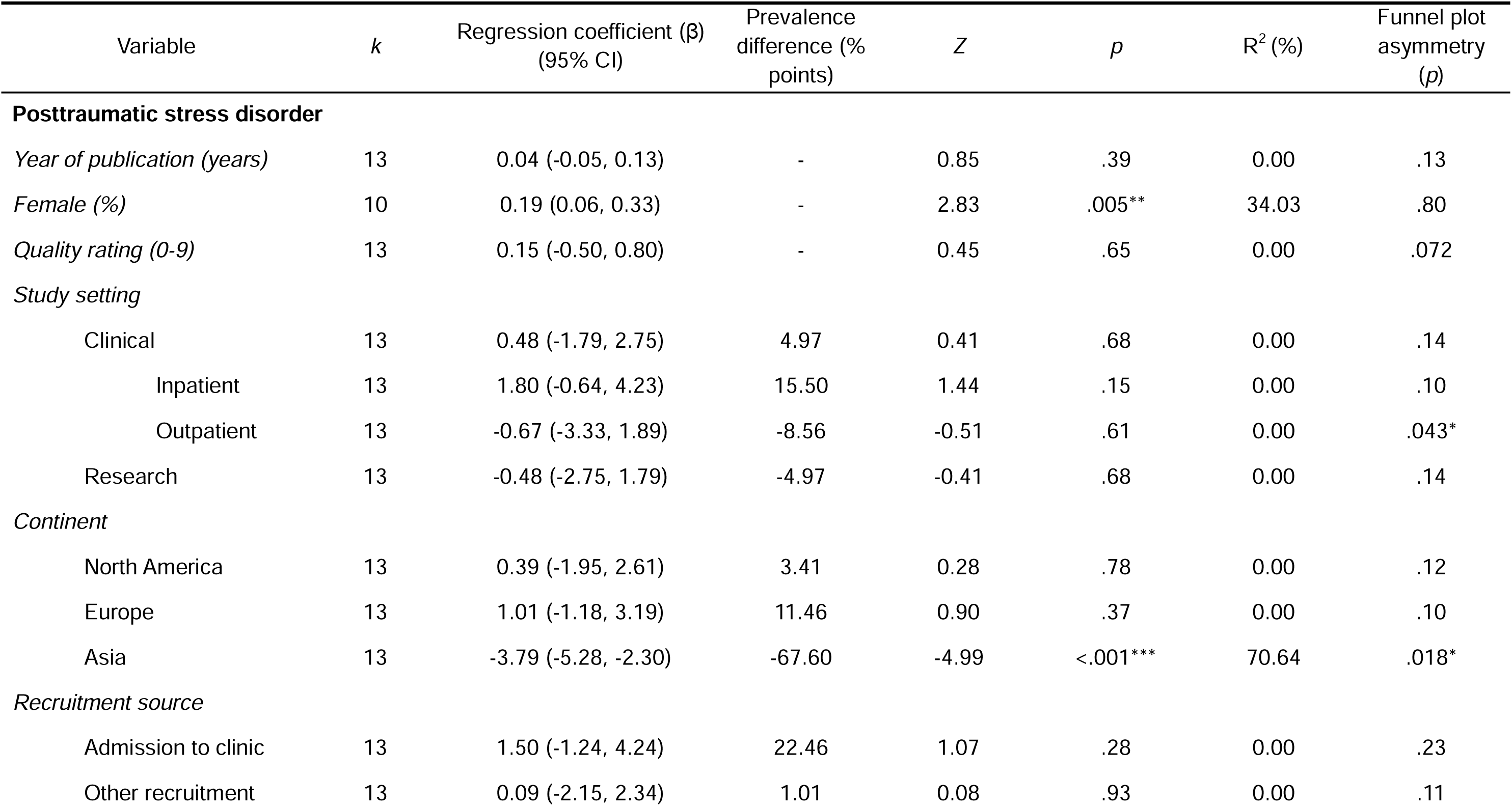
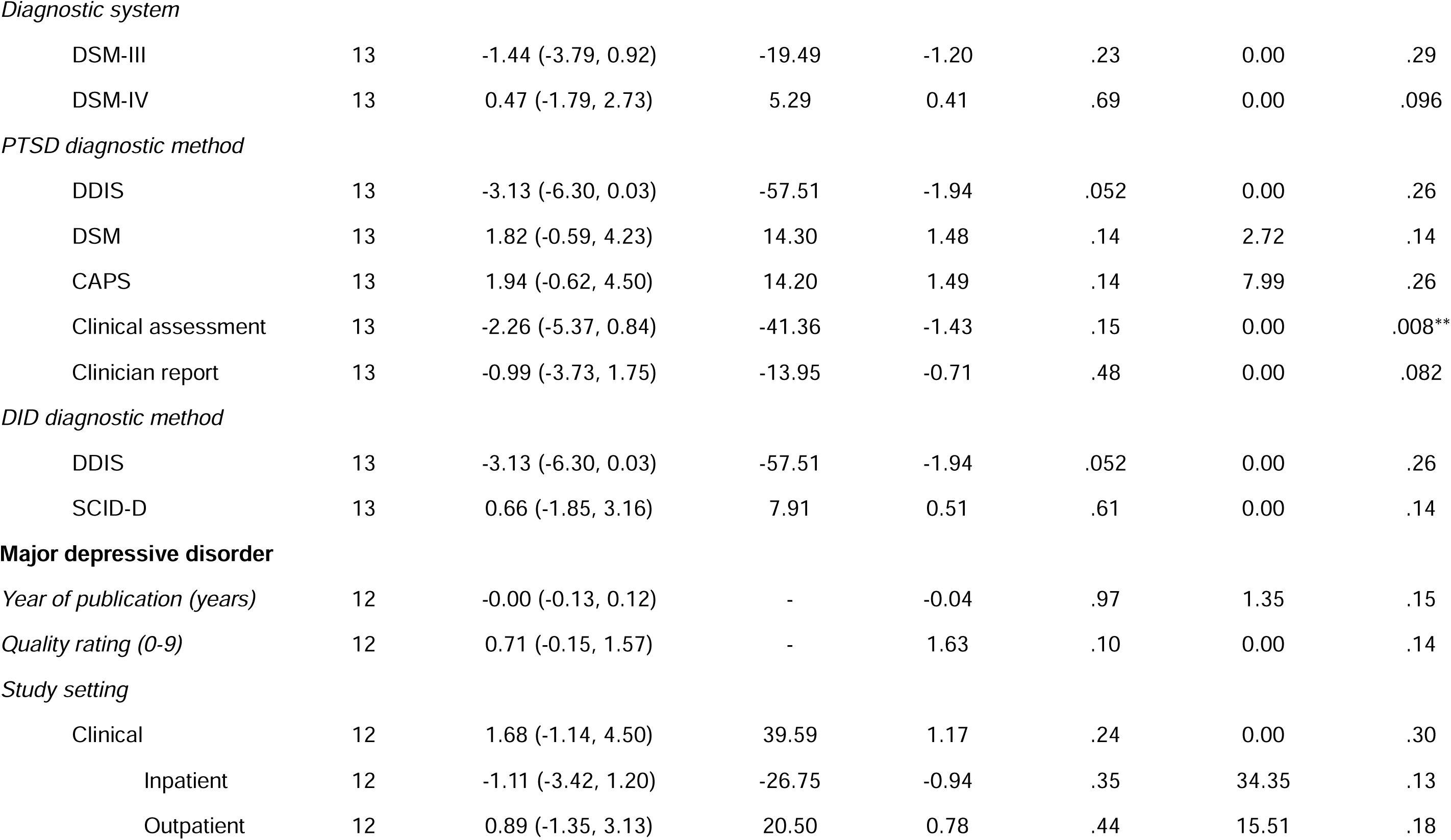
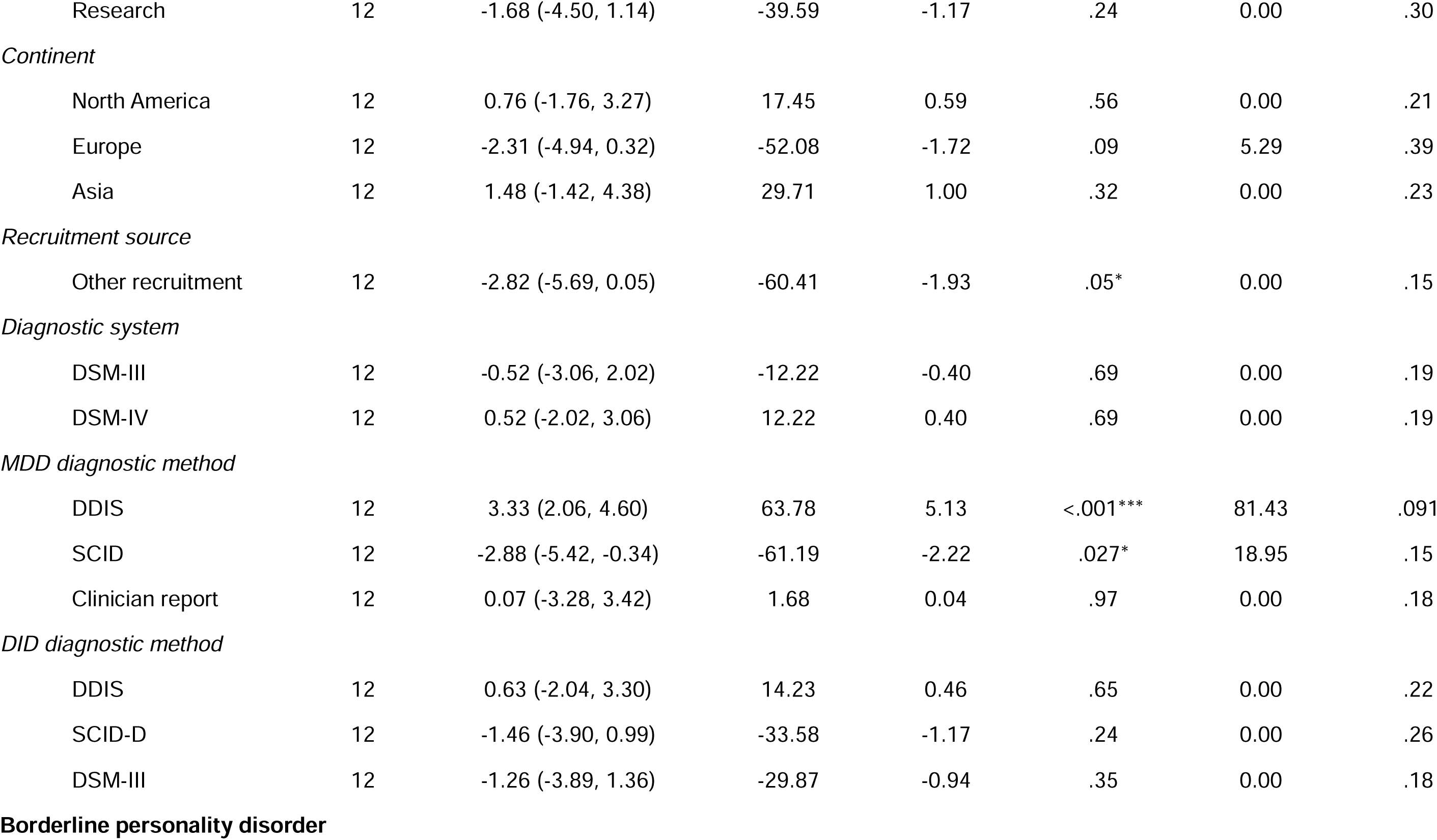
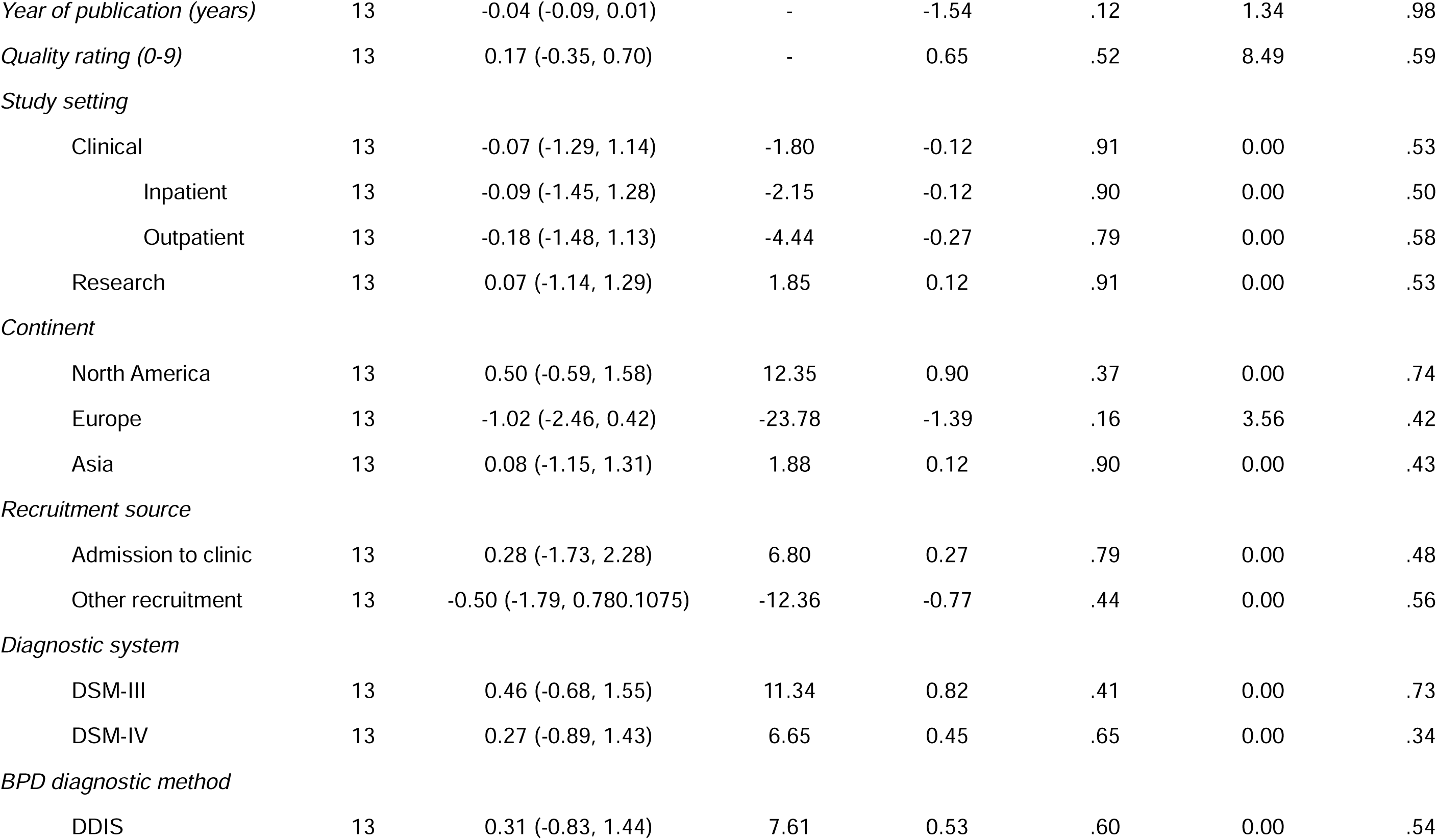
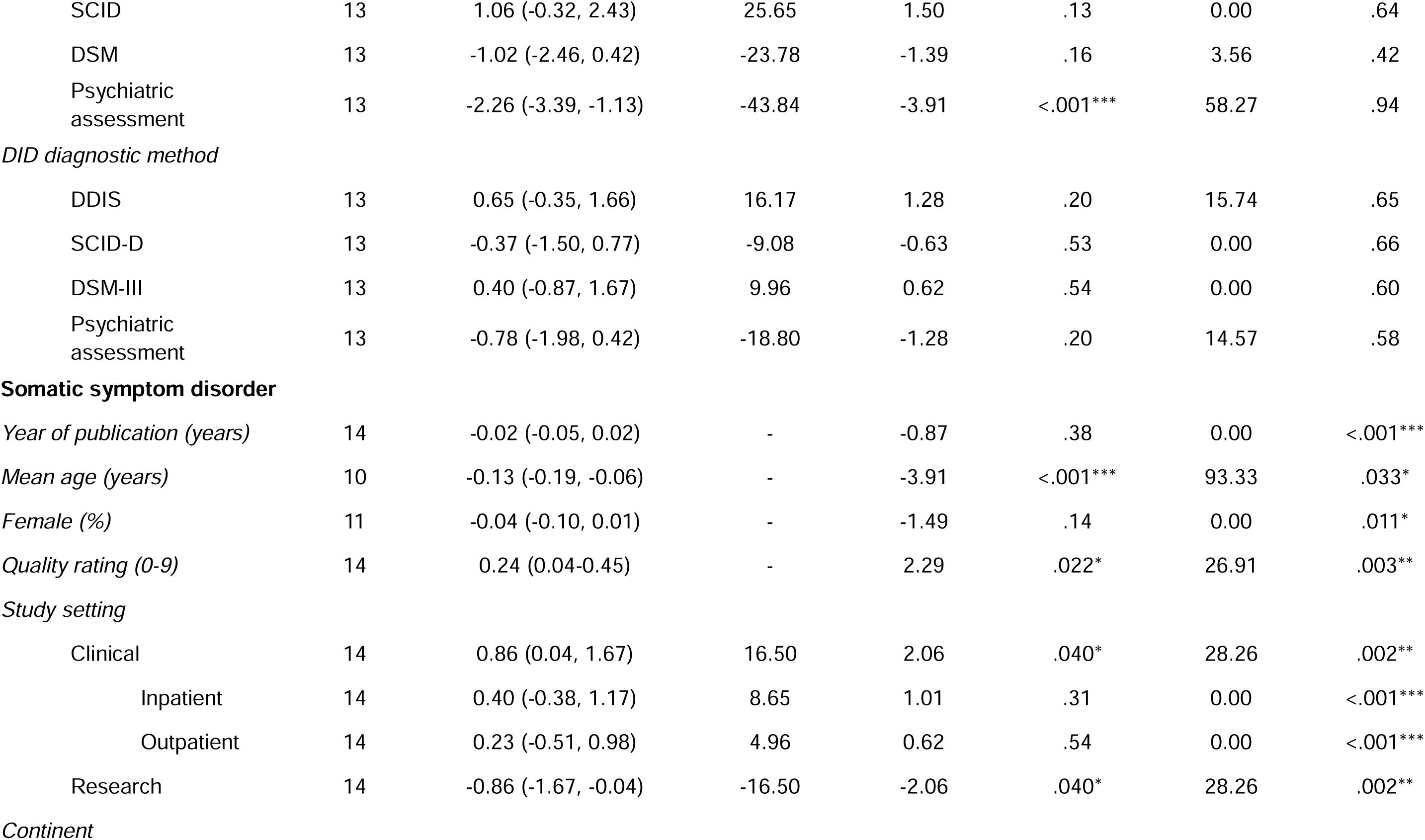
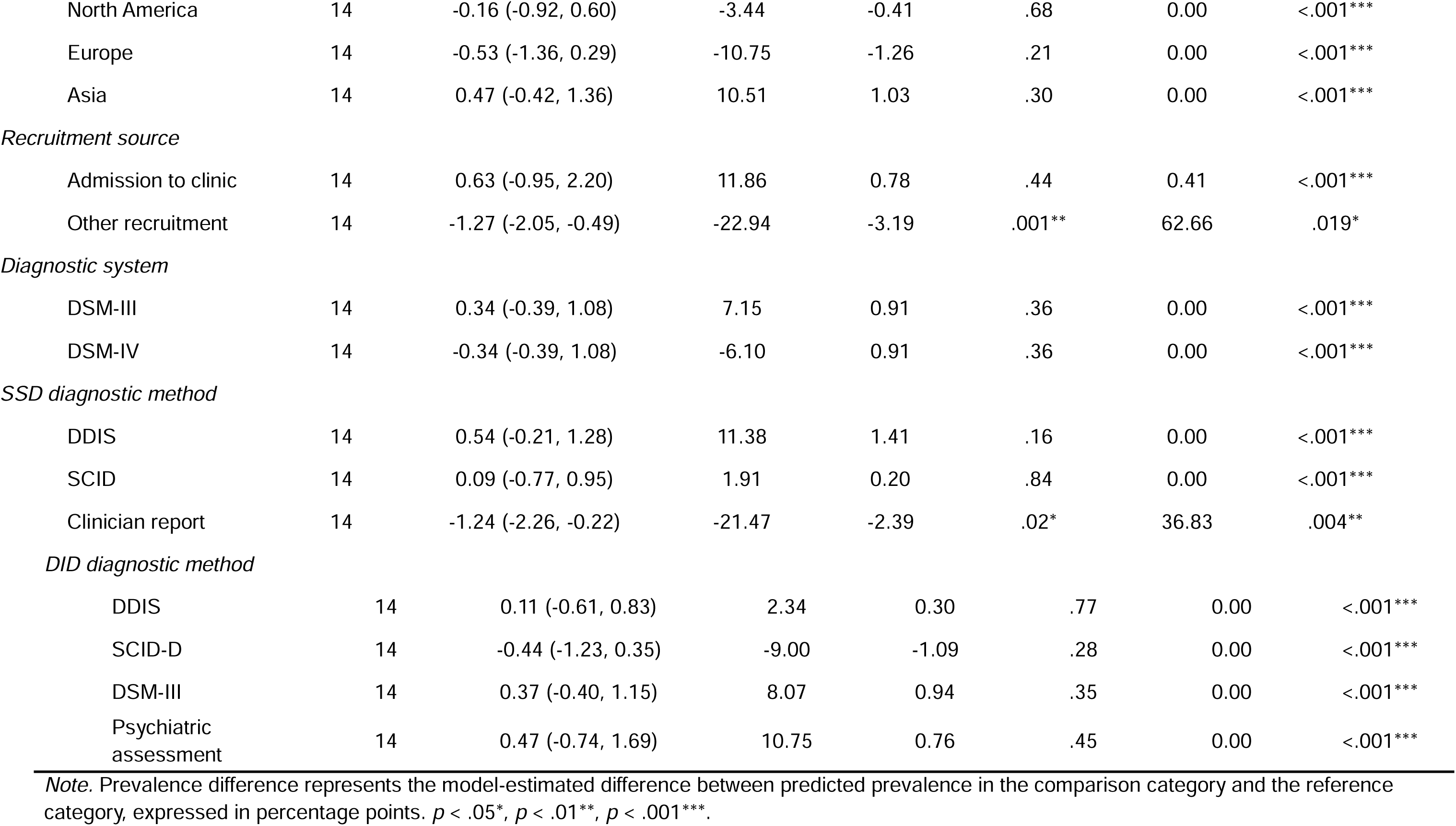
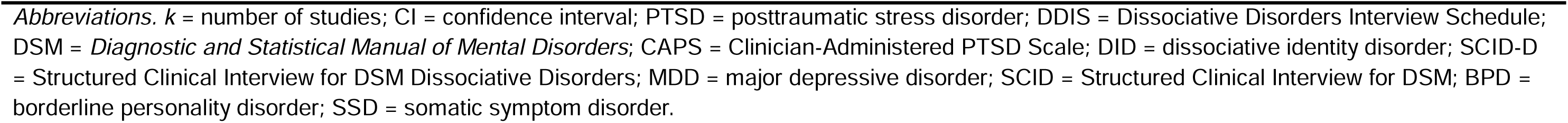
Meta-regression analyses of moderators of psychiatric comorbidity prevalence in DID.

#### Posttraumatic stress disorder

Female proportion was positively associated with PTSD prevalence (β = 0.19, 95% CI 0.06, 0.33), accounting for 34.03% of the variance. Studies conducted in Asia had a lower predicted PTSD prevalence than non-Asian studies (prevalence difference = -67.60 percentage points; β = -3.79, 95% CI -5.28, -2.30), accounting for 70.64% of the variance. DDIS-based diagnosis showed marginal evidence of lower prevalence for both PTSD and DID diagnosis (β = -3.13, 95% CI -6.30, 0.03).

#### Major depressive disorder

Recruitment source was marginally associated with MDD prevalence, with other recruitment methods yielding lower estimates (prevalence difference = -60.41 percentage points; β = -2.82, 95% CI -5.69, 0.05). MDD diagnostic method also moderated prevalence: DDIS was associated with higher estimates (prevalence difference = 63.78 percentage points; β = 3.33, 95% CI 2.06, 4.60) and SCID with lower estimated (prevalence difference = -61.19 percentage points; β = -2.88, 95% CI -5.42, -0.34)

#### Borderline personality disorder

Studies that used psychiatric assessment to diagnose BPD reported a lower pooled prevalence than studies using other diagnostic methods (prevalence difference = −43.84 percentage points; β = −2.26, 95% CI −3.39 to −1.13), with diagnostic method accounting for 58.27% of between-study heterogeneity.

#### Somatic symptom disorder

Mean age was negatively associated with SSD prevalence (β = -0.13, 95% CI -0.19, -0.06), accounting for 93.33% of heterogeneity, and positively associated with quality (β = 0.24, 95% CI 0.04, 0.45). Clinical settings yielded higher prevalence than research settings (prevalence difference = 16.50 percentage points; β = 0.86, 95% CI 0.04, 1.67), while other recruitment methods (prevalence difference = -22.94 percentage points; β = -1.27, 95% CI - 2.05, -0.49), and clinician-reported SSD diagnosis (prevalence difference = -21.47 percentage points; β = -1.24, 95% CI -2.26, -0.22) were associated with lower predicted prevalence.

### Sensitivity Analyses

Sensitivity analyses assessed the robustness of pooled estimates to outlying/influential studies and methodological restrictions. Of nine comorbidities with *k* ≥ 5, seven had one or more identified influential or outlying studies. Supplementary Table 6 reports the repeated analyses following exclusion of reported outliers and influential studies. Excluding these studies produced relatively stable estimates for PTSD, SSD and specific phobia (<4% change), while moderate increases were observed for BPD (5.19%) and panic disorder (13.25%) alongside moderate decreases for substance use disorder (-14.51%) and persistent depressive disorder (-4.76%). Heterogeneity generally decreased, most notably for specific phobia and SUD, although substantial heterogeneity remained for PTSD, BPD, panic disorder, SSD and PDD.

Additional methodological sensitivity analyses restricting sample size, JBI quality, and structured diagnostic methods yielded broadly comparable estimates (Supplementary Table 7). PTSD was particularly robust (85.81–93.52%). MDD ranged from 56.59–72.73% and PDD from 5.42–14.43%. Notable increases occurred for OCD with structured comorbidity interviews (36.03%) and SUD in studies with n ≥ 20 (35.92%). Heterogeneity remained substantial in most analyses but decreased for SSD with structured interviews and was eliminated for specific phobia.

### Certainty of Evidence

The overall certainty of evidence varied across 23 prevalence estimates, with certainty rated high for seven, moderate for four, low for three, and very low for nine. Risk of bias across estimates was assessed as low, and no estimate was downgraded on the basis of indirectness, as the evidence directly addressed psychiatric comorbidity prevalence in individuals with DID. Downgrading primarily reflected substantial between-study heterogeneity and imprecision. Supplementary Table 8 summarises the findings of the GRADE assessment.

## Discussion

Psychiatric comorbidity across multiple DSM-5 diagnostic categories was highly prevalent among individuals with DID. PTSD had the highest pooled prevalence estimate (87.44%), followed by MDD (62.64%). Just under half of individuals with DID met criteria for comorbid BPD (47.05%), panic disorder (45.24%), or schizoaffective disorder (44.40%). However, moderate-to-high heterogeneity across many disorders indicates that the precise magnitude of individual comorbidities remains uncertain. These findings demonstrate a pronounced and diverse burden of psychiatric comorbidity in DID, highlighting its importance for understanding overall symptom burden, supporting careful consideration of differential diagnosis, and informing comprehensive clinical assessment and treatment planning.

The estimated prevalence of psychiatric comorbidity in DID was generally higher than in a large nationwide German epidemiological study that was not included in this review (10). The present analyses yielded higher estimates for bipolar disorder (10.07% vs. 2.7%), schizophrenia (16.11% vs. 9.2%), and OCD (22.10% vs. 4.5%), whilst MDD (62.64% vs. 60.5%) and substance use disorder (22.10% vs. 18.4%) were comparable. Differences may partly reflect variation in the range and assessment of comorbidities rather than overall burden of psychopathology. The level of dissociation in the present sample was also comparable to that observed in a meta-analysis of dissociation in DID (27), providing support for population comparability.

The extent of comorbidity raises the question of whether DID represents an unusually high psychiatric burden relative to other psychiatric populations. High levels of comorbidity are, however, well established across psychiatric disorders. In a nationally representative US sample (33), 72.1% of individuals with lifetime MDD had at least one other lifetime psychiatric disorder, whilst an English population-based study found that 78.5% of individuals with probable PTSD had at least one psychiatric comorbidity (34). Similarly, approximately half of individuals with BPD met criteria for a mood, anxiety, or substance use disorder in a large US sample (35). More directly, a meta-analysis of 57 studies found that 52% (95% CI 48–56%) of individuals with current PTSD had comorbid MDD (36). Thus, the high level of comorbidity observed in DID should be understood within the broader context of psychiatric multimorbidity, rather than necessarily indicating uniquely elevated psychopathological burden. Nevertheless, the present findings demonstrate a particularly broad pattern spanning PTSD, depressive, personality, anxiety, somatic and schizophrenia spectrum disorders all substantially represented. Whether this burden exceeds that observed in other severely affected populations remains unclear given differences in diagnostic methods, assessment timeframes, samples, and disorders assessed. Future direct comparative studies using consistent diagnostic procedures across DID and matched samples would permit more meaningful comparisons.

The high prevalence of PTSD (87.44%) in DID is consistent with conceptualisations of DID as a disorder characterised by developmental and chronic trauma. PTSD had the highest pooled prevalence of all psychiatric comorbidities examined in the present review, with all other conditions occurring at lower rates. This result is also consistent with the interpretation of DID as a complex post-traumatic disorder originating in childhood (11, 37), broader evidence for associations between trauma exposure and dissociation (38, 39), and the substantial overlap in trauma-related symptomatology between DID and PTSD (11, 39). The high prevalence of PTSD has implications for the clinical complexity and treatment burden associated with DID, as clinicians may need to address substantial trauma-related symptomatology alongside multiple other comorbid psychiatric conditions. It also supports consideration of trauma-related processes in understanding the development and clinical presentation of DID.

Among the anxiety disorders examined, panic disorder had the highest pooled prevalence (45.24%), followed by social anxiety disorder (32.85%). The almost one-in-two prevalence of panic disorder may partly reflect the phenomenological overlap between panic and dissociative experiences, as depersonalisation and derealisation commonly occur during panic attacks (40), whilst childhood trauma, dissociation, and panic severity are also interrelated (41, 42). Panic symptoms are also common among people with PTSD; in a US population-based sample, 35% of individuals with PTSD reported past-year panic attacks, which were associated with greater PTSD-related disability (43). The prominence of social anxiety may relate to interpersonal trauma and shame-related processes described in DID, as greater shame, withdrawal and concerns about rejection or negative evaluation are commonly reported among individuals with dissociative disorders (44, 45). These findings suggest that anxiety-related psychopathology is prominent in DID and underline the importance of assessing these alongside dissociative phenomena, particularly where panic and dissociation may present with overlapping experiences. The high prevalence of anxiety disorders may also contribute to the complexity of clinical presentations and treatment needs in DID, reinforcing the importance of comprehensive assessment and formulation across anxiety and dissociative symptom domains.

MDD prevalence (62.64%) was markedly higher than PDD (10.07%), which may partly reflect differences in their diagnostic conceptualisation, with PDD’s more restrictive diagnostic criteria (e.g., a minimum duration of two years) likely contributing to its lower prevalence. The DSM-5 also consolidated several chronic depressive presentations within the PDD diagnosis, which may contribute to diagnostic heterogeneity (46). Further, diagnostic method significantly moderated MDD prevalence, suggesting that assessment may influence estimates. Nevertheless, a community-based study demonstrated a similar pattern, with MDD occurring more frequently than PDD even when assessed using the same diagnostic method (47). The difference between these disorders may therefore reflect genuine variation in depressive course alongside differences in measurement.

BPD was highly prevalent (47.05%) and included the largest contributing sample. This estimate may partly reflect the substantial clinical and phenomenological overlap between BPD and DID, with historical diagnostic overlap due to the shared associations with trauma, emotion dysregulation, and dissociation (48). However, evidence that the characteristics and function of dissociative experiences differ between BPD and DID suggests that the findings may also represent genuine comorbidity (49). BPD was assessed more frequently than other personality disorders, limiting conclusions about broader personality pathology.

The higher prevalence of bulimia nervosa (24.23%) than anorexia nervosa (10.16%) in DID is notable because both were derived from the same studies, limiting the possibility that the difference is attributable to methodological variation. The higher prevalence of bulimia nervosa may reflect a closer association between dissociation and binge–purge psychopathology. Dissociation has been reported more frequently in binge-purging than restricting presentations of anorexia nervosa (50), whilst a recent scoping review found lower dissociation in restricting-type anorexia nervosa than binge-purging anorexia nervosa, bulimia nervosa and binge-eating disorder (51). The review further identified that dissociation may mediate the relationship between trauma and binge eating. Thus, the higher observed prevalence of bulimia nervosa may reflect the greater prominence of binge–purge psychopathology in presentations characterised by dissociation. Nevertheless, further research is warranted to clarify this distinction.

Heterogeneity varied considerably across the identified schizophrenia spectrum and psychotic disorders. Notably, the pooled prevalence of schizoaffective disorder was substantially higher than that of schizophrenia (44.40% vs. 16.11%), despite schizoaffective disorder being less common than schizophrenia in the general population (52). This difference may warrant further investigation, particularly given the affective component of schizoaffective disorder, although the present data cannot establish whether this reflects differences in diagnostic classification, clinical presentation, or underlying mechanisms. Further, the pronounced heterogeneity for unspecified schizophrenia spectrum and other psychotic disorder may partly reflect phenomenological overlap between psychotic and dissociative experiences. Psychotic-like symptoms, particularly auditory hallucinations, are well documented in DID, with evidence demonstrating overlap between dissociative and schizophrenia spectrum presentations (53), whereas negative symptoms are more prominent in schizophrenia than DID (54). Less specific diagnostic classification may therefore be particularly sensitive to differences in how psychotic experiences are conceptualised.

Substantial heterogeneity across several comorbidities suggests that pooled estimates should not be interpreted as fixed prevalence parameters. SSD prevalence was lower in older samples and studies using clinician-reported diagnoses or non-clinical recruitment, but higher in studies with greater methodological quality and clinical samples. Diagnostic method similarly moderated MDD prevalence, with higher prevalence observed in studies using the DDIS and lower prevalence in those using the SCID. The DDIS was also marginally associated with lower PTSD prevalence. The higher MDD prevalence observed in studies using the DDIS suggests that its use may inflate estimates when used without a more comprehensive assessment of differential diagnoses and consideration of symptom overlap. Differences in classification between diagnostic interviews have been previously observed for MDD (55). Thus, clinical diagnoses should only proceed following comprehensive assessment.

Greater female representation was associated with higher PTSD prevalence, consistent with established sex differences in PTSD (56). PTSD prevalence was found to be lower in Asian samples. Whilst this may reflect cultural, sampling, or diagnostic differences, the small number of Asian studies and potential influence of individual estimates mean that this finding should be interpreted cautiously. More broadly, the greater representation of MDD, BPD, SSD and SUD in DDIS-based studies suggests that diagnostic batteries may systematically influence which comorbidities are identified. Further research is warranted to identify the relevant differences between these instruments.

Heterogeneity for SUD and specific phobia decreased substantially following removal of influential studies, whereas heterogeneity remained high for PTSD and PDD, suggesting that persistent heterogeneity likely reflects additional methodological or population-level differences. Across the influence analyses, pooled prevalence estimates for several comorbidities increased following removal of influential or outlying studies, suggesting that the primary analyses may, on average, underestimate prevalence. Methodological sensitivity analyses also demonstrated that prevalence estimates were sensitive to methodological restrictions, particularly the approach used to assess comorbidity. Such changes illustrate that pooled estimates may not represent a single stable population parameter, particularly given the preponderance of low sample size studies.

The high prevalence of several comorbidities raises questions about diagnostic overlap and differential diagnosis. DID shares trauma-related symptomatology with PTSD, and substantial overlap with BPD, particularly in dissociation and emotion regulation (11, 49). Such overlap may contribute to apparent comorbidity. As previous psychiatric diagnoses and diagnostic trajectories were not systematically extracted, the present review cannot determine whether comorbidities reflected genuine co-occurrence, previous diagnostic formulations, diagnostic substitution, or symptom overlap.

The present findings have several clinical implications. Substantial psychiatric comorbidity should be anticipated when working with individuals with DID, and comprehensive assessment should be incorporated into routine clinical practice. In particular, the prevalence of borderline personality disorder and schizophrenia-spectrum diagnoses highlights the importance of careful differential diagnosis, given the potential for phenomenological overlap between dissociative, personality, and psychotic symptoms to contribute to misdiagnosis. At the same time, the high prevalence of depressive and anxiety disorders, and of bulimia nervosa, indicates a substantial broader symptom burden that may require additional clinical attention. The particularly high prevalence of PTSD, MDD, and BPD suggests that formulation and intervention may need to address several domains of psychopathology alongside dissociative symptoms, rather than focusing exclusively on DID-related phenomena. Conversely, substance use disorders were generally less prevalent, although they remained clinically relevant for a proportion of individuals. Diagnostic formulation should therefore consider both comorbidity and phenomenological overlap, rather than attributing all presenting difficulties to DID or relying on a single assessment method where diagnostic complexity is present. Identifying comorbidities may inform formulation and treatment planning by clarifying the range of difficulties requiring intervention and supporting consideration of appropriate treatment targets and priorities (57). However, the present findings do not establish treatment effectiveness or optimal sequencing.

### Strengths and Limitations

To our knowledge, this is the first meta-analysis to quantify psychiatric comorbidity across multiple diagnostic categories in DID. Meta-analysis enabled synthesis of prevalence estimates, whilst moderator, sensitivity and influence analyses provided information about sources of variance in estimates and an assessment of their robustness to methodological differences and influential studies. These findings have direct clinical relevance given the substantial burden of comorbidity identified.

Several limitations should be considered. Statistical uncertainty was evident for several outcomes, with small numbers of contributing studies with mostly small sample sizes, wide confidence intervals and substantial heterogeneity, reflected in the low or very low GRADE ratings for some estimates. Variation in diagnostic instruments also introduced measurement uncertainty. Generalisability was limited by predominantly clinical or treatment-seeking samples and poor reporting of ethnicity, restricting assessment of representativeness and cultural differences.

A small number of potentially relevant papers (*n* = 5) could not be retrieved, potentially reducing the comprehensiveness of the evidence base. Multiple publications from the same research groups also raised the possibility of overlapping samples that could not always be established, meaning that some correlated samples may have been treated as independent, possibly skewing estimates. Finally, previous psychiatric diagnoses and diagnostic trajectories were not systematically extracted because these were not reliably reported; such information would help to characterise patterns of misdiagnosis and diagnostic delay in DID (5–7). Consequently, the review cannot determine the extent to which observed comorbidity reflects genuine co-occurrence, previous diagnostic formulations, diagnostic substitution or symptom overlap, nor whether the timing of DID diagnosis or previous treatment influenced the observed prevalence estimates.

### Future Directions

Future research should prioritise standardised diagnostic assessment, with clearly defined, validated and consistently applied methods for both DID and psychiatric comorbidities. Larger samples are particularly needed for less frequently studied comorbidities, alongside greater representation of diverse populations to establish whether findings from highly specialised clinical populations generalise to the wider DID population. Further research should also prioritise comorbidities for which current prevalence estimates are characterised by wide confidence intervals or few contributing studies, as these estimates remain particularly uncertain. Prospective and longitudinal studies are needed to examine diagnostic trajectories and temporal relationships between DID and comorbid psychopathology. Improved reporting of sampling frames, recruitment, diagnostic procedures and participant characteristics would improve interpretability and comparability of future studies. Finally, the geographical differences identified warrant well-designed cross-national research to determine whether cultural or diagnostic contexts contribute to variation.

### Conclusion

The current results cumulatively highlight that DID is associated with a substantial and broad burden of psychiatric comorbidity, particularly across trauma-related, depressive, anxiety, and personality disorders. This broad comorbidity profile is important for understanding the overall symptom burden in DID, interpreting diagnostic presentations in the context of potential misdiagnosis and phenomenological overlap, and informing clinical assessment and treatment. However, considerable heterogeneity and uncertainty in prevalence estimates highlight the need for standardised diagnostic assessment and representative sampling to better establish the extent and pattern of comorbidity. Improved recognition of comorbid conditions may support more comprehensive formulation and inform treatment pathways tailored to this population, with increased consideration of co-occurring difficulties alongside dissociative symptoms.

## Acknowledgements

Artificial intelligence (AI) use: The authors made use of ChatGPT (GPT-5.6 Luna) to assist with the editing and preparation of this article. The tool was accessed via https://chatgpt.com/ and used without modification between 25.04.26 – 08.09.26. All AI-generated suggestions were reviewed and edited by E.M. The authors retain responsibility for the final content of the article.

## Author Contribution

E.M. and D.B.T. conceived the idea behind the current study. A.W. and S.P. critically reviewed the content and structure of the article. E.M. and A.K. acted as independent raters/extractors at each stage of the review. D.B.T. acted as a third reviewer. D.B.T., A.W., and S.P. supervised the project and critically reviewed and edited drafts until the final version. E.M. performed the meta-analysis and worked on all aspects of the paper. All authors contributed to the final manuscript.

## Transparency Declaration

The current manuscript is an honest, accurate and transparent account of the study being reported; no important aspects of the study have been omitted.

## Data Availability

The data that support the findings of this study are available in the original papers included in the meta-analysis.

## Relevance Statement

The present review is relevant to practising psychiatrists because it highlights the substantial and broad psychiatric comorbidity associated with dissociative identity disorder (DID). The findings support comprehensive assessment of comorbid psychopathology in individuals with DID, while highlighting the potential influence of diagnostic methods and phenomenological overlap on prevalence estimates. Awareness of these factors supports more accurate diagnostic formulation and will inform clinical assessment and treatment planning.

