## Supplementary Materials for "Psychiatric comorbidity in dissociative identity disorder: A meta-analysis"

16 De Crespigny Park

London, UK, SE5 8AB

**Supplementary Table 1.**

*Preferred Reporting Items for Systematic Reviews and Meta-Analyses (PRISMA) Checklist.*

| **Section and Topic** | **Item #** | **Checklist item** | **Location where item is reported** |
| --- | --- | --- | --- |
| **TITLE** | | |  |
| Title | 1 | Identify the report as a systematic review. | Title page (meta-analysis) |
| **ABSTRACT** | | |  |
| Abstract | 2 | See the PRISMA 2020 for Abstracts checklist. | Abstract |
| **INTRODUCTION** | | |  |
| Rationale | 3 | Describe the rationale for the review in the context of existing knowledge. | Introduction |
| Objectives | 4 | Provide an explicit statement of the objective(s) or question(s) the review addresses. | Introduction |
| **METHODS** | | |  |
| Eligibility criteria | 5 | Specify the inclusion and exclusion criteria for the review and how studies were grouped for the syntheses. | Methods, Supplementary Table 2 |
| Information sources | 6 | Specify all databases, registers, websites, organisations, reference lists and other sources searched or consulted to identify studies. Specify the date when each source was last searched or consulted. | Methods |
| Search strategy | 7 | Present the full search strategies for all databases, registers and websites, including any filters and limits used. | Supplementary Materials |
| Selection process | 8 | Specify the methods used to decide whether a study met the inclusion criteria of the review, including how many reviewers screened each record and each report retrieved, whether they worked independently, and if applicable, details of automation tools used in the process. | Methods |
| Data collection process | 9 | Specify the methods used to collect data from reports, including how many reviewers collected data from each report, whether they worked independently, any processes for obtaining or confirming data from study investigators, and if applicable, details of automation tools used in the process. | Methods |
| Data items | 10a | List and define all outcomes for which data were sought. Specify whether all results that were compatible with each outcome domain in each study were sought (e.g. for all measures, time points, analyses), and if not, the methods used to decide which results to collect. | Methods |
|  | 10b | List and define all other variables for which data were sought (e.g. participant and intervention characteristics, funding sources). Describe any assumptions made about any missing or unclear information. | Methods |
| Study risk of bias assessment | 11 | Specify the methods used to assess risk of bias in the included studies, including details of the tool(s) used, how many reviewers assessed each study and whether they worked independently, and if applicable, details of automation tools used in the process. | Methods |
| Effect measures | 12 | Specify for each outcome the effect measure(s) (e.g. risk ratio, mean difference) used in the synthesis or presentation of results. | Methods |
| Synthesis methods | 13a | Describe the processes used to decide which studies were eligible for each synthesis (e.g. tabulating the study intervention characteristics and comparing against the planned groups for each synthesis (item #5)). | Methods |
|  | 13b | Describe any methods required to prepare the data for presentation or synthesis, such as handling of missing summary statistics, or data conversions. | Methods |
|  | 13c | Describe any methods used to tabulate or visually display results of individual studies and syntheses. | Methods |
|  | 13d | Describe any methods used to synthesize results and provide a rationale for the choice(s). If meta-analysis was performed, describe the model(s), method(s) to identify the presence and extent of statistical heterogeneity, and software package(s) used. | Methods |
|  | 13e | Describe any methods used to explore possible causes of heterogeneity among study results (e.g. subgroup analysis, meta-regression). | Methods |
|  | 13f | Describe any sensitivity analyses conducted to assess robustness of the synthesized results. | Methods |
| Reporting bias assessment | 14 | Describe any methods used to assess risk of bias due to missing results in a synthesis (arising from reporting biases). | Methods |
| Certainty assessment | 15 | Describe any methods used to assess certainty (or confidence) in the body of evidence for an outcome. | Methods |
| **RESULTS** | | |  |
| Study selection | 16a | Describe the results of the search and selection process, from the number of records identified in the search to the number of studies included in the review, ideally using a flow diagram. | Figure 1 |
|  | 16b | Cite studies that might appear to meet the inclusion criteria, but which were excluded, and explain why they were excluded. | - |
| Study characteristics | 17 | Cite each included study and present its characteristics. | Results, Table 1 |
| Risk of bias in studies | 18 | Present assessments of risk of bias for each included study. | Results, Supplementary Table 3 |
| Results of individual studies | 19 | For all outcomes, present, for each study: (a) summary statistics for each group (where appropriate) and (b) an effect estimate and its precision (e.g. confidence/credible interval), ideally using structured tables or plots. | Results, Table 2, Supplementary Figures 1-23, and 24-46 |
| Results of syntheses | 20a | For each synthesis, briefly summarise the characteristics and risk of bias among contributing studies. | Results |
|  | 20b | Present results of all statistical syntheses conducted. If meta-analysis was done, present for each the summary estimate and its precision (e.g. confidence/credible interval) and measures of statistical heterogeneity. If comparing groups, describe the direction of the effect. | Results, Table 2 |
|  | 20c | Present results of all investigations of possible causes of heterogeneity among study results. | Results, Table 3, Supplementary Tables 5, 6, and 7 |
|  | 20d | Present results of all sensitivity analyses conducted to assess the robustness of the synthesized results. | Results, Supplementary Tables 6 and 7 |
| Reporting biases | 21 | Present assessments of risk of bias due to missing results (arising from reporting biases) for each synthesis assessed. | - |
| Certainty of evidence | 22 | Present assessments of certainty (or confidence) in the body of evidence for each outcome assessed. | Results, Supplementary Table 8 |
| **DISCUSSION** | | |  |
| Discussion | 23a | Provide a general interpretation of the results in the context of other evidence. | Discussion |
|  | 23b | Discuss any limitations of the evidence included in the review. | Discussion |
|  | 23c | Discuss any limitations of the review processes used. | Discussion |
|  | 23d | Discuss implications of the results for practice, policy, and future research. | Discussion |
| **OTHER INFORMATION** | | |  |
| Registration and protocol | 24a | Provide registration information for the review, including register name and registration number, or state that the review was not registered. | Methods |
|  | 24b | Indicate where the review protocol can be accessed, or state that a protocol was not prepared. | Methods |
|  | 24c | Describe and explain any amendments to information provided at registration or in the protocol. | Transparency Declaration |
| Support | 25 | Describe sources of financial or non-financial support for the review, and the role of the funders or sponsors in the review. | Funding |
| Competing interests | 26 | Declare any competing interests of review authors. | Declaration of interest |
| Availability of data, code and other materials | 27 | Report which of the following are publicly available and where they can be found: template data collection forms; data extracted from included studies; data used for all analyses; analytic code; any other materials used in the review. | Data Availability, Analytic Code Availability, Research Material Availability |

**Search terms**

The following search strategy, based on a PubMed search via NLM was used and adapted based on database syntax rules and default search behaviours:

("Dissociative Identity Disorder" OR DID OR "Multiple Personality Disorder") AND (Comorbid* OR co-morbid* OR coexist* OR co-exist* OR multimorbid* OR multi-morbid* OR pre-exist OR preexist* OR "intellectual disabilit*" OR "mental retardation" OR "developmental delay" OR "global developmental delay" OR autism* OR ASD OR ASC OR “infantile autism” OR "pervasive developmental" OR "asperger*" OR ADHD OR "attention deficit hyperactivity disorder" OR "attachment disorder" OR "specific learning disorder*" OR "communication disorder*" OR "developmental coordination disorder*" OR stuttering OR "language disorder*" OR schizophrenia* OR SSD OR "schizophreniform disorder*" OR "schizoaffective disorder*" OR "brief psychotic disorder*" OR "delusional disorder*" OR "shared psychotic disorder*" OR "psychotic disorder*" OR psychosis OR bipolar OR "bipolar I disorder" OR "bipolar II disorder" OR "bipolar affective disorder" OR BPAD OR mania OR hypomania OR cyclothymi* OR "manic depress*" OR "major depress*" OR MDD OR dysthymi* OR "persistent depressive disorder*" OR "panic disorder*" OR agoraphobia OR "social phobia" OR "social anxiety disorder" OR SAD OR "specific phobia" OR "generalized anxiety disorder" OR "generalised anxiety disorder" OR GAD OR "obsessive-compulsive disorder*" OR OCD OR "body dysmorphic disorder*" OR BDD OR "hoarding disorder*" OR PTSD OR "post-traumatic stress disorder*" OR "acute stress disorder*" OR "adjustment disorder*" OR "somatization disorder*" OR "somatisation disorder*"OR "somatic symptom disorder*" OR "somatoform disorder" OR "bodily distress disorder" OR "conversion disorder*" OR "hypochondria*" OR "psychogenic pain disorder*" OR "depersonalization disorder*" OR "depersonalisation disorder*" OR "derealization disorder*" OR "derealisation disorder*" OR "possession trance disorder" OR "trance disorder" OR "anorexia nervosa" OR "bulimia nervosa" OR "binge eating disorder*" OR "avoidant restrictive food intake disorder*" OR ARFID OR pica OR enuresis* OR encopresis* OR "insomnia disorder*" OR hypersomnia OR narcolepsy OR "sleep apnoea" OR "sleep apnea" OR parasomnia* OR "sleep terror*" OR "sleepwalking disorder*" OR "nightmare disorder" OR "sexual dysfunction*" OR paraphilia* OR exhibitionism OR fetishism OR frotteurism OR pedophilia OR paedophilia OR "sexual masochism" OR "sexual sadism" OR "transvestic fetishism" OR "voyeurism" OR "gender dysphoria*" OR "gender identity disorder*" OR "substance use disorder*" OR “substance use” OR SUD OR “drug dependence syndrome” OR "alcohol use disorder*" OR AUD OR "alcohol dependence” OR "opioid use disorder*" OR OUD OR "cannabis use disorder*" OR CUD OR "stimulant use disorder*" OR "sedative use disorder*" OR "hallucinogen use disorder*" OR "inhalant use disorder*" OR "gambling disorder*" OR "personality disorder*" OR PD OR "borderline personality" OR "emotionally unstable personality" OR EUPD OR "antisocial personality" OR "narcissistic personality" OR "avoidant personality" OR "dependent personality" OR "asthenic disorder" OR "obsessive-compulsive personality" OR OCPD OR "passive-aggressive personality" OR "self-defeating personality" OR "histrionic personality" OR "schizo* personality" OR "paranoid personality" OR "intermittent explosive disorder*" OR kleptomania* OR pyromania* OR trichotillomania* OR "excoriation disorder" OR "conduct disorder*" OR "conduct dissocial disorder" OR CDD OR "oppositional defiant disorder*" OR ODD OR "factitious disorder" OR malingering OR "Munchausen by proxy" OR "tic disorder" OR "Tourette syndrome" OR "Tourette disorder" OR "mental disorder*" OR "mental illness*" OR "psychiatric disorder*" OR "psychiatric illness*" OR psychopathology)

| **Supplementary Table 2**  *Selection criteria for study inclusion and exclusion.* | | |
| --- | --- | --- |
| Criterion | Inclusion | Exclusion |
| Population | Adults with a diagnosis of DID or MPD according to DSM or ICD criteria (from DSM-III or ICD-10) identified by clinician report, psychiatric records, or administration of a structured, validated diagnostic instrument. | - Paediatric populations. - Self-report diagnosis of DID. - Combined data for participants with DID and other dissociative disorders, unless data are presented separately. - People with DID within another clinical population (e.g., individuals with schizophrenia). |
| Exposure (Condition of Interest) | One or more comorbid psychiatric disorders according to DSM or ICD criteria (from DSM-III or ICD-10) identified by clinician report, psychiatric records, or administration of a structured, validated diagnostic instrument. | - Self-report diagnosis of psychiatric comorbidities. - Studies reporting only broad categories combining multiple psychiatric disorders (e.g., anxiety disorders). - Psychiatric disorders reported as ‘previous diagnoses’. |
| Comparator | No comparator is required for inclusion in this review. |  |
| Outcome | Prevalence estimates or counts of individual psychiatric comorbidities with DID. | Comorbidity not reported in ≥3 independent studies. |
| Study Type | - Observational, epidemiological, clinical, and experimental studies (including cross-sectional, cohort, case-control, and case series designs). - Peer-reviewed articles published from 1980 onwards. - English-language studies. - Sample size ≥ 10. | - Opinions articles, reviews. - Non-consecutively recruited case series. - Unpublished sources (e.g., conference abstracts and doctoral theses). |

| **Supplementary Table 3.**  *References of included studies.* | |
| --- | --- |
| Study | Reference |
| Abdellah et al. 2025 | Abdellah MM, Ragheb KM, Elshafie TM, Hamed RA. Prevalence of dissociative identity disorder among psychiatric outpatients in different cultural groups. Middle East Current Psychiatry. 2025;32(1):54. |
| Boon & Draijer 1993 | Boon S, Draijer N. The differentiation of patients with MPD or DDNOS from patients with a cluster B personality disorder. Dissociation. 1993;6(2-3):126-35. |
| Brand et al. 2006, 2009 | Brand BL, Armstrong JG, Loewenstein RJ. Psychological Assessment of Patients with Dissociative Identity Disorder. Psychiatric Clinics. 2006;29(1):145-68. |
| Brand et al. 2006, 2009 | Brand BL, Armstrong JG, Loewenstein RJ, McNary SW. Personality differences on the Rorschach of dissociative identity disorder, borderline personality disorder, and psychotic inpatients. Educational Publishing Foundation; 2009. p. 188-205. |
| Brand et al. 2021 | Brand BL, Barth M, Schlumpf YR, Schielke H, Chalavi S, Vissia EM, et al. The utility of the Structured Inventory of Malingered Symptomatology for distinguishing individuals with Dissociative Identity Disorder (DID) from DID simulators and healthy controls. European Journal of Psychotraumatology. 2021;12(1):1984048. |
| Chalavi et al. 2015 | Chalavi S, Vissia EM, Giesen ME, Nijenhuis ERS, Draijer N, Cole JH, et al. Abnormal hippocampal morphology in dissociative identity disorder and post-traumatic stress disorder correlates with childhood trauma and dissociative symptoms. Human Brain Mapping. 2015;36(5):1692-704. |
| Coons et al. 1991 | Coons PM, Bowman ES, Kluft RP, Milstein V. The cross-cultural occurrence of MPD: Additional cases from a recent survey. Dissociation. 1991;4(3):124-8. |
| Dale et al. 2009 | Dale KY, Berg R, Elden Å, Ødegård A, Holte A. Testing the Diagnosis of Dissociative Identity Disorder Through Measures of Dissociation, Absorption, Hypnotizability and PTSD: A Norwegian Pilot Study. Journal of Trauma & Dissociation. 2009;10(1):102-12. |
| Ellason et al. 1996 | Ellason JW, Ross CA, Fuchs DL. Lifetime Axis I and II Comorbidity and Childhood Trauma History in Dissociative Identity Disorder. Psychiatry. 1996;59(3):255-66. |
| Fink & Golinkoff 1990 | Fink DL, Golinkoff M. MPD, borderline personality disorder and schizophrenia: A comparative study of clinical features. 1990;3:127-34. |
| Lauer et al. 1993 | Lauer J, Black DW, Keen P. Multiple Personality Disorder and Borderline Personality Disorder Distinct Entities or Variations On a Common Theme? Annals of Clinical Psychiatry. 1993;5(2):129-34. |
| Manchester et al. 1998 | Manchester CF, Allen T, Tachiki KH. Treatment of dissociative identity disorder with neurotherapy and group self-exploration. Journal of Neurotherapy. 1998;2(4):40-53. |
| Merker et al. 2021 | Merker JB, Hill SB, Wolff JD, Winternitz SR, Ressler KJ, Kaufman ML, et al. Posttraumatic cognitions predict distorted body perceptions in women with dissociative identity disorder. Journal of Psychiatric Research. 2021;134:166-72. |
| Öztürk & Şar 2008 | Öztürk E, Sar V. Somatization as a predictor of suicidal ideation in dissociative disorders. Psychiatry and Clinical Neurosciences. 2008;62(6):662-8. |
| Reinders et al. 2018 | Reinders AATS, Chalavi S, Schlumpf YR, Vissia EM, Nijenhuis ERS, Jäncke L, et al. Neurodevelopmental origins of abnormal cortical morphology in dissociative identity disorder. Acta Psychiatrica Scandinavica. 2018;137(2):157-70. |
| Rodewald et al. 2011 | Rodewald F, Wilhelm-Göling C, Emrich H, Reddemann L, Gast U. Axis-I Comorbidity in Female Patients With Dissociative Identity Disorder and Dissociative Identity Disorder Not Otherwise Specified. Journal of Nervous & Mental Disease. 2011;199(2):122-31. |
| Ross et al. 1989 | Ross CA, Heber S, Anderson G, Norton GR, Anderson BA, del Campo M, et al. Differentiating multiple personality disorder and complex partial seizures. General Hospital Psychiatry. 1989;11(1):54-8. |
| Ross et al. 1990 | Ross CA, Fast E, Anderson G, Auty A, Todd J. Somatic symptoms in multiple sclerosis and MPD. Dissociation. 1990;3(2):102-6. |
| Ross et al. 1992 | Ross CA, Anderson G, Fraser GA, Reagor P, et al. Differentiating multiple personality disorder and dissociative disorder not otherwise specified. Ridgeview Inst; 1992. p. 87-90. |
| Ross et al. 2014 | Ross CA, Ferrell L, Schroeder E. Co-Occurrence of Dissociative Identity Disorder and Borderline Personality Disorder. Journal of Trauma & Dissociation. 2014;15(1):79-90. |
| Şar et al. 2001 | Şar V, Unal SN, Kiziltan E, Kundakci T, Ozturk E. HMPAO SPECT Study of Regional Cerebral Blood Flow in Dissociative Identity Disorder. Journal of Trauma & Dissociation. 2001;2(2):5-25. |
| Şar et al. 2007 | Şar V, Unal SN, Ozturk E. Frontal and occipital perfusion changes in dissociative identity disorder. Psychiatry Research: Neuroimaging. 2007;156(3):217-23. |
| Vermetten et al. 2006 | Vermetten E, Schmahl C, Lindner S, Loewenstein Richard J, Bremner JD. Hippocampal and Amygdalar Volumes in Dissociative Identity Disorder. American Journal of Psychiatry. 2006;163:630-6. |
| Yargiç et al. 1998 | Yargiç L, Şar V, Tutkun H, Alyanak B. Comparison of dissociative identity disorder with other diagnostic groups using a structured interview in Turkey. Comprehensive Psychiatry. 1998;39(6):345-51. |

**Forest plots of meta-analysis results**

Supplementary Figures 1-23 present forest plots for all psychiatric comorbidities included in the meta-analyses. Each square marker (and whiskers) represents the individual study logit-transformed prevalence estimate (and 95% CI), with marker size reflecting study weight (larger size denotes greater influence). The bottom diamond represents the pooled prevalence estimate and its 95% CI, with the pooled estimate on the logit-transformed scale. Pooled estimates and 95% confidence intervals are reported as back-transformed prevalence values in the main text and tables for ease of interpretation.

**Supplementary Figure 1.**

*Forest plot of logit-transformed prevalence estimates (with 95% CIs) for posttraumatic stress disorder among individuals with DID.*

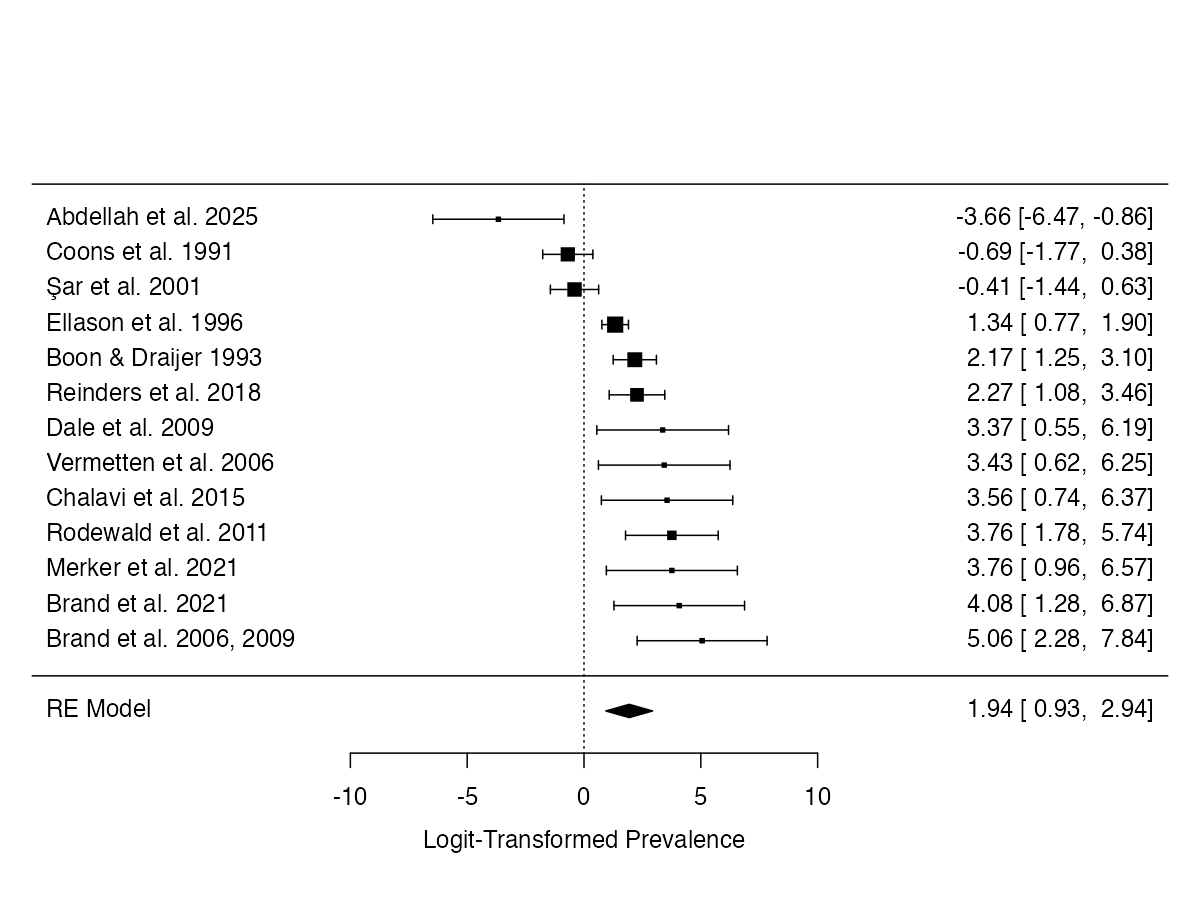

**Supplementary Figure 2.**

*Forest plot of logit-transformed prevalence estimates (with 95% CIs) for major depressive disorder among individuals with DID.*

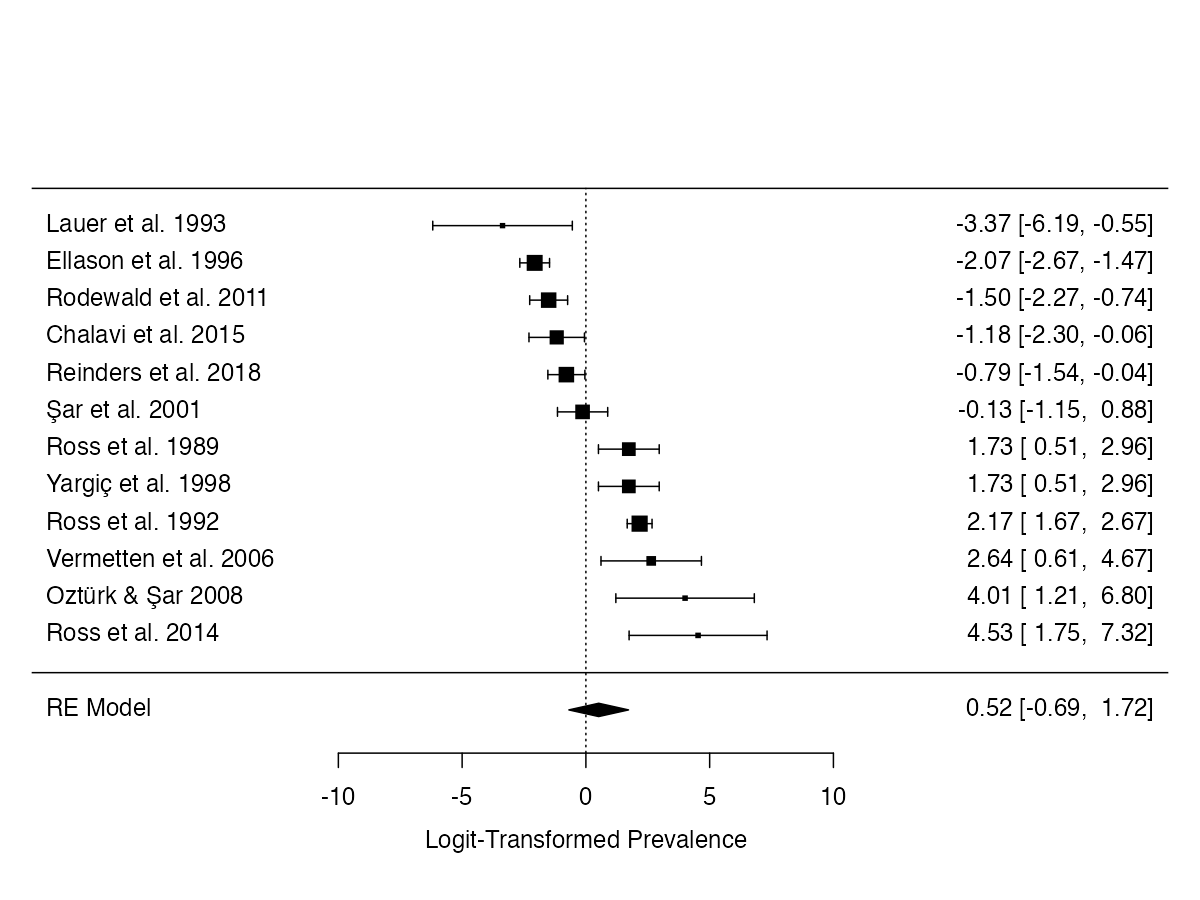

**Supplementary Figure 3.**

*Forest plot of logit-transformed prevalence estimates (with 95% CIs) for borderline personality disorder among individuals with DID.*

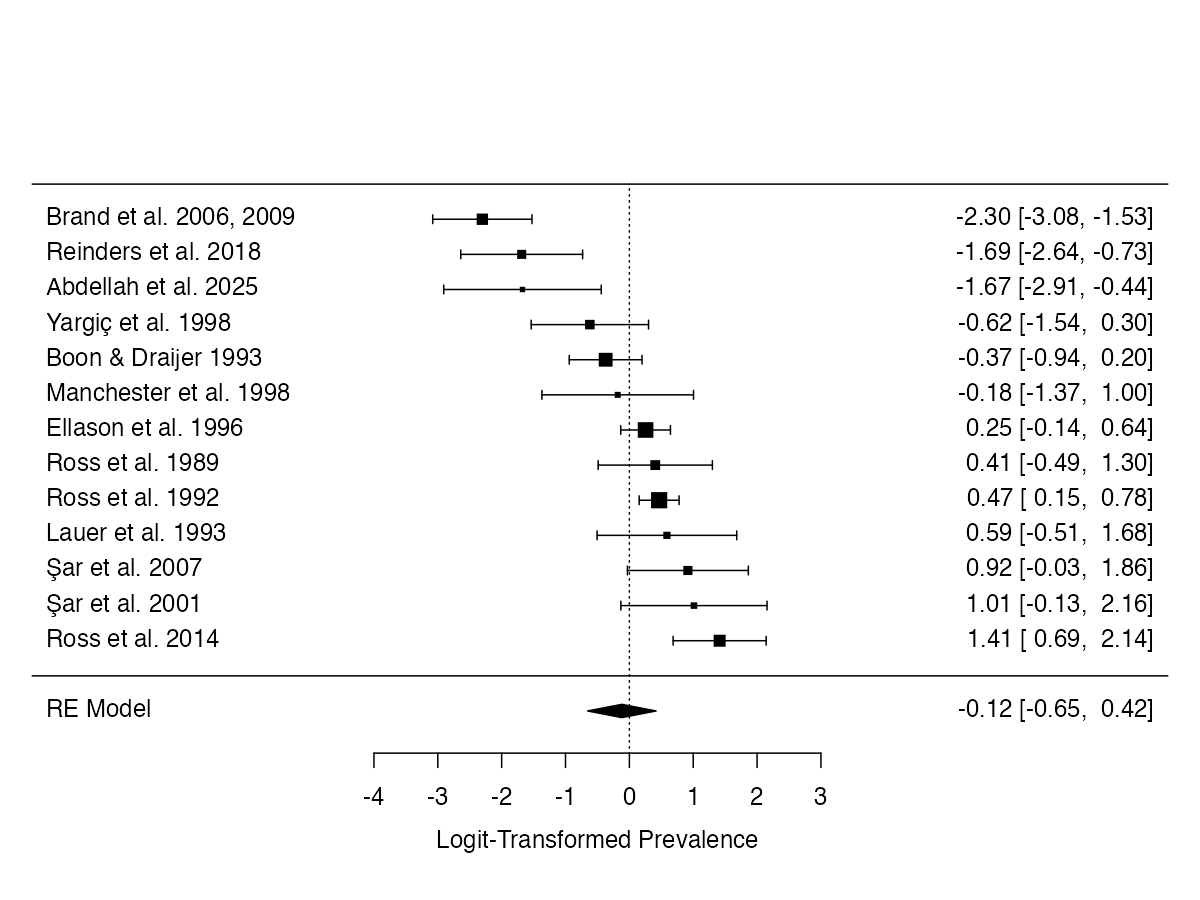

**Supplementary Figure 4.**

*Forest plot of logit-transformed prevalence estimates (with 95% CIs) for panic disorder among individuals with DID.*

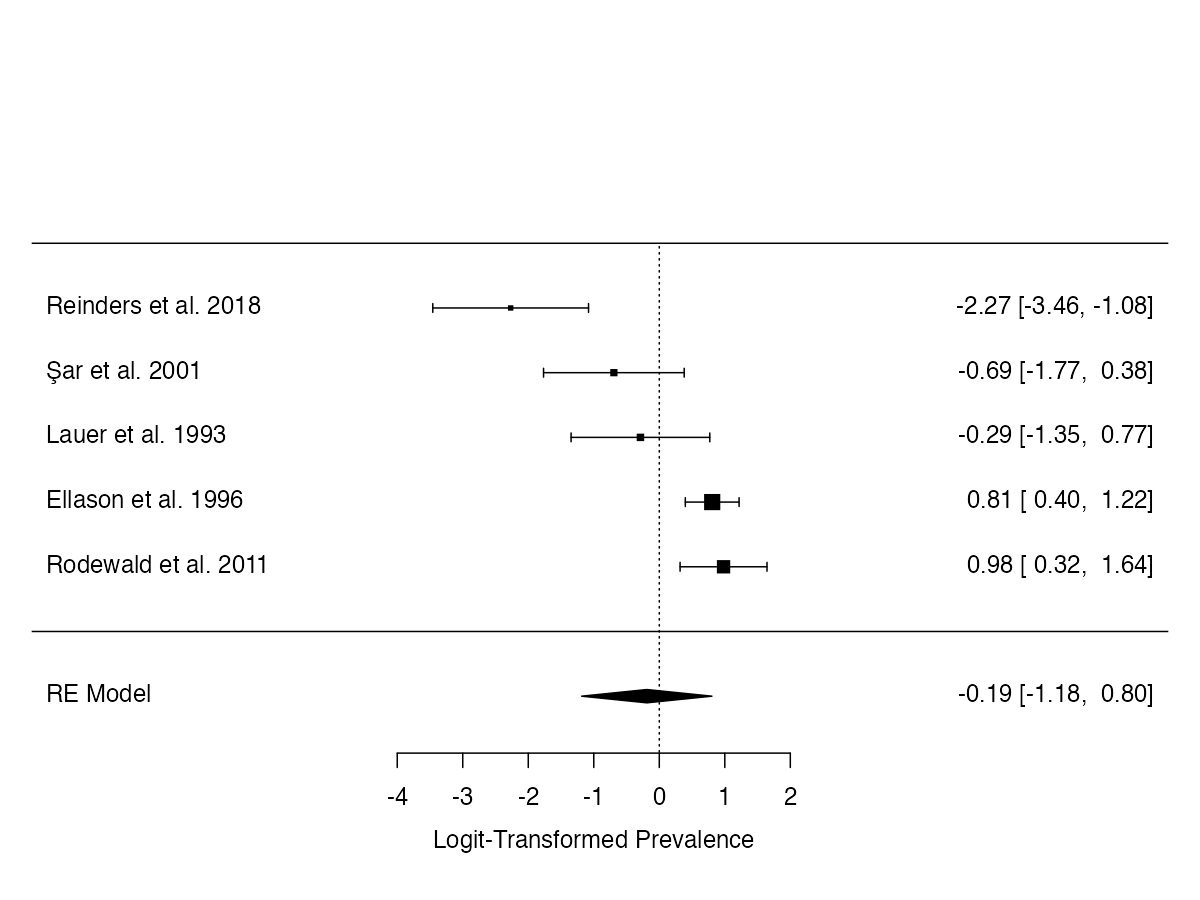

**Supplementary Figure 5.**

*Forest plot of logit-transformed prevalence estimates (with 95% CIs) for schizoaffective disorder among individuals with DID.*

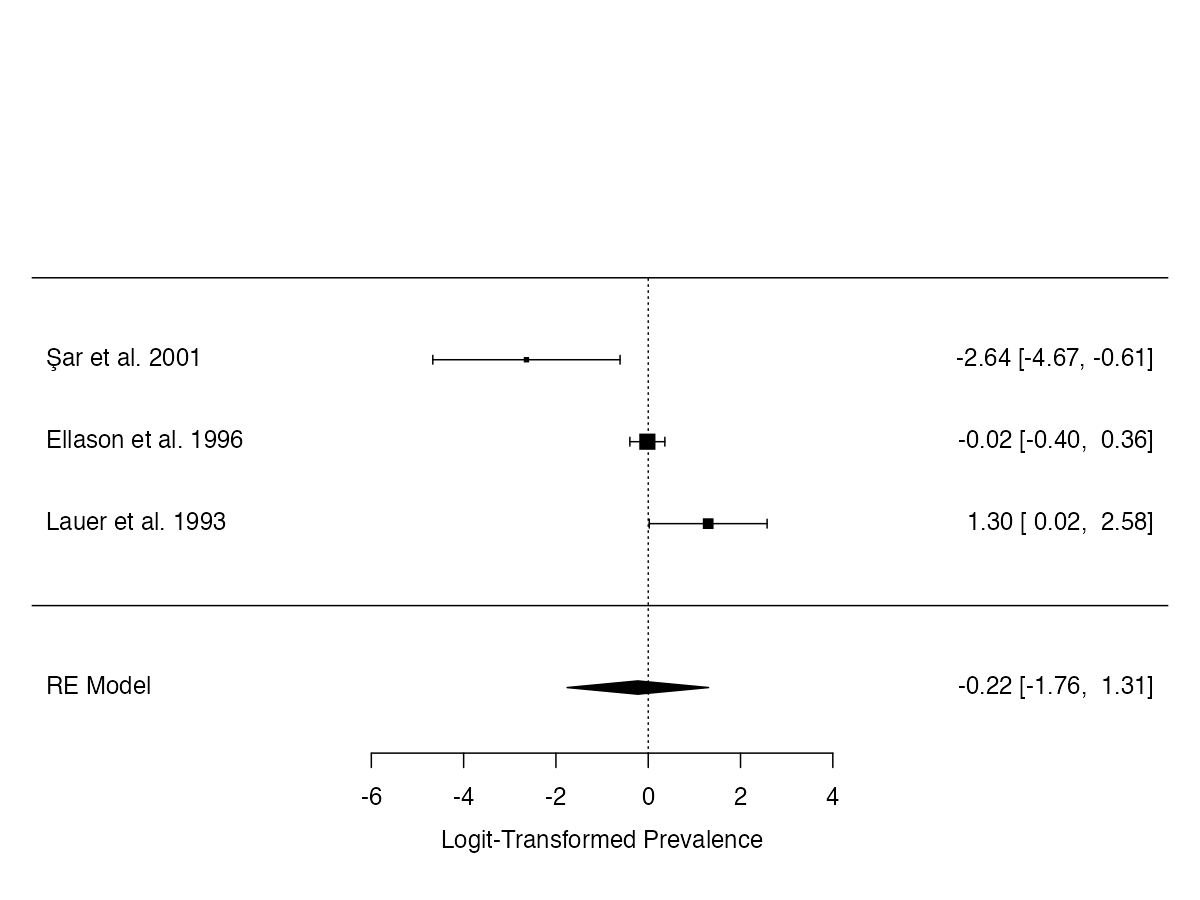

**Supplementary Figure 6.**

*Forest plot of logit-transformed prevalence estimates (with 95% CIs) for social anxiety disorder among individuals with DID.*

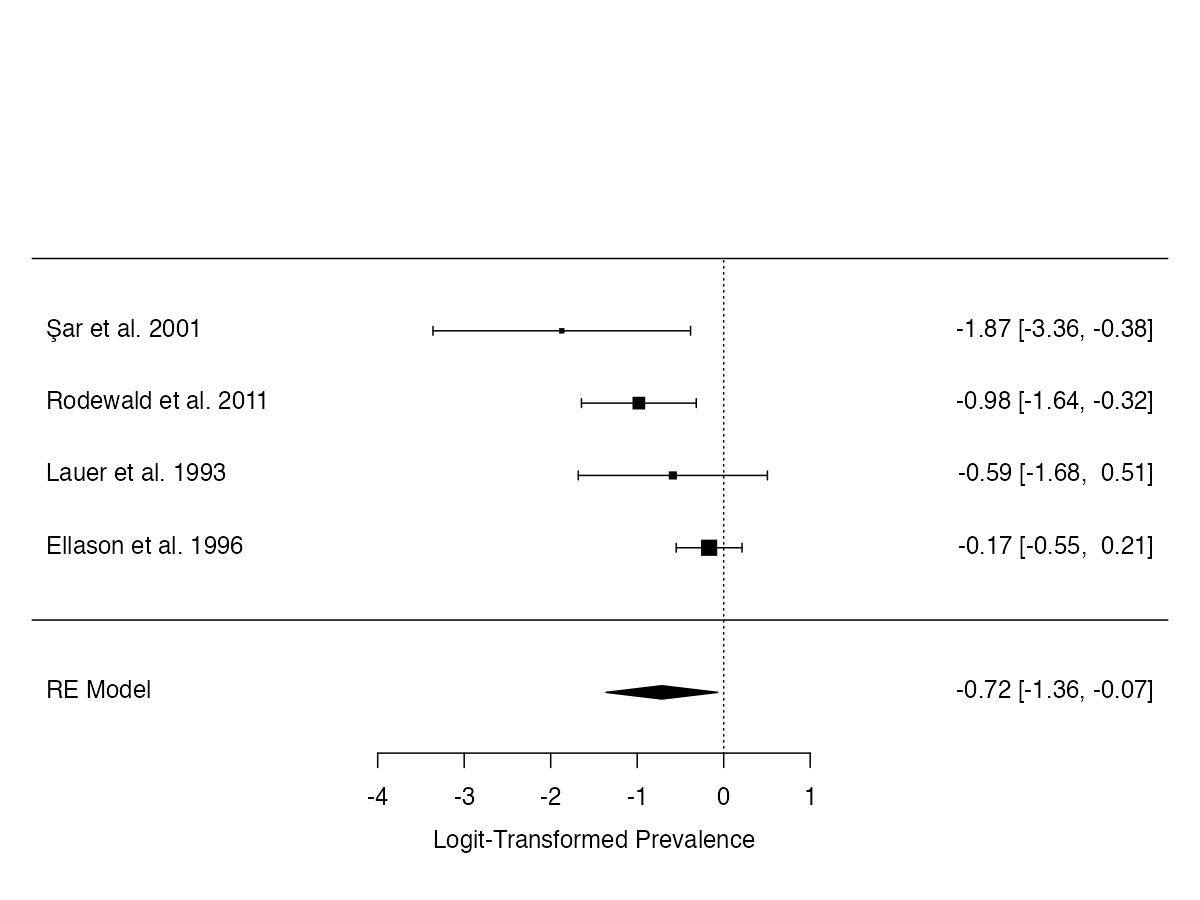

**Supplementary Figure 7.**

*Forest plot of logit-transformed prevalence estimates (with 95% CIs) for somatic symptom disorder among individuals with DID.*

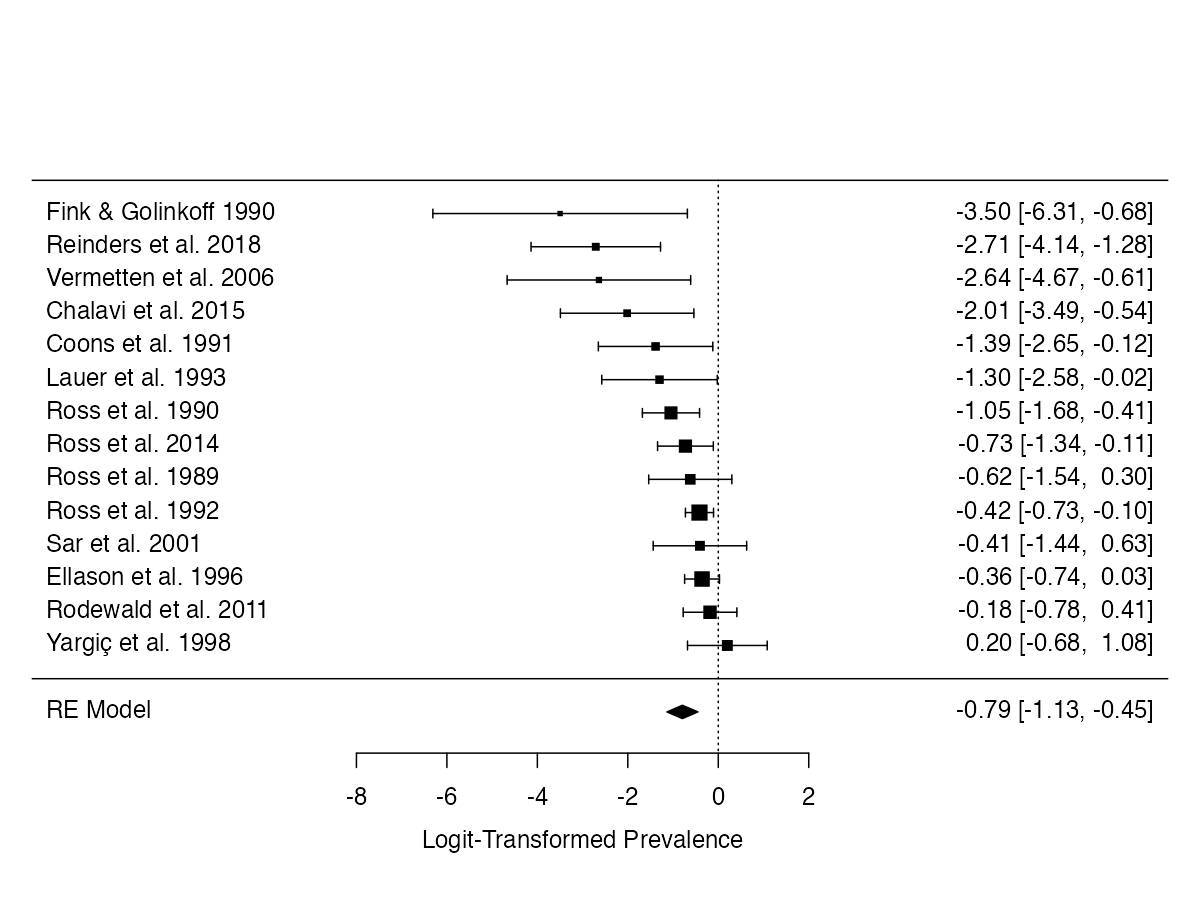

**Supplementary Figure 8.** Forest plot of logit-transformed prevalence estimates (with 95% CIs) for alcohol use disorder among individuals with DID.

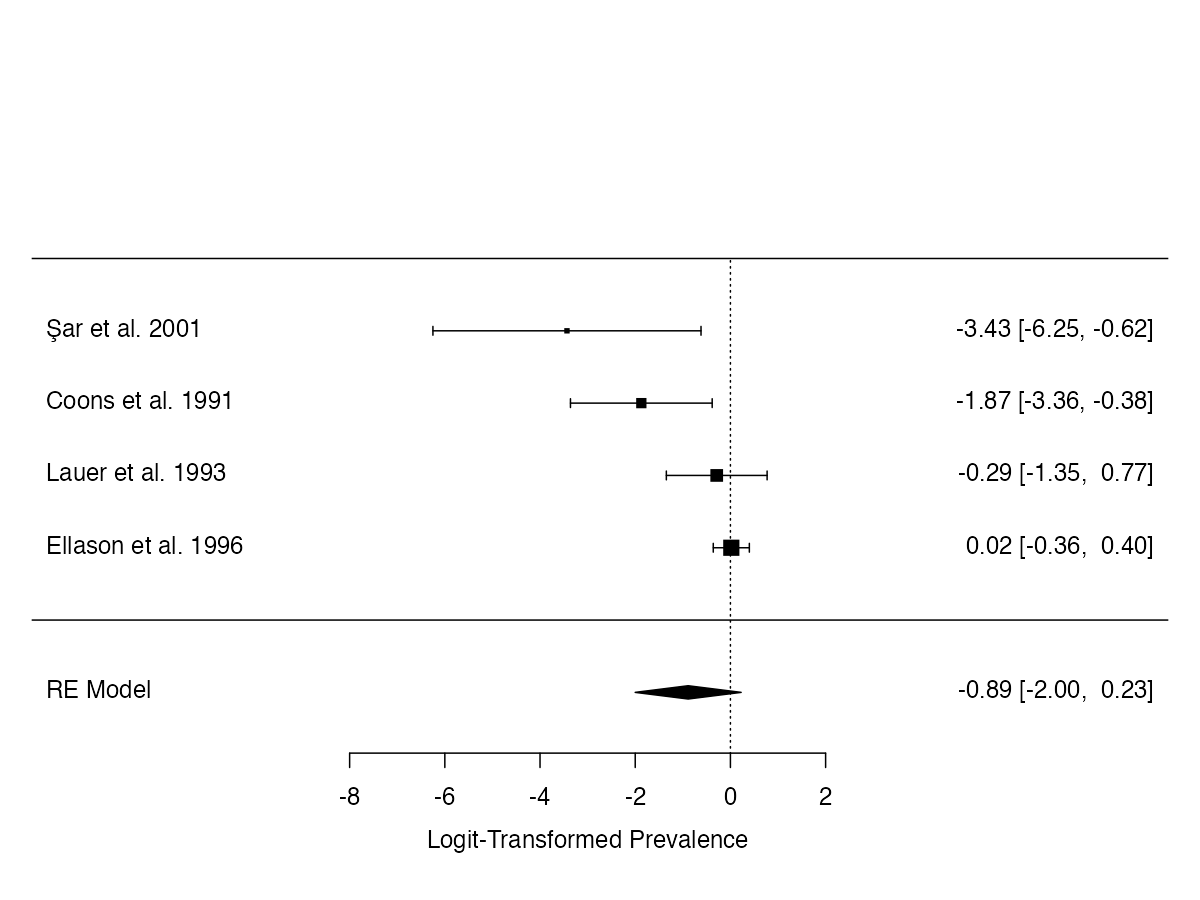

**Supplementary Figure 9.**

*Forest plot of logit-transformed prevalence estimates (with 95% CIs) for specific phobia among individuals with DID.*

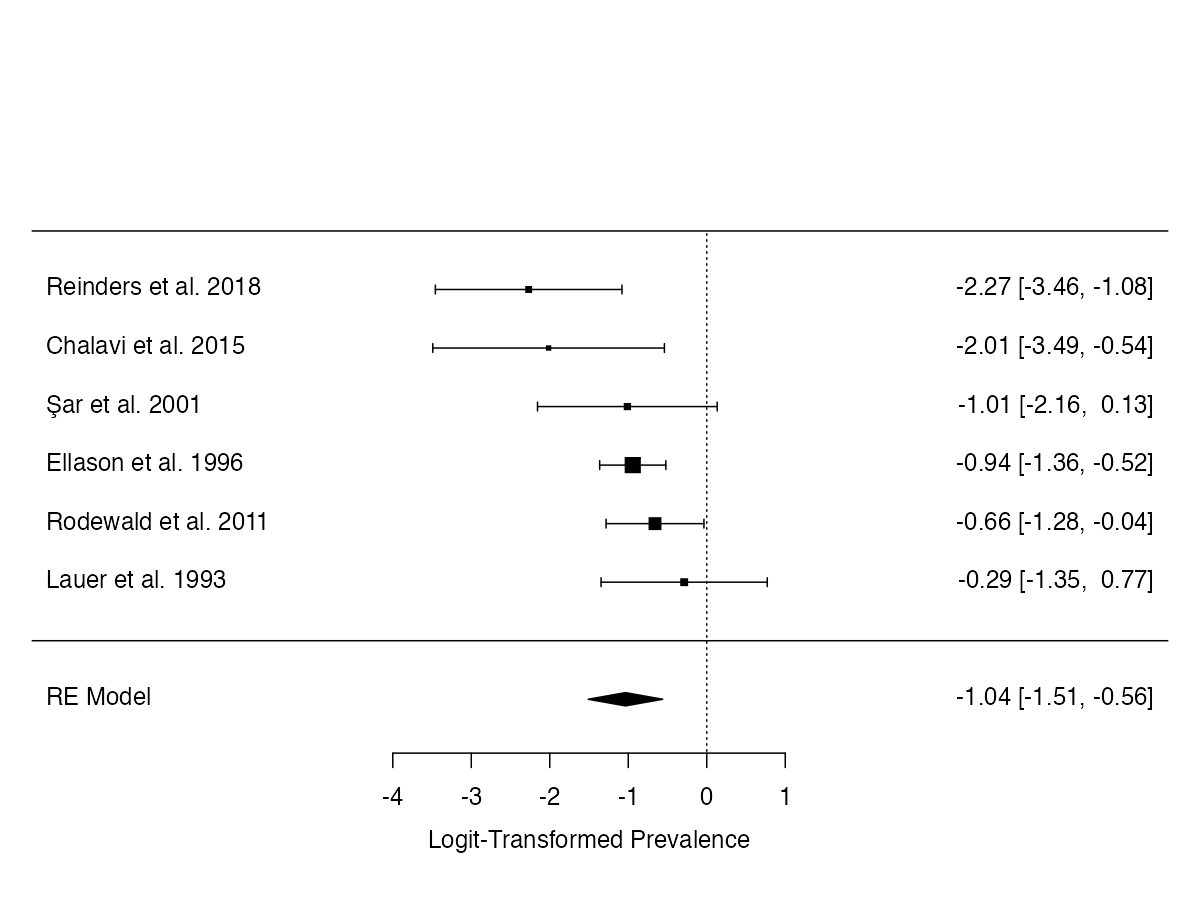

**Supplementary Figure 10.**

*Forest plot of logit-transformed prevalence estimates (with 95% CIs) for agoraphobia among individuals with DID.*

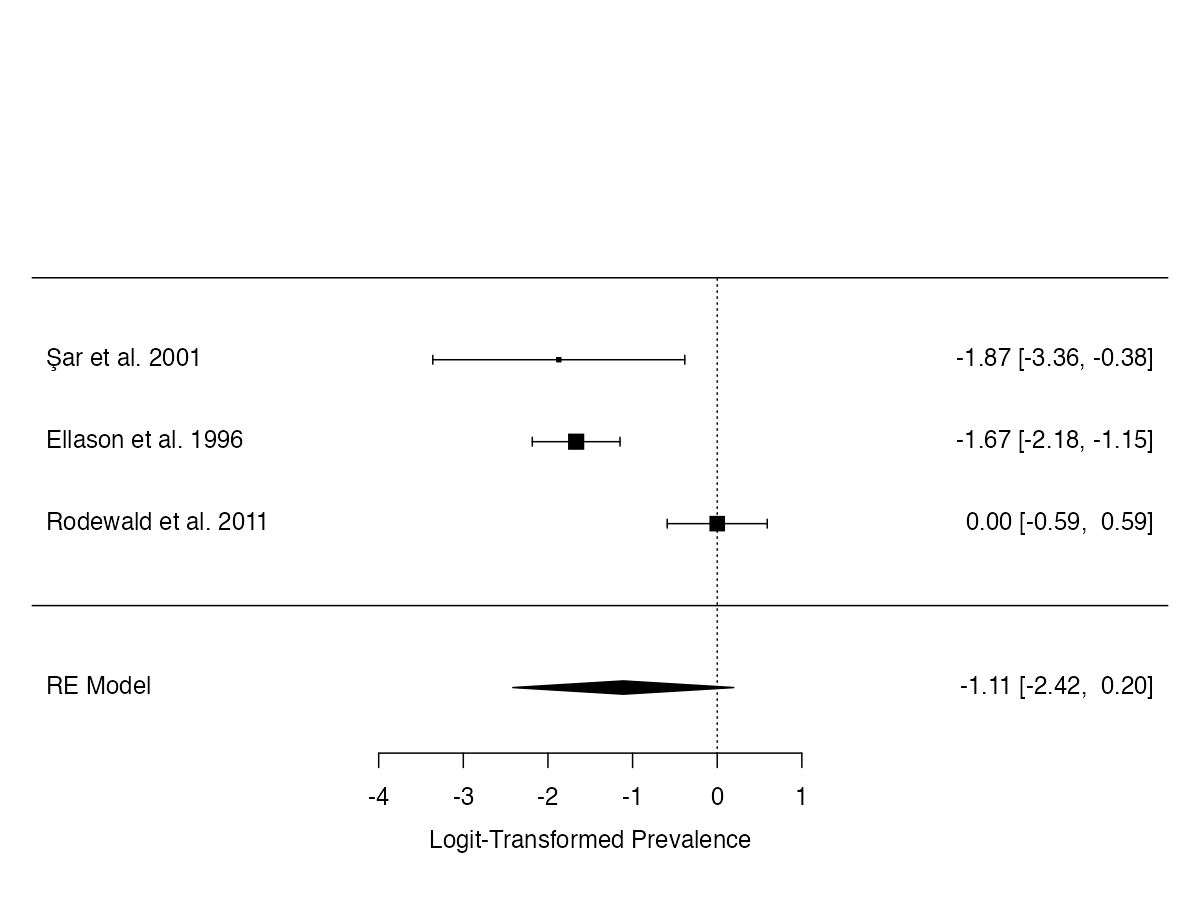

**Supplementary Figure 11.**

*Forest plot of logit-transformed prevalence estimates (with 95% CIs) for bulimia nervosa among individuals with DID.*

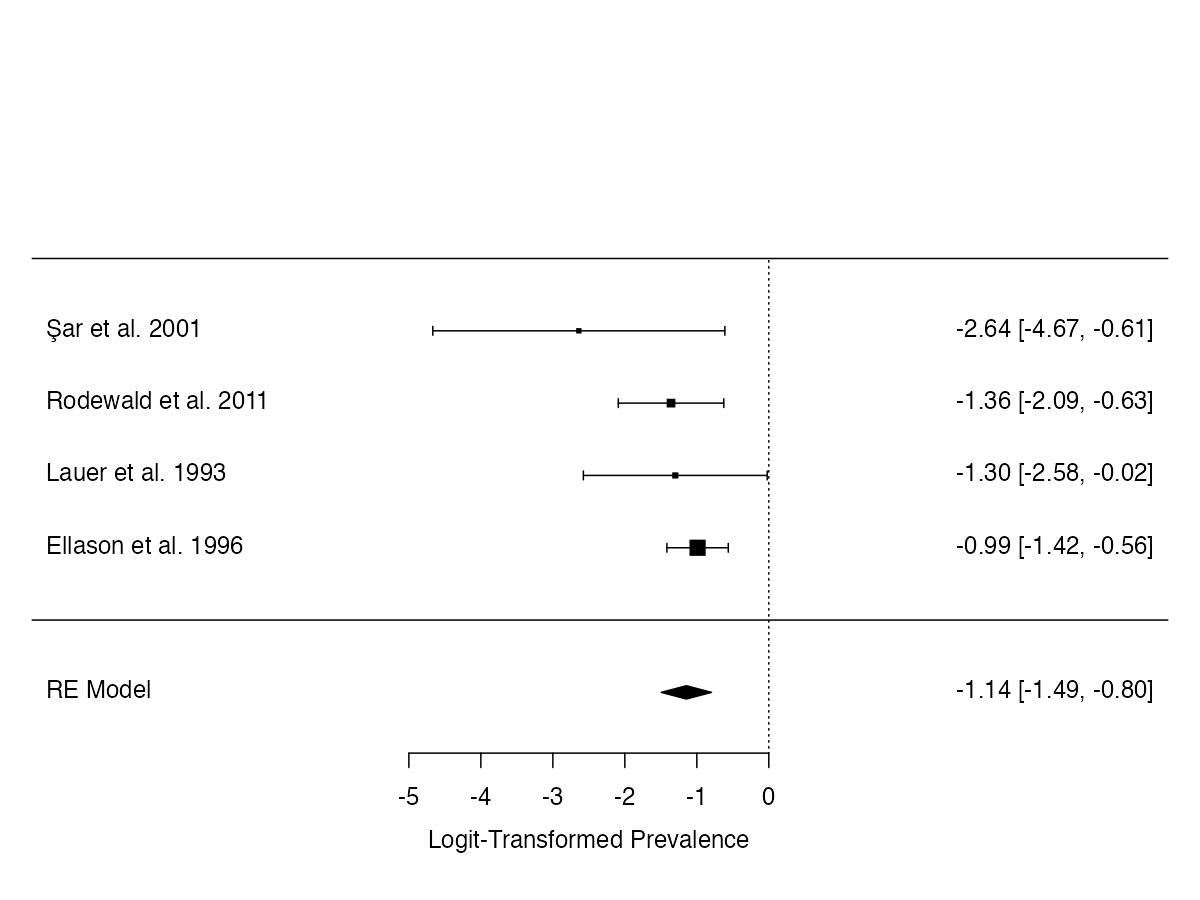

**Supplementary Figure 12.**

*Forest plot of logit-transformed prevalence estimates (with 95% CIs) for obsessive-compulsive disorder among individuals with DID.*

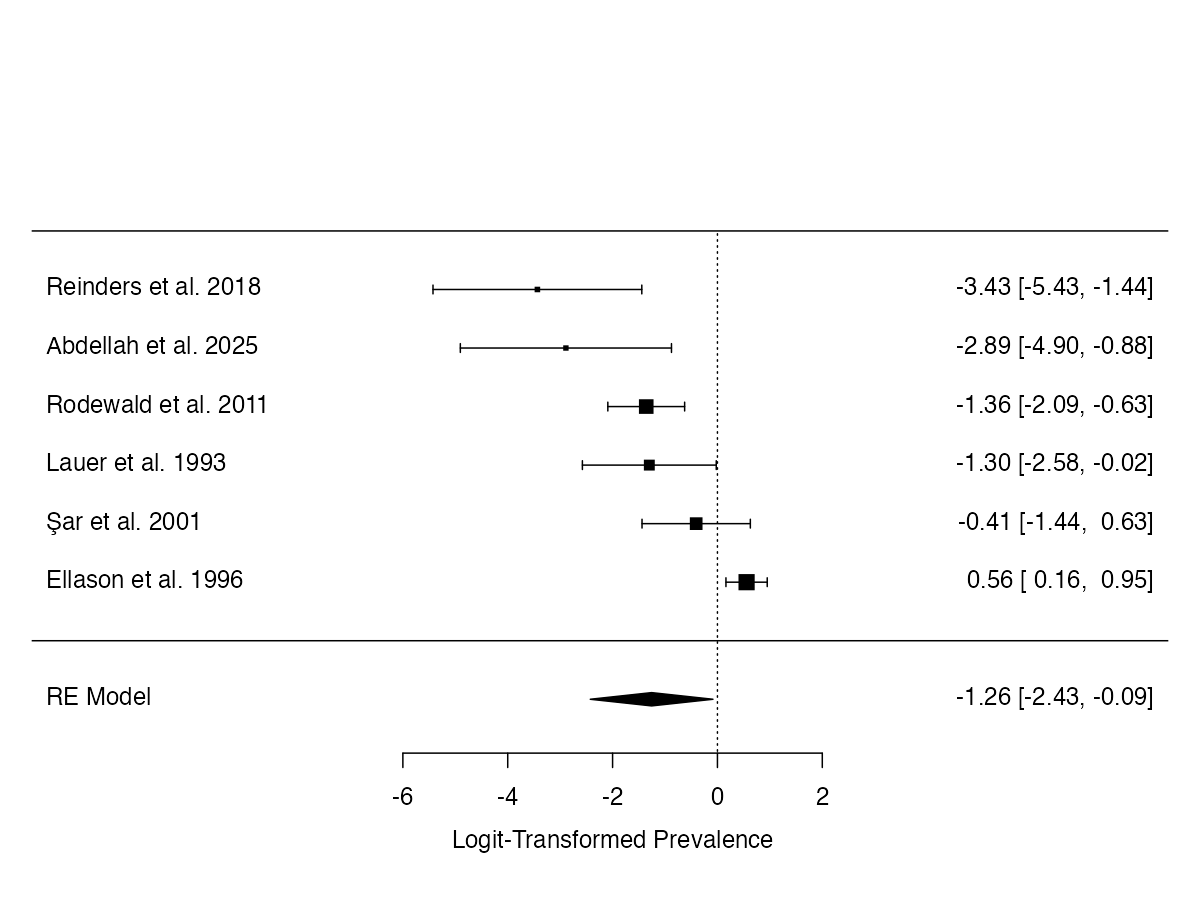

**Supplementary Figure 13.**

*Forest plot of logit-transformed prevalence estimates (with 95% CIs) for substance use disorder among individuals with DID.*

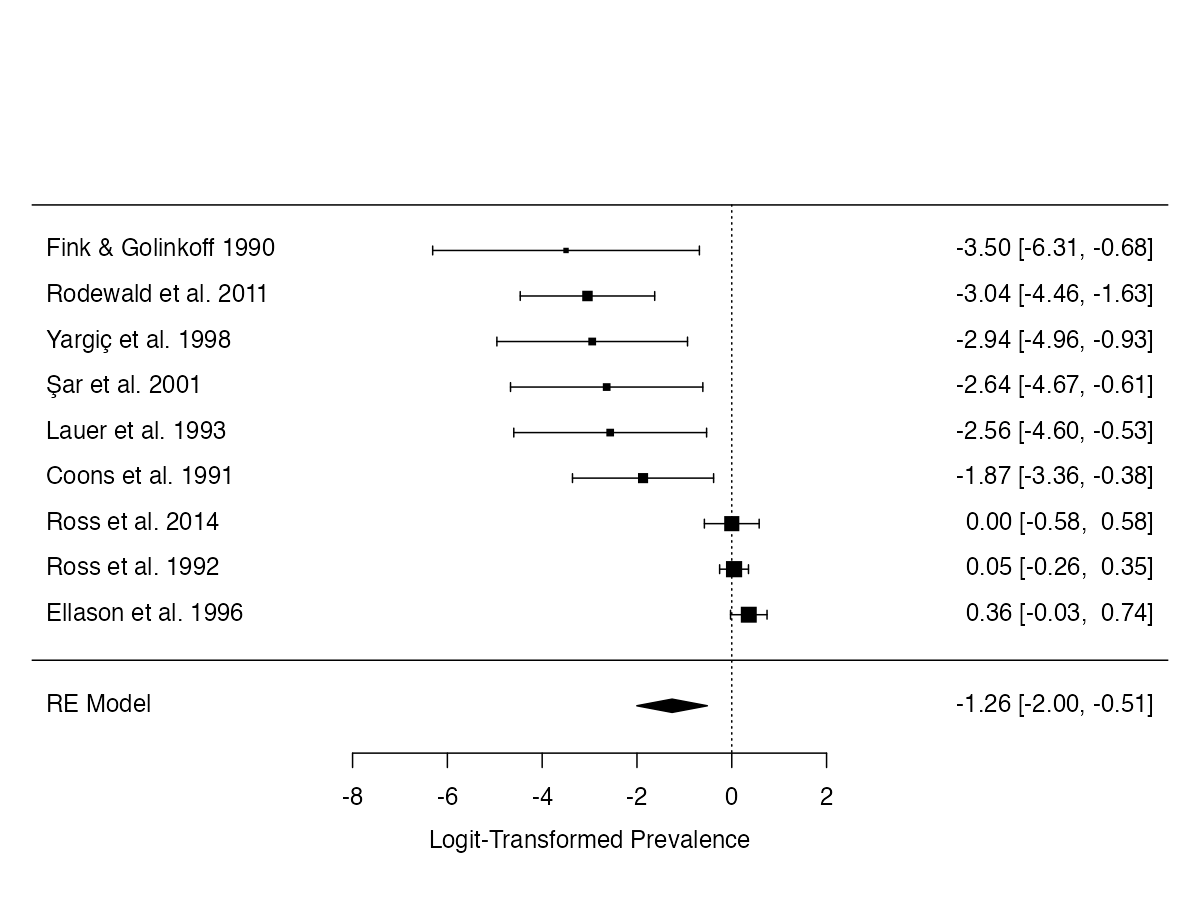

**Supplementary Figure 14.**

*Forest plot of logit-transformed prevalence estimates (with 95% CIs) for schizophrenia among individuals with DID.*

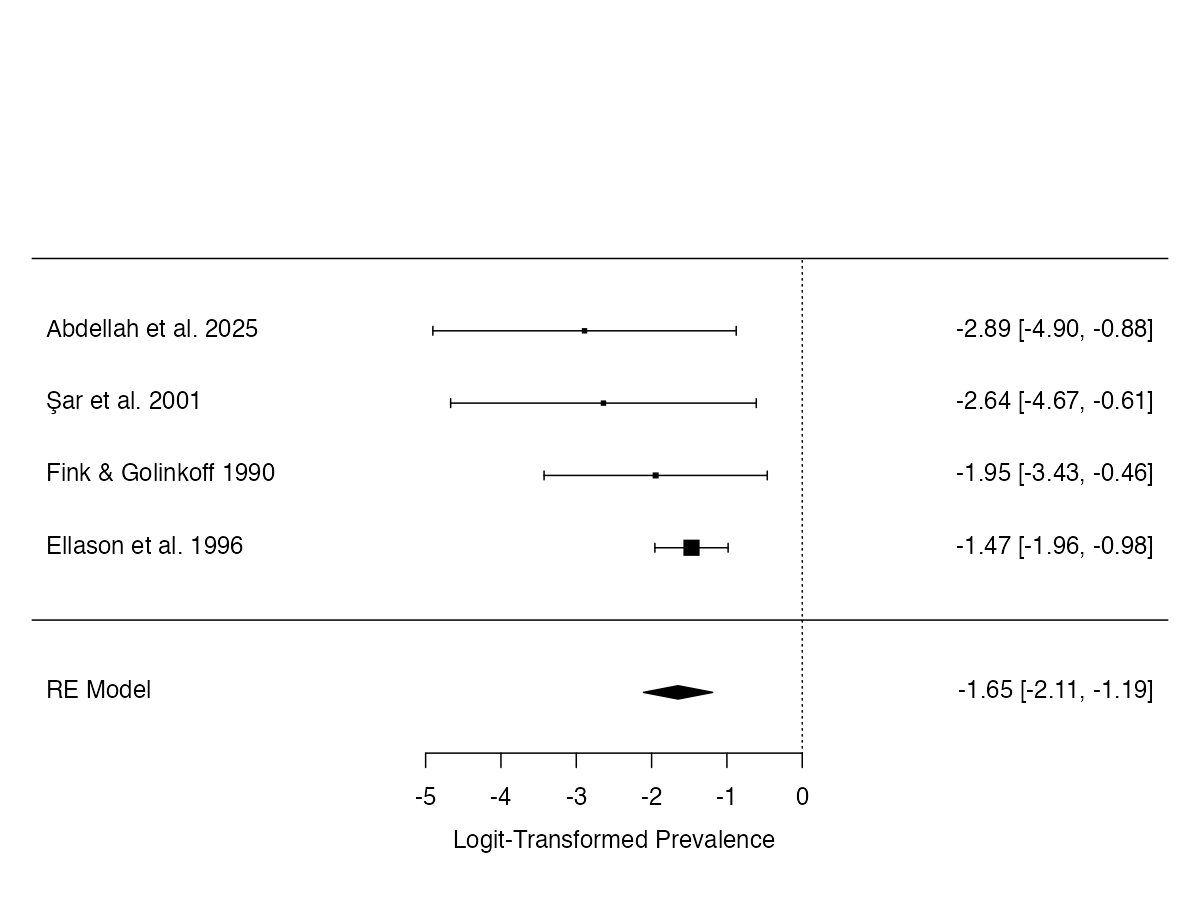

**Supplementary Figure 15.**

*Forest plot of logit-transformed prevalence estimates (with 95% CIs) for unspecified schizophrenia spectrum and other psychotic disorder among individuals with DID.*

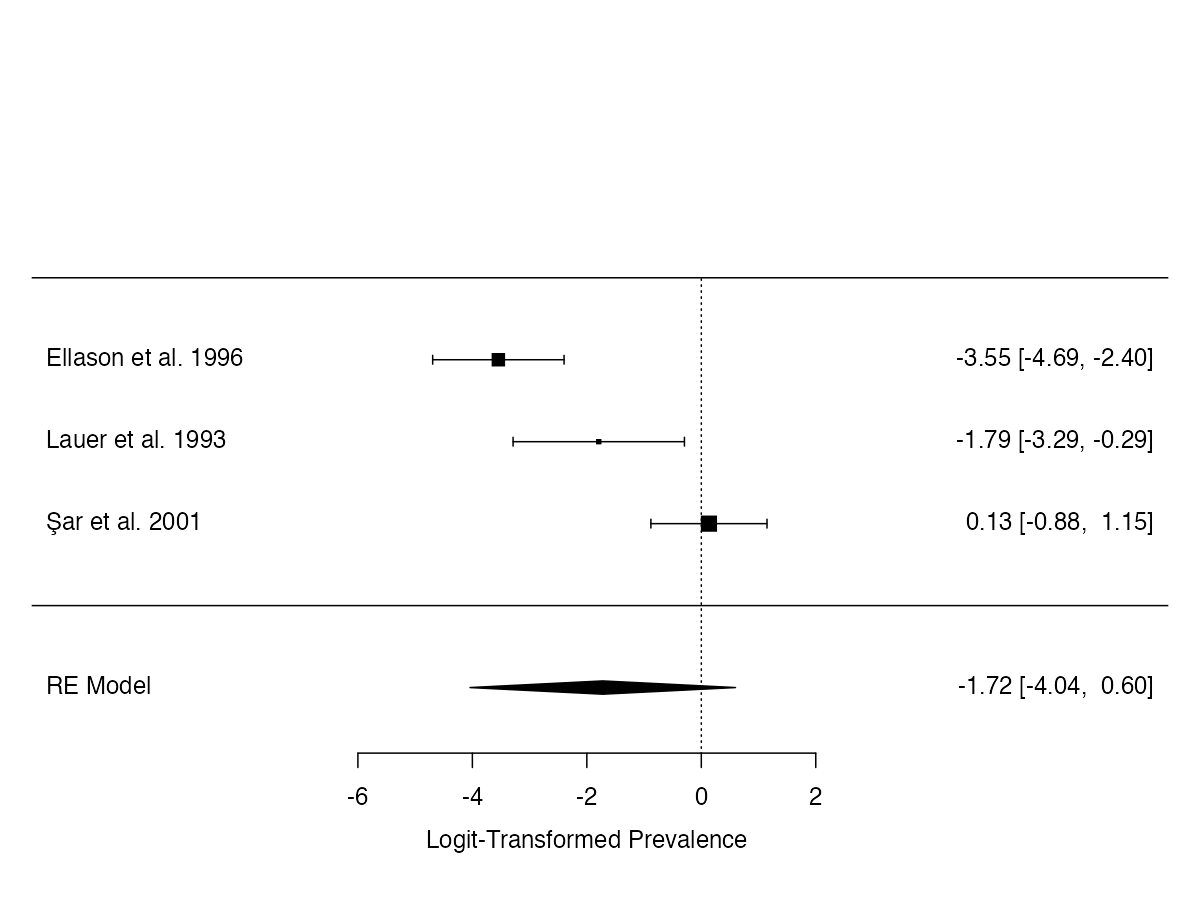

**Supplementary Figure 16.**

*Forest plot of logit-transformed prevalence estimates (with 95% CIs) for functional neurological symptom disorder among individuals with DID.*

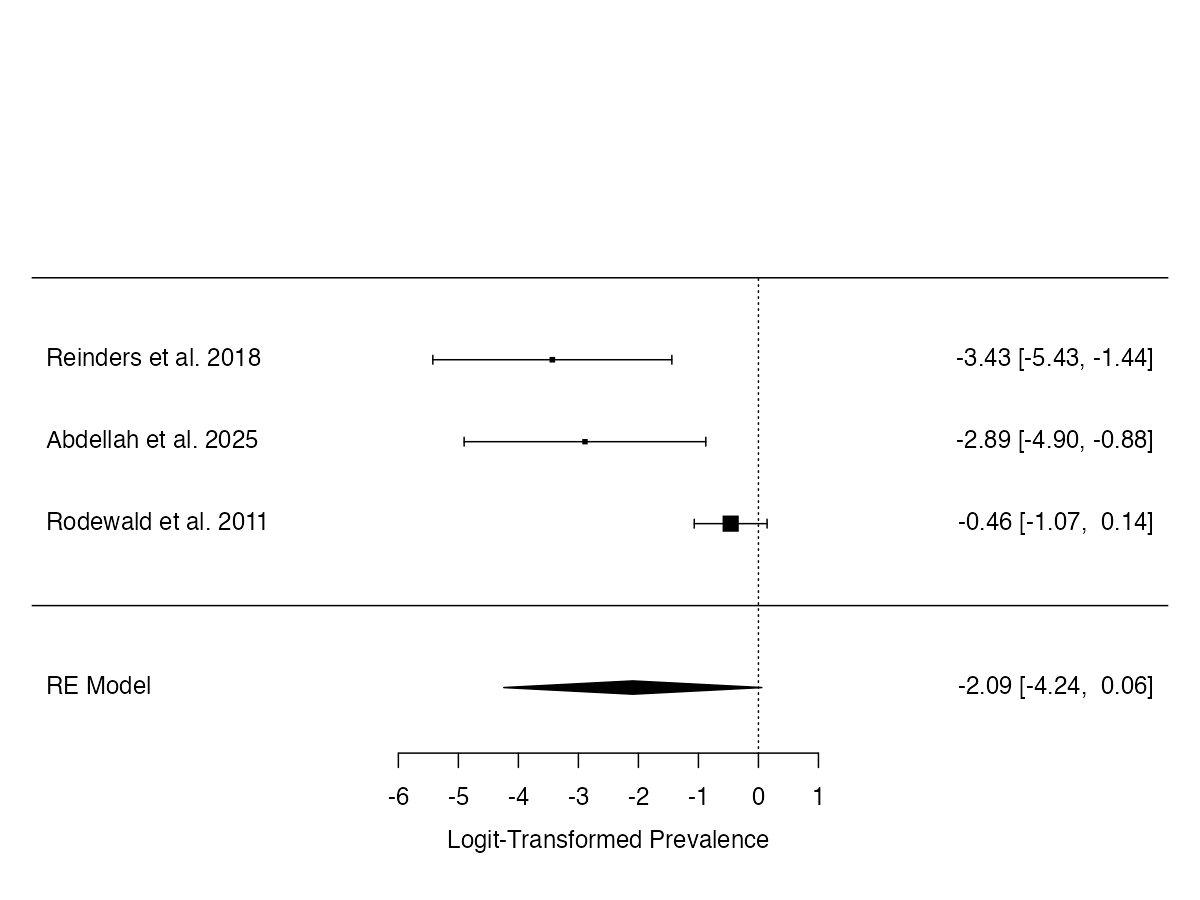

**Supplementary Figure 17.**

*Forest plot of logit-transformed prevalence estimates (with 95% CIs) for anorexia nervosa among individuals with DID.*

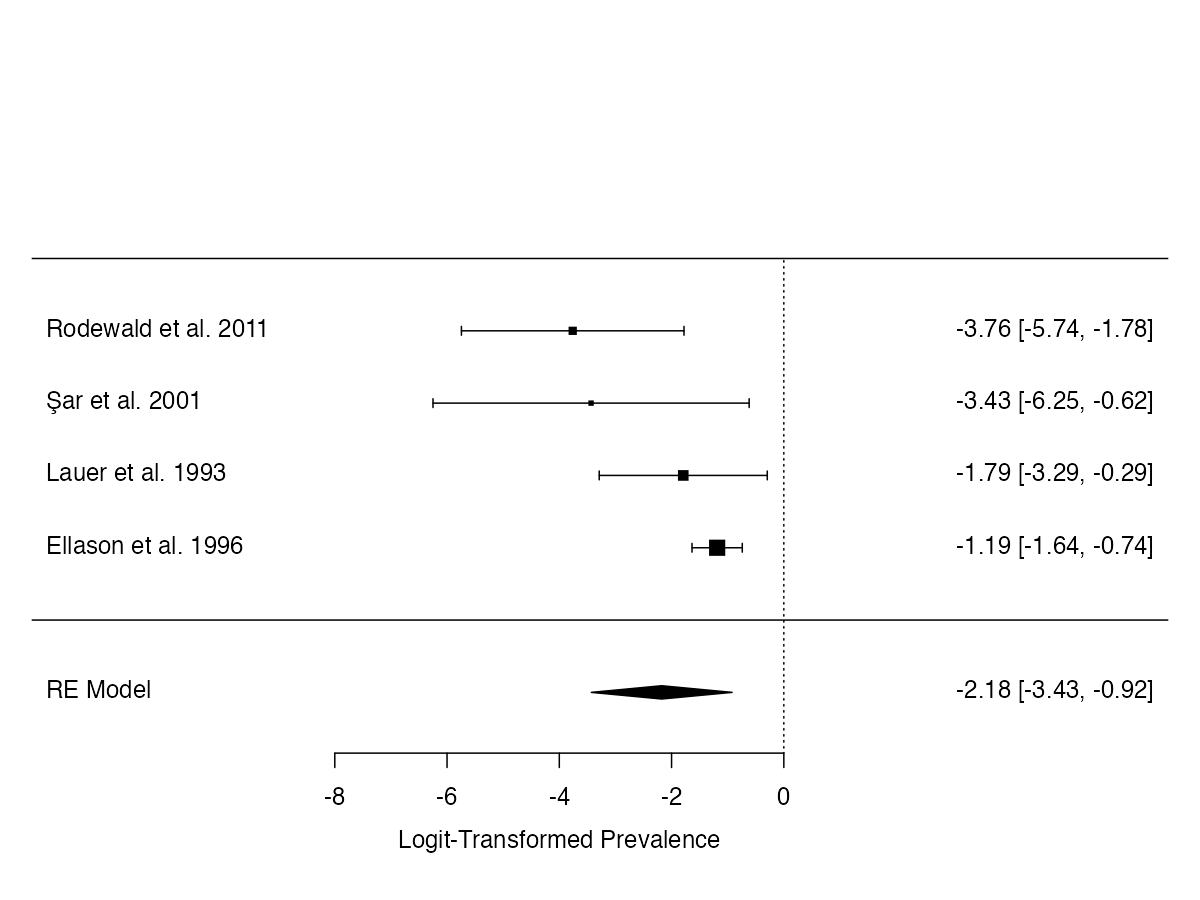

**Supplementary Figure 18.**

*Forest plot of logit-transformed prevalence estimates (with 95% CIs) for bipolar disorder among individuals with DID.*

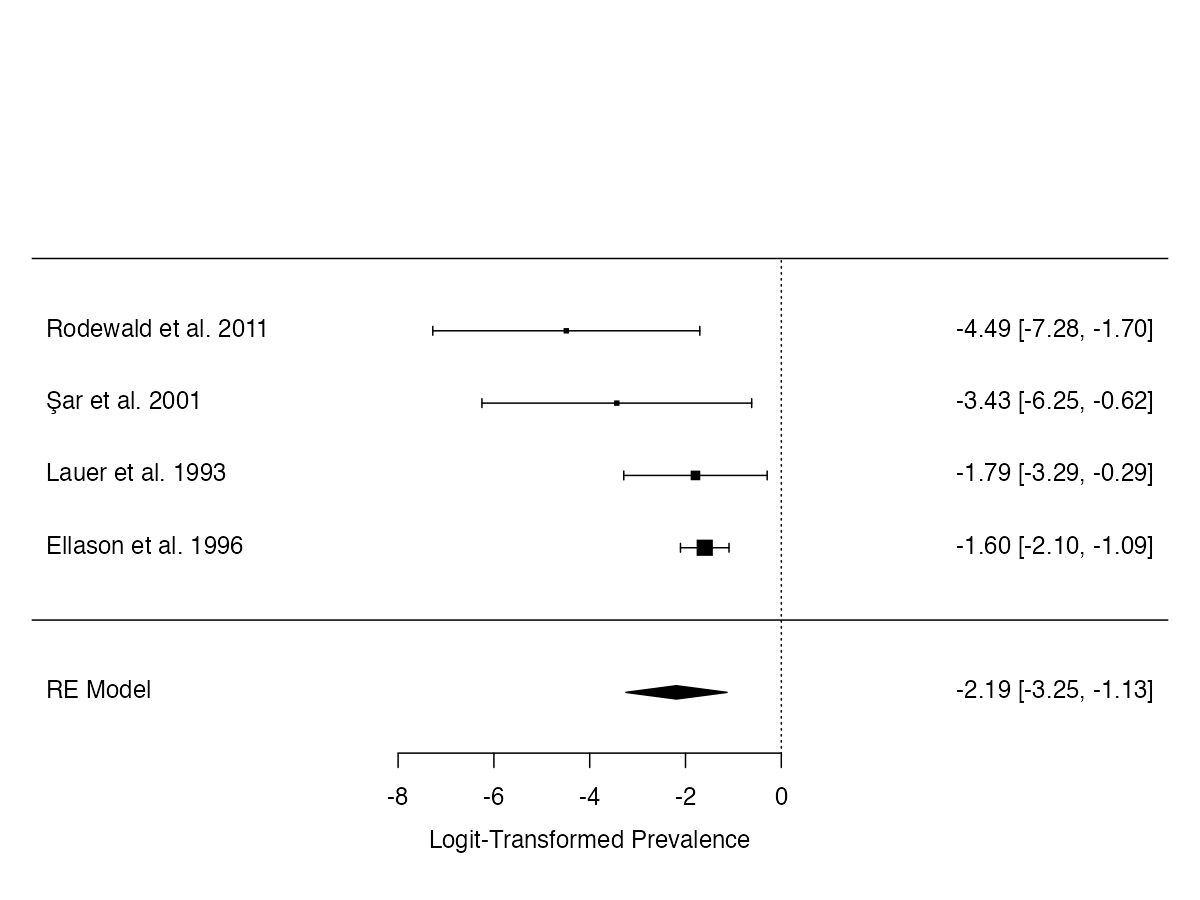

**Supplementary Figure 19.**

*Forest plot of logit-transformed prevalence estimates (with 95% CIs) for persistent depressive disorder among individuals with DID.*

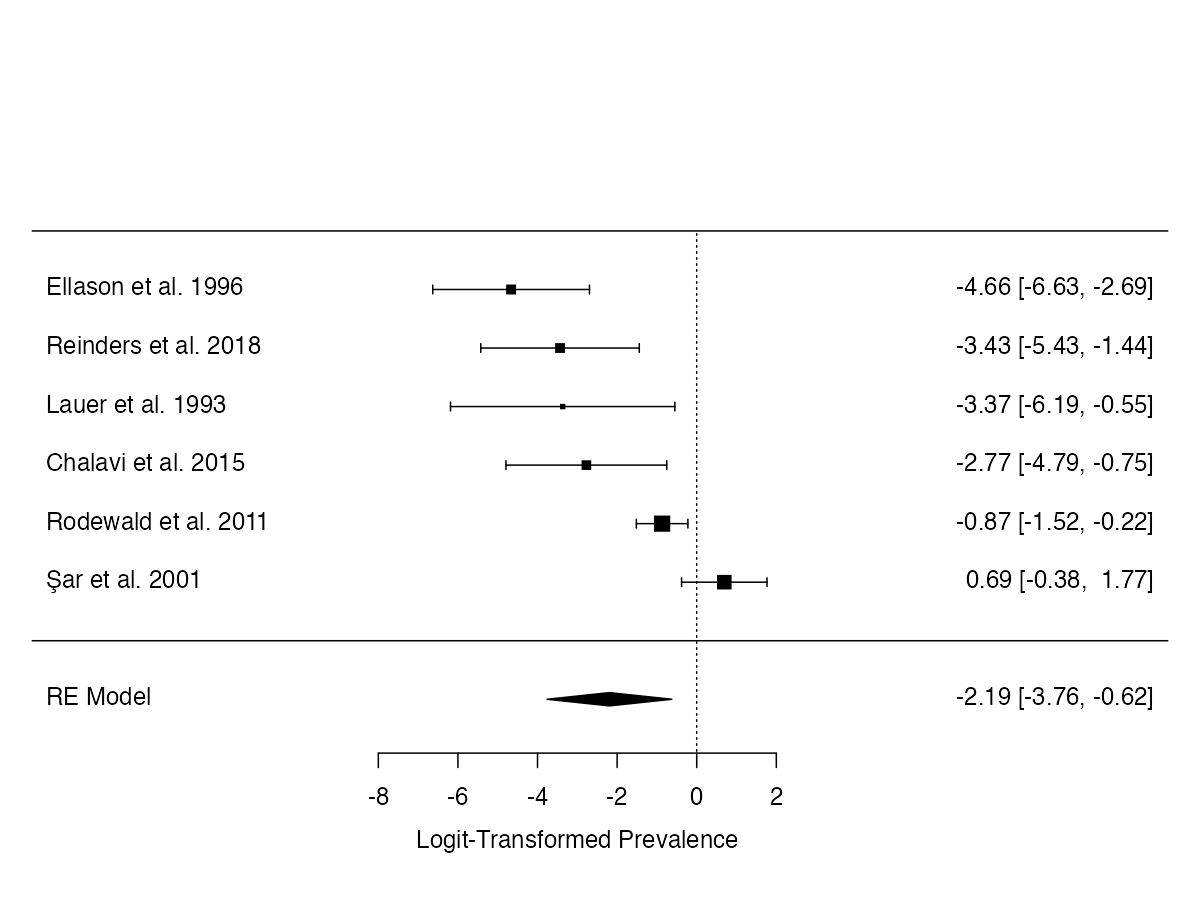

**Supplementary Figure 20.**

*Forest plot of logit-transformed prevalence estimates (with 95% CIs) for generalised anxiety disorder among individuals with DID.*

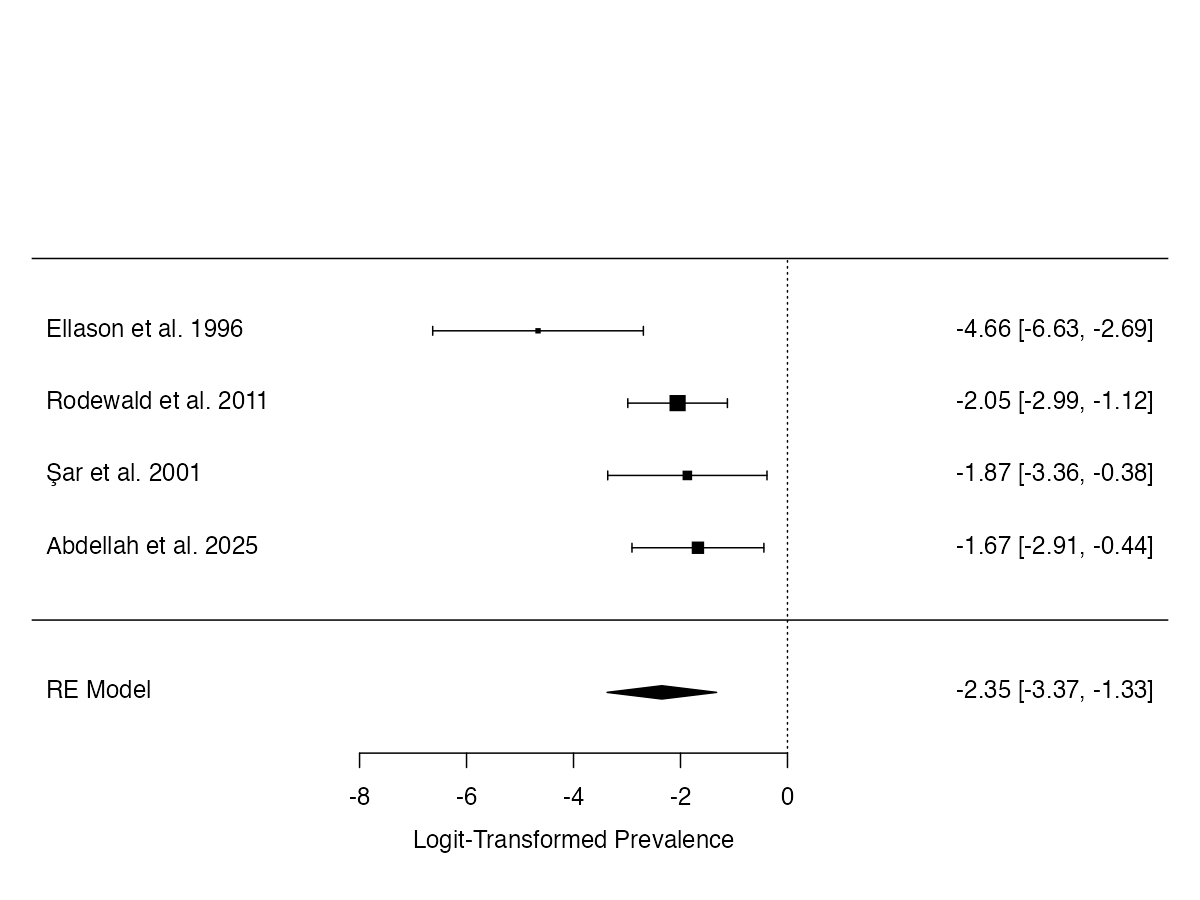

**Supplementary Figure 21.**

*Forest plot of logit-transformed prevalence estimates (with 95% CIs) for unspecified personality disorder among individuals with DID.*

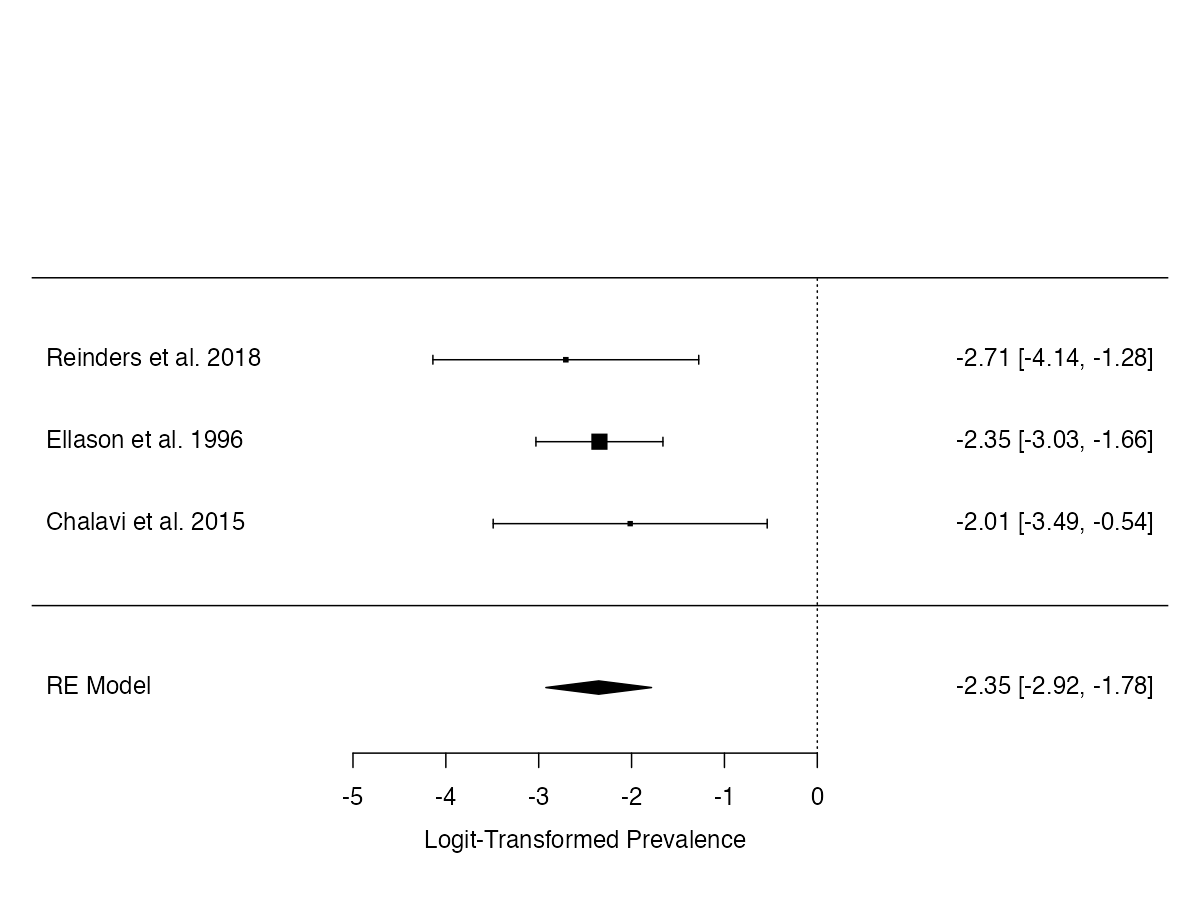

**Supplementary Figure 22.**

*Forest plot of logit-transformed prevalence estimates (with 95% CIs) for illness anxiety disorder among individuals with DID.*

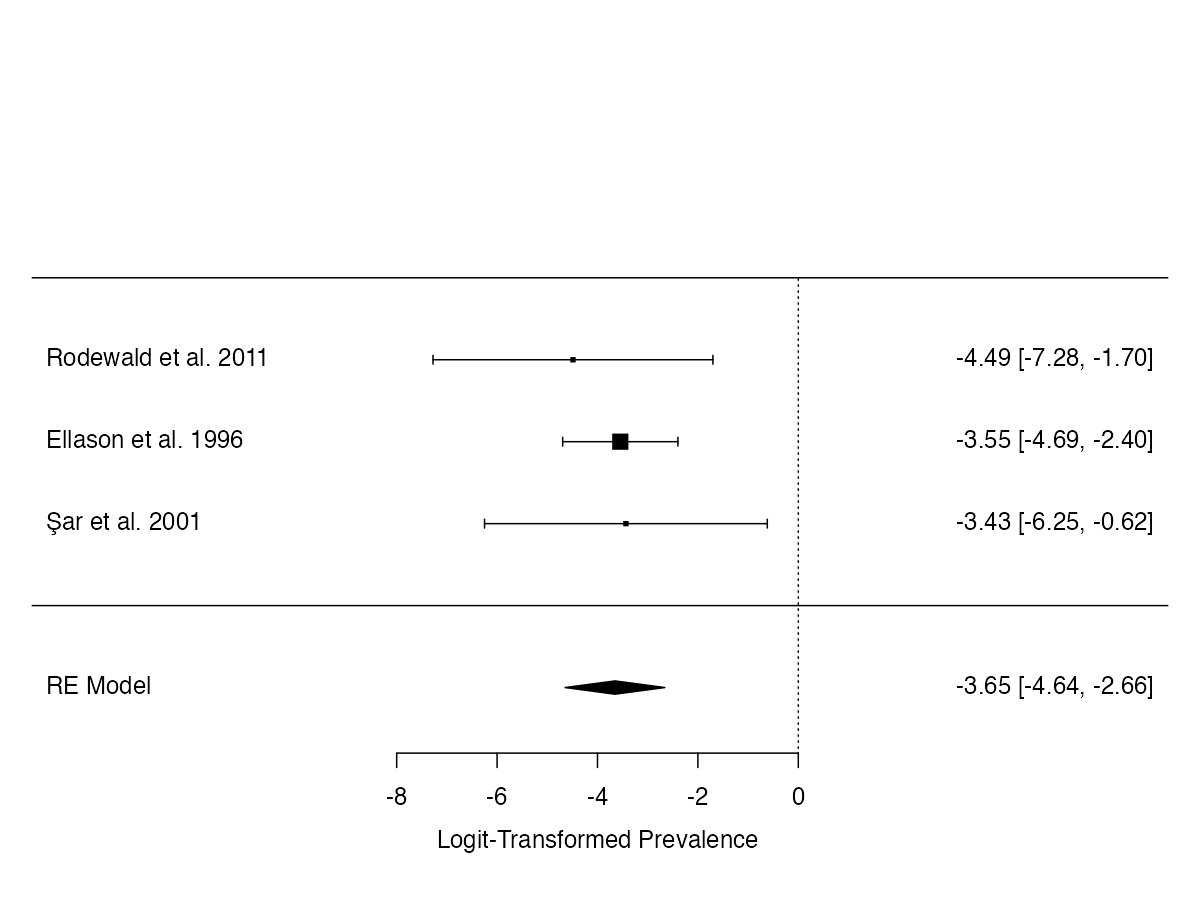

**Supplementary Figure 23.**

*Forest plot of logit-transformed prevalence estimates (with 95% CIs) for brief psychotic disorder among individuals with DID.*

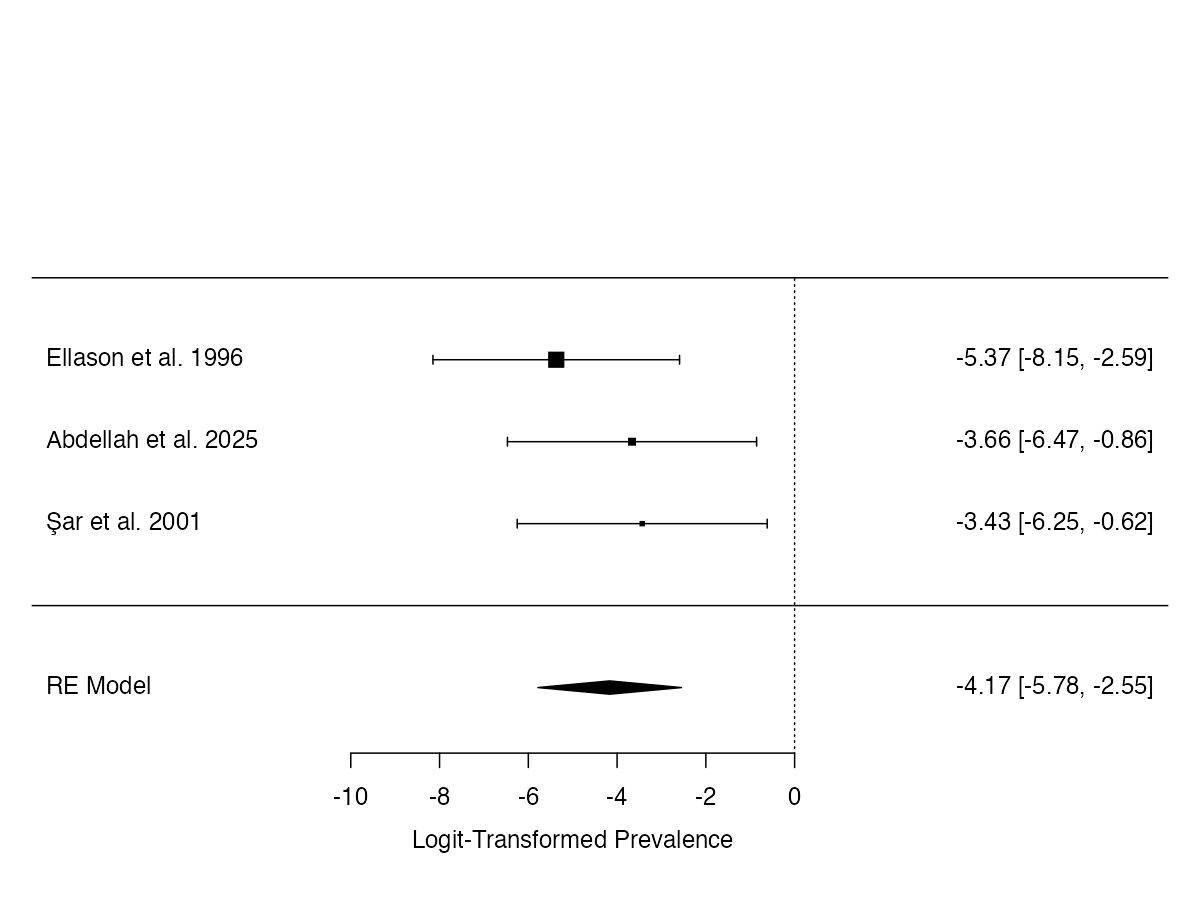

**Publication bias**

Supplementary Figures 24–46 present funnel plots of logit-transformed prevalence estimates as a function of standard errors (SEs) for all meta-analyses. Funnel plot asymmetry was assessed as an indicator of potential small-study effects. Meta-analyses with small study numbers (k < 10) were included as a reference point but should be interpreted with caution due to limited statistical power. Significant funnel plot asymmetry (see Table 2) was observed for panic disorder, social anxiety disorder, somatic symptom disorder, alcohol use disorder, obsessive-compulsive disorder, substance use disorder, functional neurological symptom disorder, anorexia nervosa, bipolar disorder, and persistent depressive disorder.

**Supplementary Figure 24.**

*Funnel plot of logit-transformed prevalence estimates (with 95% CIs) for posttraumatic stress disorder among individuals with DID.*

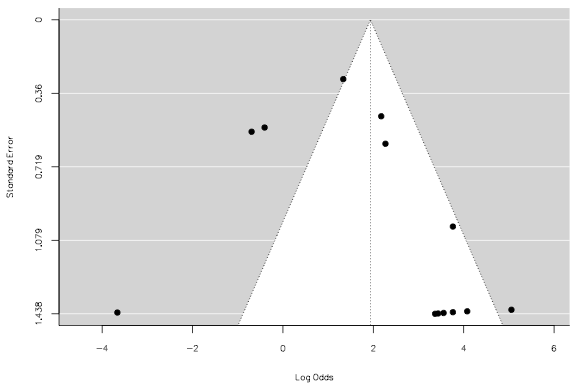

**Supplementary Figure 25.**

*Funnel plot of logit-transformed prevalence estimates (with 95% CIs) for major depressive disorder among individuals with DID.*

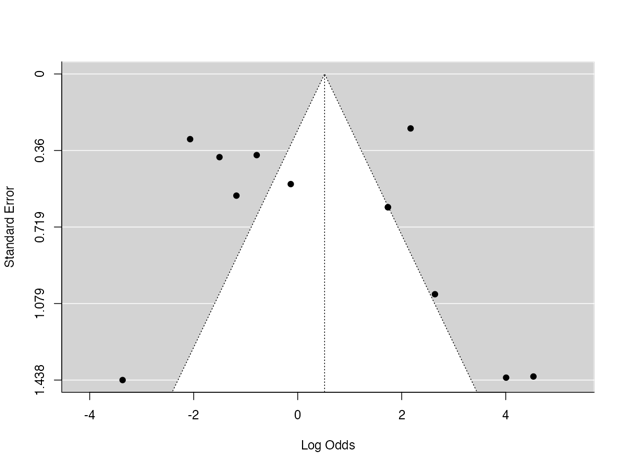

**Supplementary Figure 26.**

*Funnel plot of logit-transformed prevalence estimates (with 95% CIs) for borderline personality disorder among individuals with DID.*

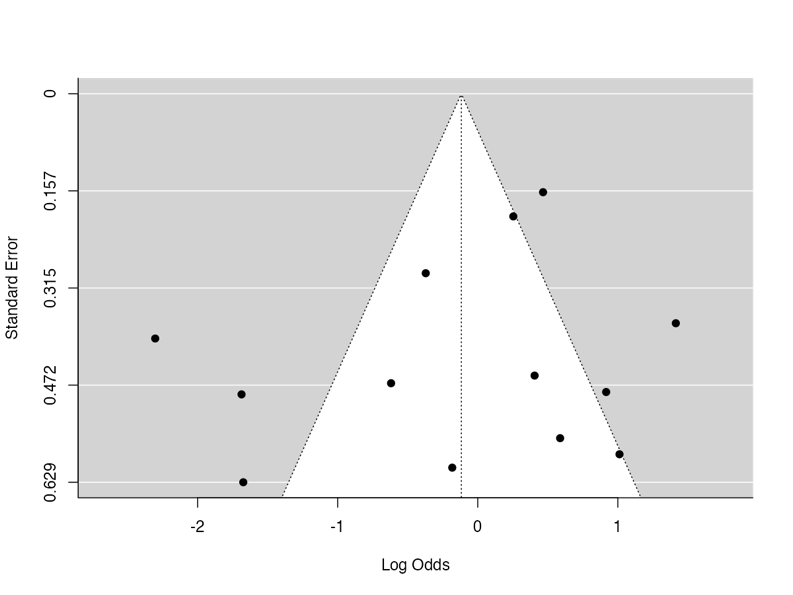

**Supplementary Figure 27.**

*Funnel plot of logit-transformed prevalence estimates (with 95% CIs) for panic disorder among individuals with DID.*

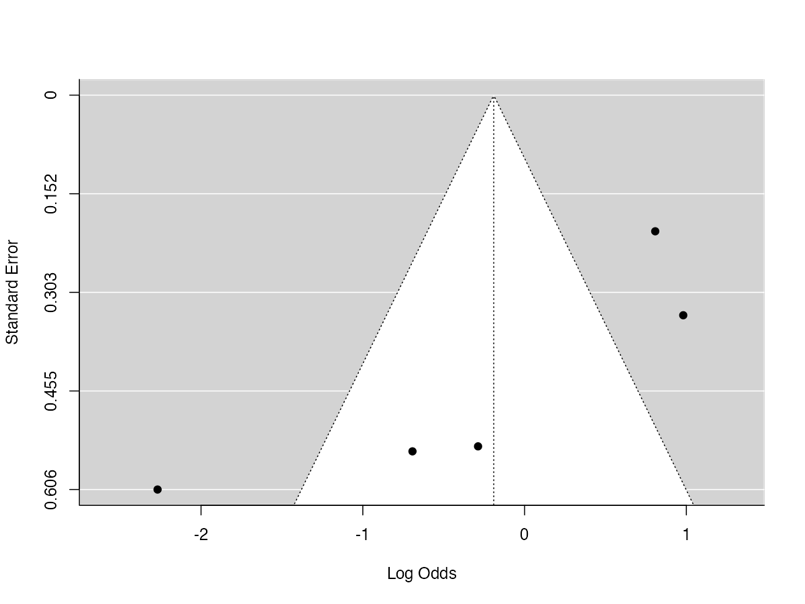

**Supplementary Figure 28.**

*Funnel plot of logit-transformed prevalence estimates (with 95% CIs) for schizoaffective disorder among individuals with DID.*

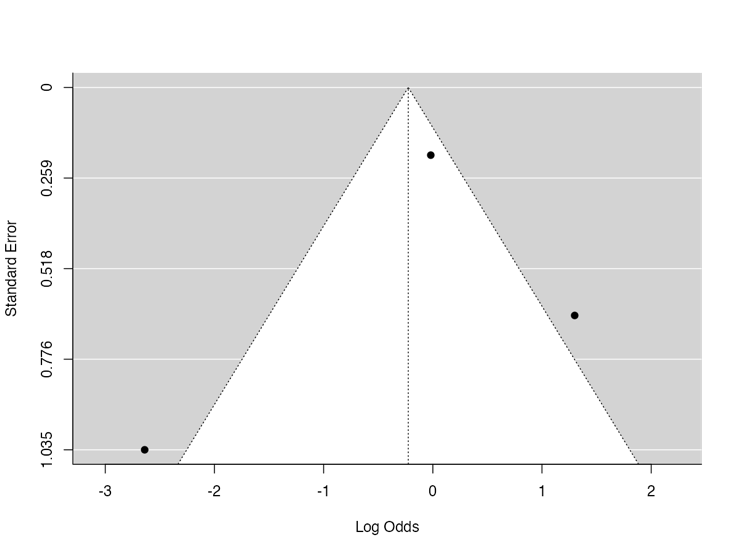

**Supplementary Figure 29.**

*Funnel plot of logit-transformed prevalence estimates (with 95% CIs) for social anxiety disorder among individuals with DID.*

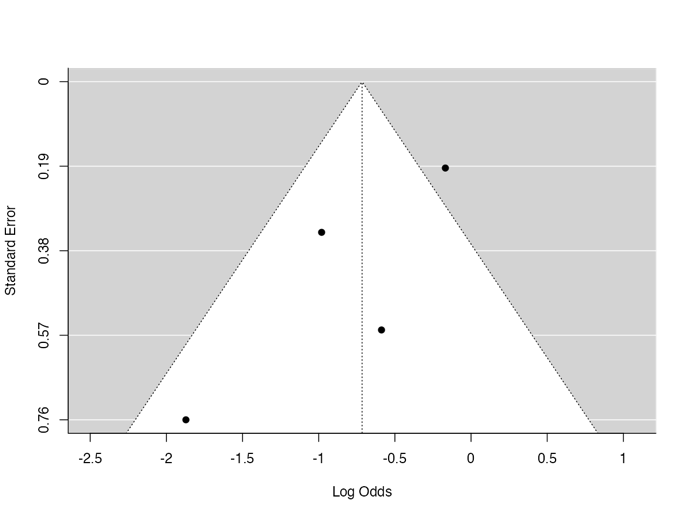

**Supplementary Figure 30.**

*Funnel plot of logit-transformed prevalence estimates (with 95% CIs) for somatic symptom disorder among individuals with DID.*

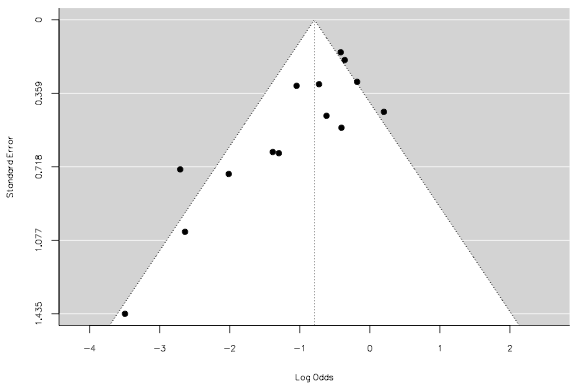

**Supplementary Figure 31.**

*Funnel plot of logit-transformed prevalence estimates (with 95% CIs) for alcohol use disorder among individuals with DID.*

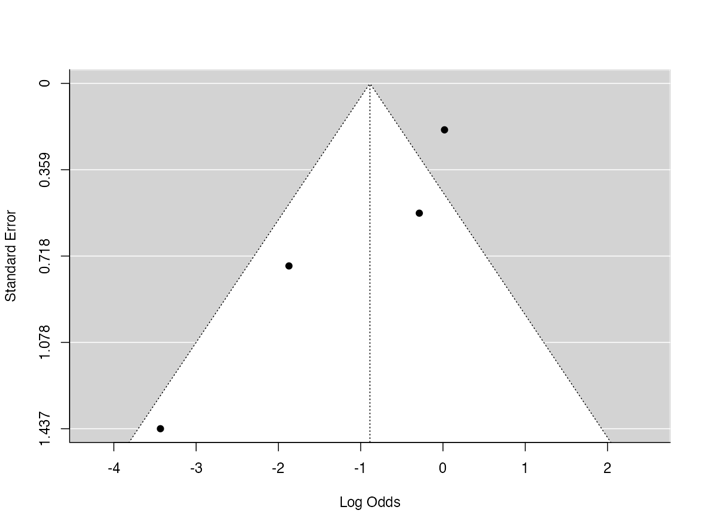

**Supplementary Figure 32.**

*Funnel plot of logit-transformed prevalence estimates (with 95% CIs) for specific phobia among individuals with DID.*

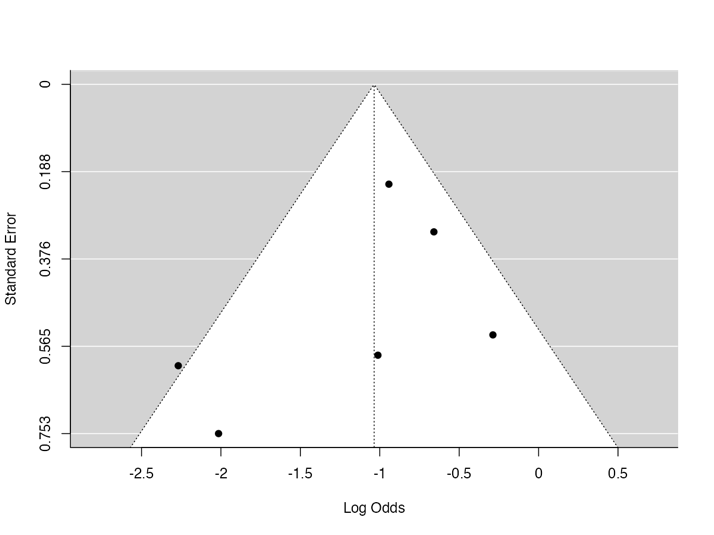

**Supplementary Figure 33.**

*Funnel plot of logit-transformed prevalence estimates (with 95% CIs) for agoraphobia among individuals with DID.*

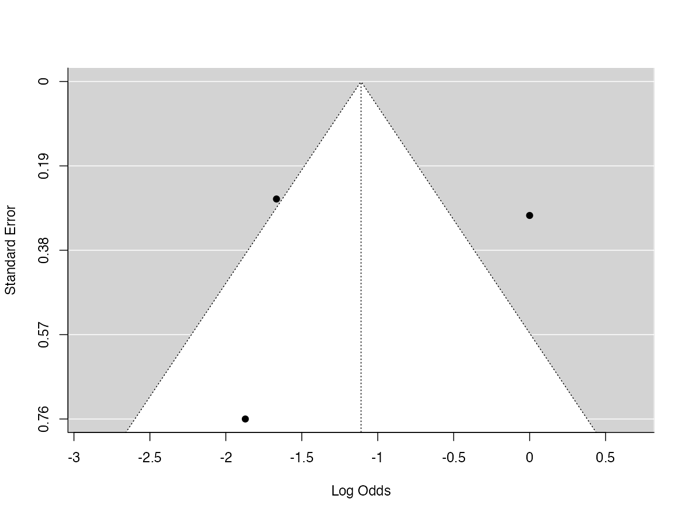

**Supplementary Figure 34.**

*Funnel plot of logit-transformed prevalence estimates (with 95% CIs) for bulimia nervosa among individuals with DID.*

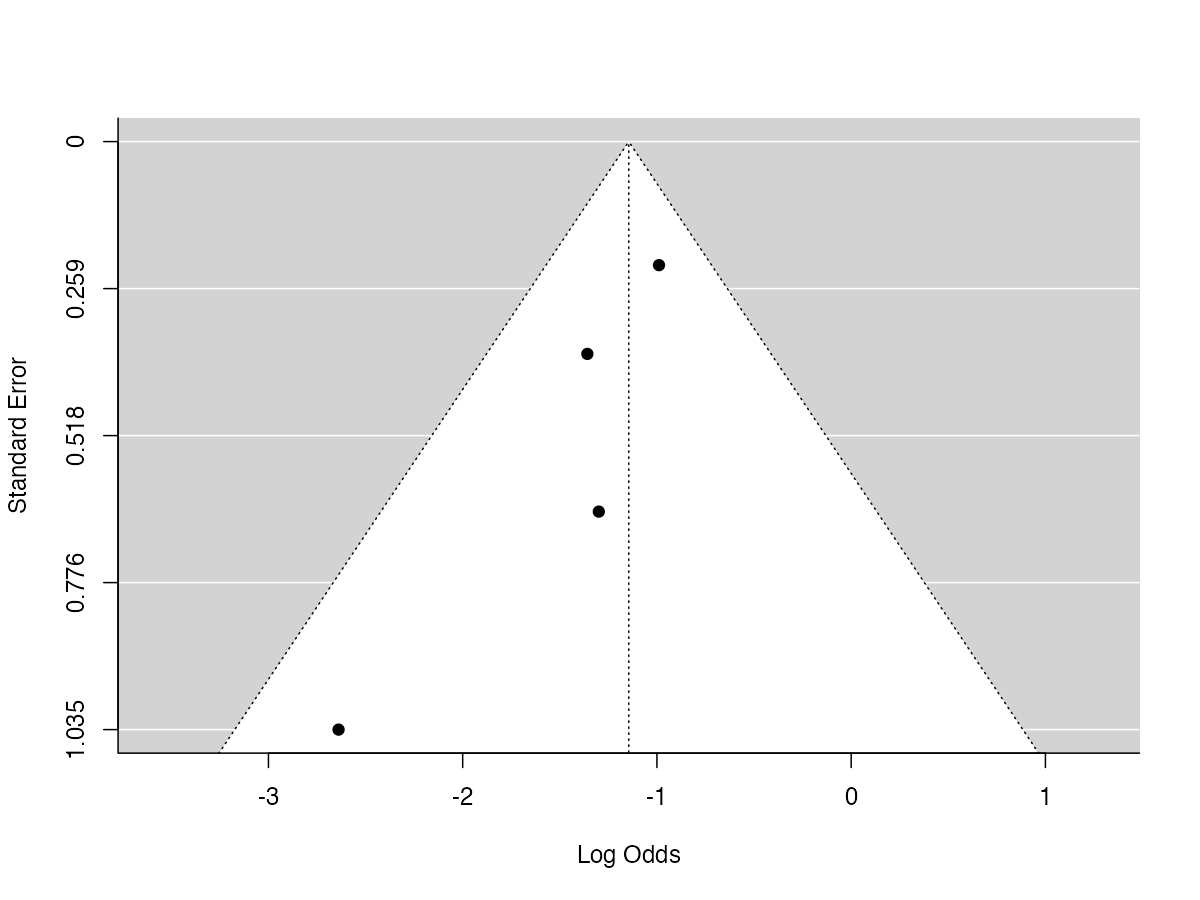

**Supplementary Figure 35.**

*Funnel plot of logit-transformed prevalence estimates (with 95% CIs) for obsessive-compulsive disorder among individuals with DID.*

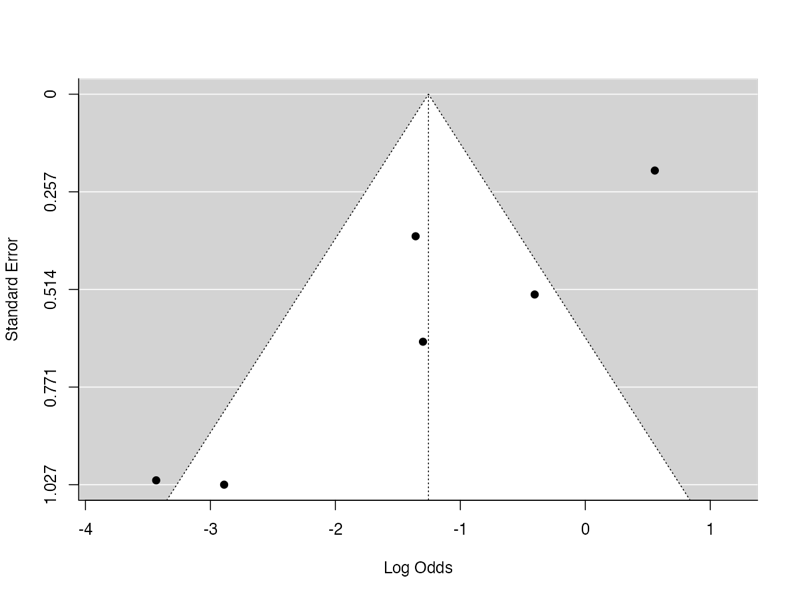

**Supplementary Figure 36.**

*Funnel plot of logit-transformed prevalence estimates (with 95% CIs) for substance use disorder among individuals with DID.*

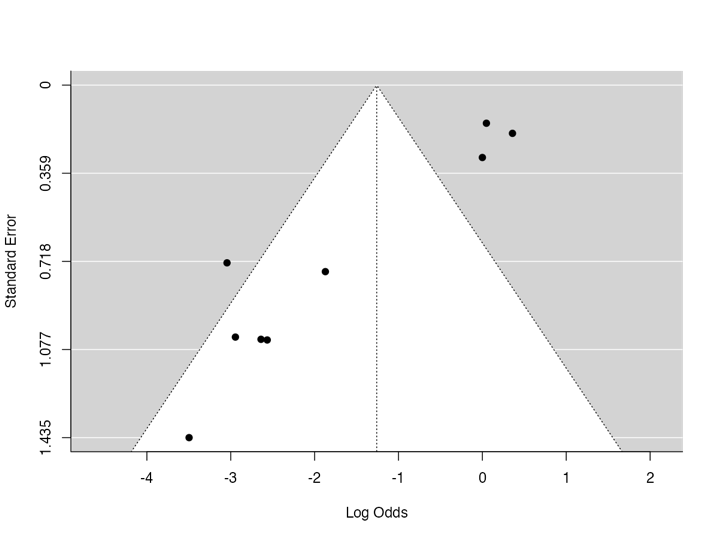

**Supplementary Figure 37.**

*Funnel plot of logit-transformed prevalence estimates (with 95% CIs) for schizophrenia among individuals with DID.*

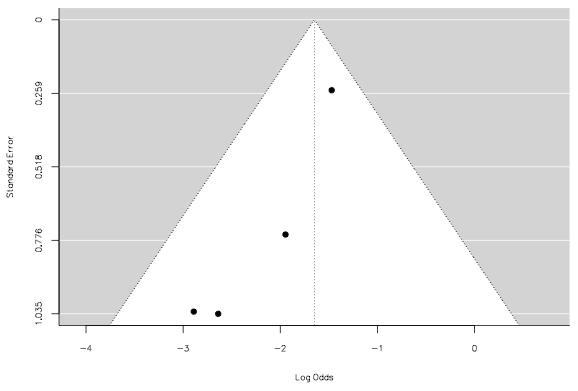

**Supplementary Figure 38.**

*Funnel plot of logit-transformed prevalence estimates (with 95% CIs) for unspecified schizophrenia spectrum and other psychotic disorder among individuals with DID.*

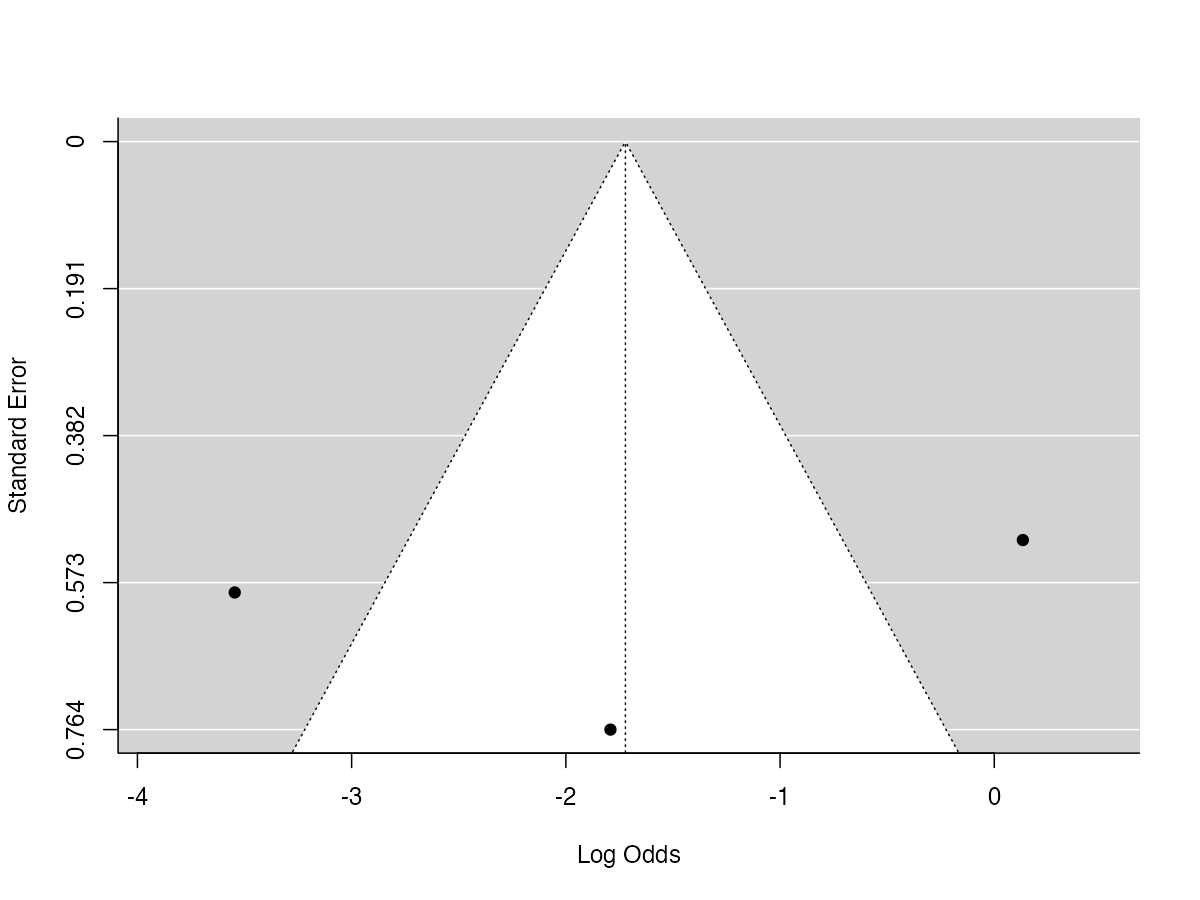

**Supplementary Figure 39.**

*Funnel plot of logit-transformed prevalence estimates (with 95% CIs) for functional neurological symptom disorder among individuals with DID.*

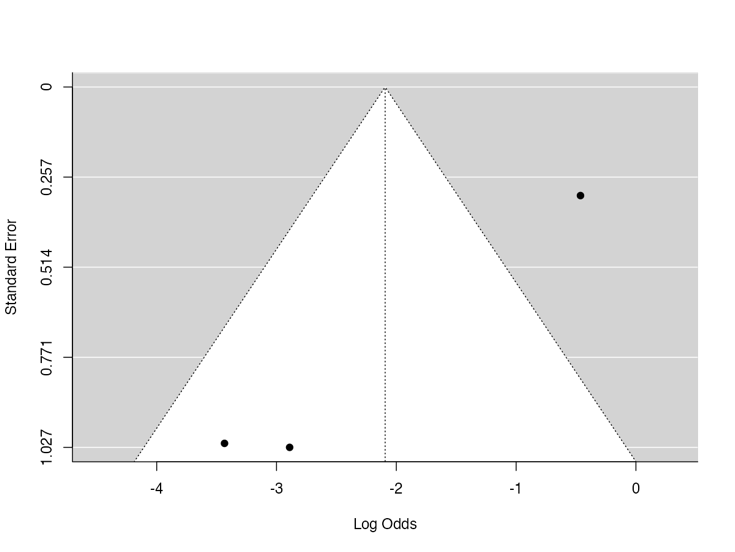

**Supplementary Figure 40.**

*Funnel plot of logit-transformed prevalence estimates (with 95% CIs) for anorexia nervosa among individuals with DID.*

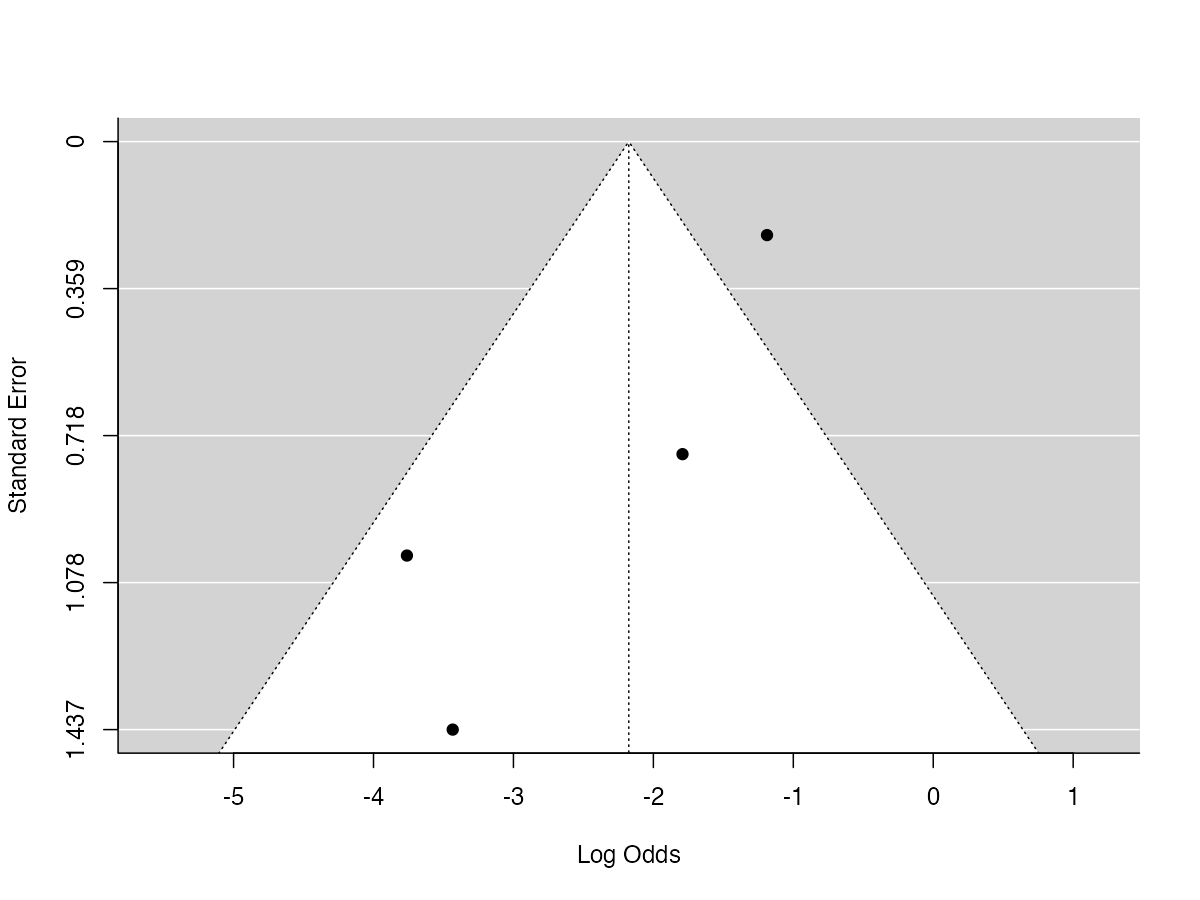

**Supplementary Figure 41.**

*Funnel plot of logit-transformed prevalence estimates (with 95% CIs) for bipolar disorder among individuals with DID.*

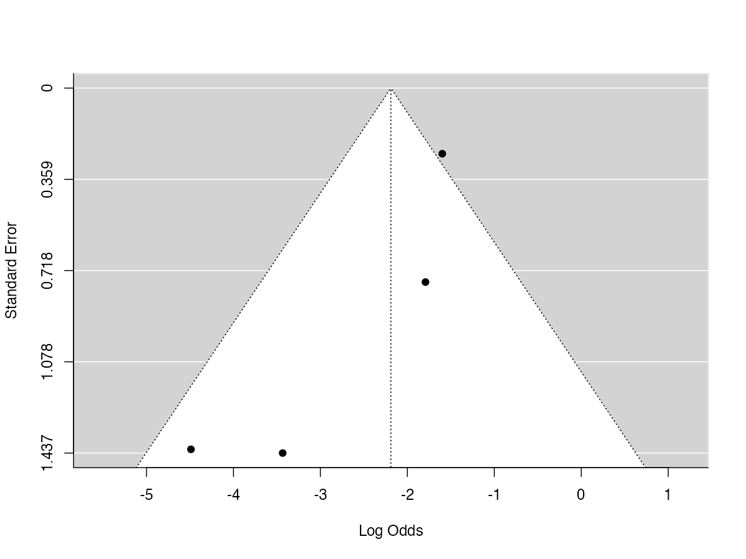

**Supplementary Figure 42.**

*Funnel plot of logit-transformed prevalence estimates (with 95% CIs) for persistent depressive disorder among individuals with DID.*

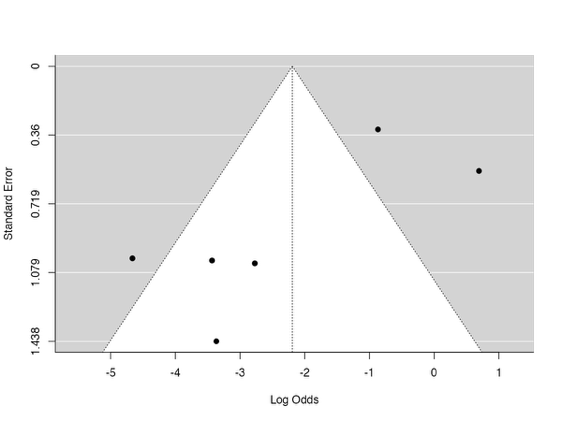

**Supplementary Figure 43.**

*Funnel plot of logit-transformed prevalence estimates (with 95% CIs) for generalised anxiety disorder among individuals with DID.*

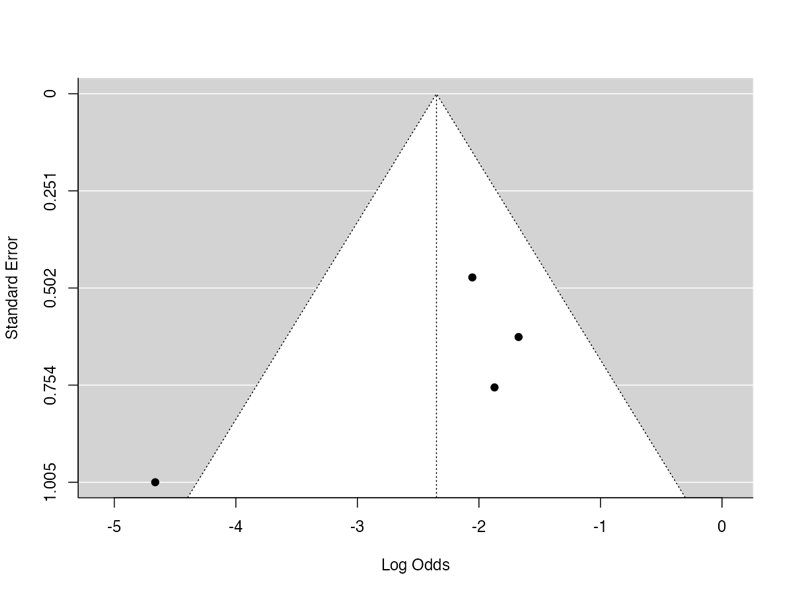

**Supplementary Figure 44.**

*Funnel plot of logit-transformed prevalence estimates (with 95% CIs) for unspecified personality disorder among individuals with DID.*

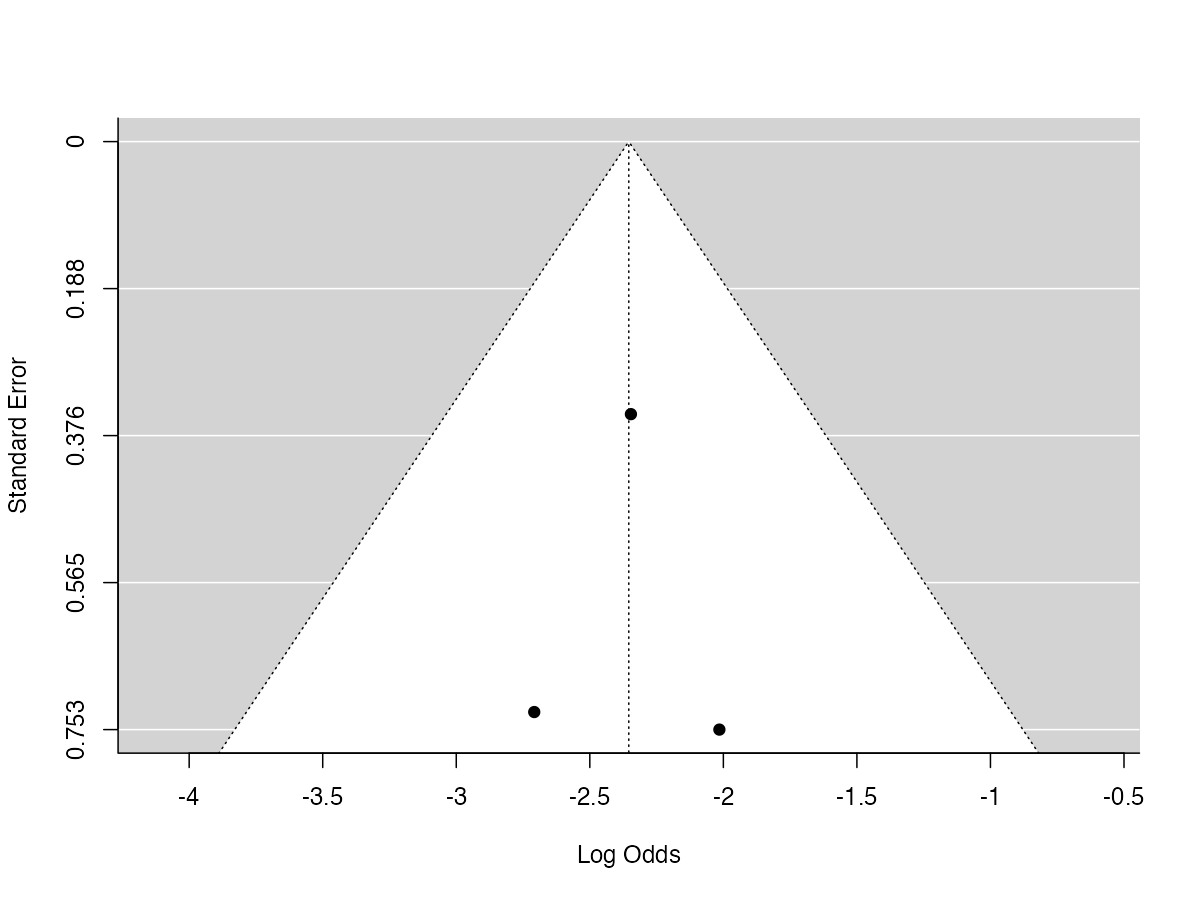

**Supplementary Figure 45.**

*Funnel plot of logit-transformed prevalence estimates (with 95% CIs) for illness anxiety disorder among individuals with DID.*

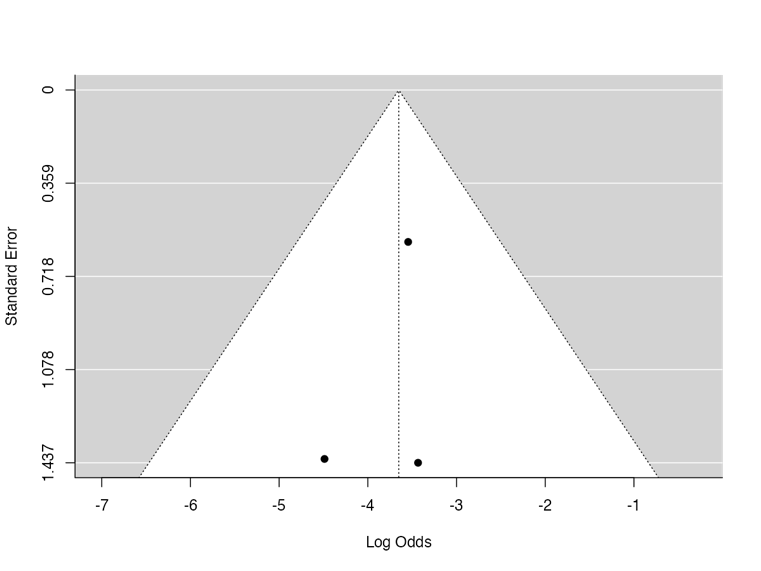

**Supplementary Figure 46.**

*Funnel plot of logit-transformed prevalence estimates (with 95% CIs) for brief psychotic disorder among individuals with DID.*

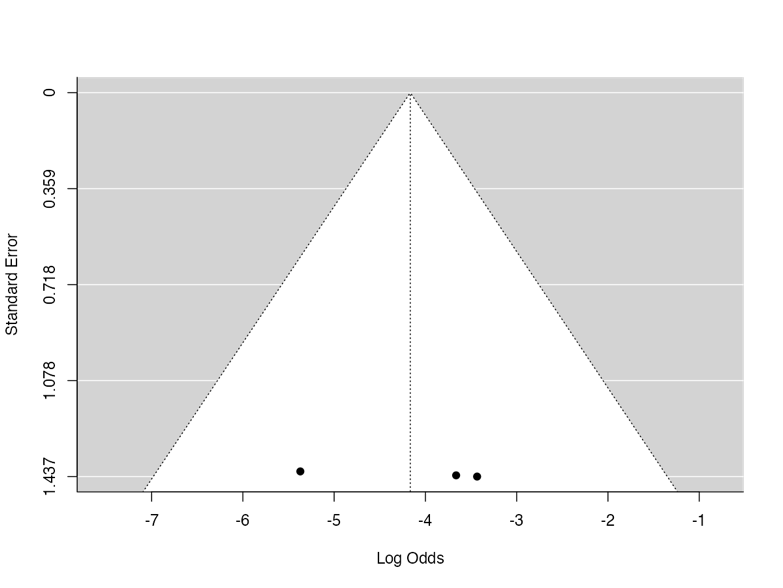

| **Supplementary Table 4.**  *Methodological quality ratings of included studies reporting psychiatric comorbidity prevalence estimates in DID.* | | | | | | | | | |
| --- | --- | --- | --- | --- | --- | --- | --- | --- | --- |
| Study | 1. Was the sample frame appropriate to address the target population? | 2. Were study participants sampled in an appropriate way? | 3. Was the sample size adequate? | 4. Were the study subjects and the setting described in detail? | 5. Was the data analysis conducted with sufficient coverage of the identified sample? | 6. Were valid methods used for the identification of the condition? | 7. Was the condition measured in a standard, reliable way for all participants? | 8. Was there appropriate statistical analysis? | 9. Was the response rate adequate, and if not, was the low response rate managed appropriately? |
| Abdellah et al. 2025 | No | Yes | No | Yes | Yes | Yes | Yes | Yes | No |
| Boon & Draijer 1993 | No | Unclear | Yes | No | Yes | Yes | Yes | Yes | No |
| Brand et al. 2006, 2009 | No | Unclear | Yes | No | Yes | No | Yes | Yes | No |
| Brand et al. 2021 | No | Unclear | Yes | No | No | No | Yes | Yes | No |
| Chalavi et al. 2015 | Unclear | Unclear | No | No | Yes | No | No | Yes | No |
| Coons et al. 1991 | No | No | No | No | Yes | No | No | No | No |
| Dale et al. 2009 | No | Yes | No | No | Yes | Yes | Yes | Yes | No |
| Ellason et al. 1996 | No | Yes | Yes | No | Yes | Yes | Yes | Yes | No |
| Fink & Golinkoff 1990 | No | Unclear | No | No | Yes | Yes | Yes | Yes | No |
| Lauer et al. 1993 | No | Unclear | No | No | Yes | Yes | Yes | Yes | No |
| Manchester et al. 1998 | No | Yes | No | Yes | Yes | Yes | Yes | Yes | No |
| Merker et al. 2021 | No | Unclear | Yes | Yes | Yes | Yes | Yes | Yes | No |
| Öztürk & Şar 2008 | No | Yes | Yes | No | Yes | Yes | Yes | Yes | No |
| Reinders et al. 2018 | No | Unclear | Yes | No | Yes | Yes | Yes | Yes | No |
| Rodewald et al. 2011 | No | Yes | Yes | No | Yes | Yes | Yes | Yes | No |
| Ross et al. 1989 | No | Yes | Yes | No | Yes | Yes | Yes | Yes | Yes |
| Ross et al. 1990 | No | Yes | Yes | No | Yes | Yes | Yes | Yes | Yes |
| Ross et al. 1992 | No | Yes | Yes | No | Yes | Yes | Yes | Yes | No |
| Ross et al. 2014 | No | Yes | Yes | No | Yes | Yes | Yes | Yes | No |
| Şar et al. 2001 | No | Unclear | No | No | Yes | Yes | Yes | Yes | No |
| Şar et al. 2007 | No | Unclear | Yes | No | Yes | Yes | Yes | Yes | No |
| Vermetten et al. 2006 | No | Unclear | No | Yes | Yes | Yes | Yes | Yes | No |
| Yargiç et al. 1998 | No | Yes | Yes | No | Yes | Yes | Yes | Yes | Yes |

| **Supplementary Table 5.**  *Psychiatric comorbidities reported across the included studies.* | | |
| --- | --- | --- |
| Study | DID sample size (*n*) | Psychiatric comorbidities reported (*n*) |
| Abdellah et al. 2025 | 19 | **Anxiety Disorders:** generalised anxiety disorder (3)  **Obsessive-Compulsive and Related Disorders:** body dysmorphic disorder (0), obsessive-compulsive disorder (1)  **Personality Disorders:** borderline personality disorder (3)  **Schizophrenia Spectrum and Other Psychotic Disorders:** brief psychotic disorder (0), schizophrenia (1)  **Somatic Symptom and Related Disorders:** functional neurological symptom disorder (1)  **Trauma- and Stressor-Related Disorders:** posttraumatic stress disorder (0) |
| Boon & Draijer 1993 | 49 | **Personality Disorders:** borderline personality disorder (20)  **Trauma- and Stressor-Related Disorders:** posttraumatic stress disorder (44) |
| Brand et al. 2006, 2009 | 77, 78* | **Personality Disorders:** borderline personality disorder (7)  **Trauma- and Stressor-Related Disorders:** posttraumatic stress disorder (78) |
| Brand et al. 2021 | 29 | **Trauma- and Stressor-Related Disorders:** posttraumatic stress disorder (29) |
| Chalavi et al. 2015 | 17 | **Anxiety Disorders:** specific phobia (2)  **Depressive Disorders:** major depressive disorder (4), persistent depressive disorder (1)  **Personality Disorders:** unspecified personality disorder (2)  **Somatic Symptom and Related Disorders:** somatic symptom disorder (2)  **Trauma- and Stressor-Related Disorders:** posttraumatic stress disorder (17) |
| Coons et al. 1991 | 15 | **Somatic Symptom and Related Disorders:** somatic symptom disorder (3)  **Substance-Related and Addictive Disorders:** alcohol use disorder (2), substance use disorder (2),  **Trauma- and Stressor-Related Disorders:** posttraumatic stress disorder (5) |
| Dale et al. 2009 | 14 | **Trauma- and Stressor-Related Disorders:** posttraumatic stress disorder (14) |
| Ellason et al. 1996 | 72, 103, 107* | **Anxiety Disorders:** agoraphobia (17), generalised anxiety disorder (1), panic disorder (74), specific phobia (30), social anxiety disorder (49)  **Bipolar and Related Disorders:** bipolar disorder (18)  **Depressive Disorders:** major depressive disorder (12), persistent depressive disorder (1)  **Feeding and Eating Disorders:** anorexia nervosa (25), bulimia nervosa (29)  **Obsessive-Compulsive and Related Disorders:** obsessive-compulsive disorder (68)  **Personality Disorders:** antisocial personality disorder (24), borderline personality disorder (58), dependent personality disorder (41), histrionic personality disorder (9), narcissistic personality disorder (13), obsessive-compulsive personality disorder (37), paranoid personality disorder (45), passive-aggressive personality disorder (13), schizoid personality disorder (6), schizotypal personality disorder (28), unspecified personality disorder (9)  **Schizophrenia Spectrum and Other Psychotic Disorders:** brief psychotic disorder (0), delusional disorder (2), schizoaffective disorder (53), schizophrenia (20), schizophreniform disorder (0), unspecified schizophrenia spectrum and other psychotic disorder (3)  **Somatic Symptom and Related Disorders:** illness anxiety disorder (3), somatic symptom disorder (44)  **Substance-Related and Addictive Disorders:** alcohol use disorder (54), substance use disorder (63)  **Trauma- and Stressor-Related Disorders:** posttraumatic stress disorder (57) |
| Fink & Golinkoff 1990 | 16 | **Schizophrenia Spectrum and Other Psychotic Disorders:** schizophrenia (2)  **Somatic Symptom and Related Disorders:** somatic symptom disorder (0)  **Substance-Related and Addictive Disorders:** substance use disorder (0) |
| Lauer et al. 1993 | 14 | **Anxiety Disorders:** panic disorder (6), social anxiety disorder (5), specific phobia (6),  **Bipolar and Related Disorders:** bipolar disorder (2)  **Depressive Disorders:** major depressive disorder (0), persistent depressive disorder (0)  **Feeding and Eating Disorders:** anorexia nervosa (2), bulimia nervosa (3)  **Obsessive-Compulsive and Related Disorders:** obsessive-compulsive disorder (3)  **Personality Disorders:** antisocial personality disorder (0), borderline personality disorder (9), dependent personality disorder (6), histrionic personality disorder (2), narcissistic personality disorder (1), obsessive-compulsive personality disorder (3), paranoid personality disorder (0), passive-aggressive personality disorder (1), sadistic personality disorder (0), schizoid personality disorder (0), schizotypal personality disorder (3)  **Schizophrenia Spectrum and Other Psychotic Disorders:** schizoaffective disorder (11), unspecified schizophrenia spectrum and other psychotic disorder (2)  **Somatic Symptom and Related Disorders:** somatic symptom disorder (3)  **Substance-Related and Addictive Disorders:** alcohol use disorder (6), substance use disorder (1) |
| Manchester et al. 1998 | 11 | **Personality Disorders:** Borderline personality disorder (5) |
| Merker et al. 2021 | 21 | **Trauma- and Stressor-Related Disorders:** posttraumatic stress disorder (21) |
| Öztürk & Şar 2008 | 27 | **Depressive Disorders:** major depressive disorder (27) |
| Reinders et al. 2018 | 32 | **Anxiety Disorders:** panic disorder (3), specific phobia (3)  **Depressive Disorders:** major depressive disorder (10), persistent depressive disorder (1)  **Neurodevelopmental Disorders:** attention-deficit/ hyperactivity disorder (1)  **Obsessive-Compulsive and Related Disorders:** obsessive-compulsive disorder (1)  **Personality Disorders:** borderline personality disorder (5), unspecified personality disorder (2)  **Somatic Symptom and Related Disorders:** functional neurological symptom disorder (1), somatic symptom disorder (2)  **Trauma- and Stressor-Related Disorders:** posttraumatic stress disorder (29) |
| Rodewald et al. 2011 | 44 | **Anxiety Disorders:** agoraphobia (22), generalised anxiety disorder (5), panic disorder (32), social anxiety disorder (12), specific phobia (15),  **Bipolar and Related Disorders:** bipolar disorder (0)  **Depressive Disorders:** major depressive disorder (8), persistent depressive disorder (13)  **Feeding and Eating Disorders:** anorexia nervosa (1), bulimia nervosa (9)  **Obsessive-Compulsive and Related Disorders:** obsessive-compulsive disorder (9)  **Somatic Symptom and Related Disorders:** functional neurological symptom disorder (17), illness anxiety disorder (0), somatic symptom disorder (20),  **Substance-Related and Addictive Disorders:** substance use disorder (2),  **Trauma- and Stressor-Related Disorders:** posttraumatic stress disorder (43) |
| Ross et al. 1989 | 20 | **Depressive Disorders:** major depressive disorder (17)  **Personality Disorders:** borderline personality disorder (12)  **Somatic Symptom and Related Disorders:** somatic symptom disorder (7) |
| Ross et al. 1990 | 50 | **Somatic Symptom and Related Disorders:** Somatic symptom disorder (13) |
| Ross et al. 1992 | 166 | **Depressive Disorders:** major depressive disorder (149),  **Personality Disorders:** borderline personality disorder (102)  **Somatic Symptom and Related Disorders:** somatic symptom disorder (66)  **Substance-Related and Addictive Disorders:** substance use disorder (85) |
| Ross et al. 2014 | 46 | **Depressive Disorders:** major depressive disorder (46)  **Personality Disorders:** borderline personality disorder (37)  **Somatic Symptom and Related Disorders:** somatic symptom disorder (15)  **Substance-Related and Addictive Disorders:** substance use disorder (23) |
| Şar et al. 2001 | 15 | **Anxiety Disorders:** agoraphobia (2), generalised anxiety disorder (2), panic disorder (5), social anxiety disorder (2), specific phobia (4)  **Bipolar and Related Disorders:** bipolar disorder (0)  **Depressive Disorders:** major depressive disorder (7), persistent depressive disorder (10)  **Feeding and Eating Disorders:** anorexia nervosa (0), bulimia nervosa (1)  **Obsessive-Compulsive and Related Disorders:** obsessive-compulsive disorder (6)  **Personality Disorders:** borderline personality disorder (11)  **Schizophrenia Spectrum and Other Psychotic Disorders:** brief psychotic disorder (0), delusional disorder (0), schizoaffective disorder (1), schizophrenia (1), schizophreniform disorder (2), unspecified schizophrenia spectrum and other psychotic disorder (8)  **Somatic Symptom and Related Disorders:** illness anxiety disorder (0), somatic symptom disorder (6)  **Substance-Related and Addictive Disorders:** alcohol use disorder (0), substance use disorder (1)  **Trauma- and Stressor-Related Disorders:** posttraumatic stress disorder (6) |
| Şar et al. 2007 | 21 | **Personality Disorders:** borderline personality disorder (15) |
| Vermetten et al. 2006 | 15 | **Depressive Disorders:** major depressive disorder (14)  **Somatic Symptom and Related Disorders:** somatic symptom disorder (1)  **Trauma- and Stressor-Related Disorders:** posttraumatic stress disorder (15) |
| Yargiç et al. 1998 | 20 | **Depressive Disorders:** major depressive disorder (17)  **Personality Disorders:** borderline personality disorder (7)  **Somatic Symptom and Related Disorders:** somatic symptom disorder (11)  **Substance-Related and Addictive Disorders:** substance use disorder (1) |
| *Different sample sizes were reported for different psychiatric comorbidities; all reported sample sizes are presented, separated by commas.  ***Abbreviations.*** n = number of participants; DID = dissociative identity disorder. | | |

| **Supplementary Table 6.**  *Pooled prevalence estimates following exclusion of outlying or influential studies.* | | | | | | | | | | | | | |
| --- | --- | --- | --- | --- | --- | --- | --- | --- | --- | --- | --- | --- | --- |
| Disorder | Exclusion | Outliers, *n* | DFBETAS, *n* | Cook’s *D*, *n* | Studies Excluded | *k* | *N* | Back-transformed pooled prevalence, % (95% CI) | Leave-one-out prevalence range, % | *τ²* (SE) | *I²* (%) | *Q* (*p*) | Funnel plot asymmetry (*p*) |
| Posttraumatic stress disorder | Outlier | 1 | 0 | 0 | Abdellah et al. 2025 | 12 | 401 | 90.38 (78.24, 96.07) | 84.88, 90.36 | 1.90 (1.36) | 80.20 | 55.56 (<.001***) | 0.002** |
| Major depressive disorder | - | 0 | 0 | 0 | - | - | - | - | 56.01, 68.51 | - | - | - | - |
| Borderline personality disorder | Outlier/ Influential | 1 | 0 | 1 | Brand et al. 2006, 2009 | 12 | 516 | 52.24 (41.24, 63.02) | 43.70, 52.24 | 0.42 (0.30) | 77.73 | 49.39 (<.001***) | 0.36 |
| Panic disorder | Influential | 0 | 1 | 1 | Reinders et al. 2018 | 4 | 180 | 58.49 (41.29, 73.83) | 36.71, 58.48 | 0.34 (0.42) | 71.46 | 10.51 (0.02*) | 0.022* |
| Somatic symptom disorder | Influential | 0 | 0 | 3 | Ross et al. 1992;  Ellason et al. 1996;  Reinders et al. 2018 | 11 | 272 | 30.01 (21.77, 39.80) | 29.04, 33.90 | 0.24 (0.23) | 51.84 | 20.77 (0.02*) | 0.003** |
| Specific phobia | Influential | 0 | 1 | 0 | Reinders et al. 2018 | 5 | 197 | 29.59 (23.40, 36.63) | 23.50, 29.49 | 0.00 (0.12) | 2.14 | 4.09  (0.59) | 0.39 |
| Obsessive-compulsive disorder | - | 0 | 0 | 0 | - | - | - | - | 17.19, 28.81 | - | - | - | - |
| Substance use disorder | Influential | 0 | 1 | 4 | Ross et al. 1992;  Ellason et al. 1996;  Rodewald et al. 2011;  Ross et al. 2014 | 5 | 80 | 7.59 (3.31, 16.41) | 14.92, 28.92 | 0.00 (0.73) | 0.00 | 1.38  (0.85) | 0.26 |
| Persistent depressive disorder | Influential | 0 | 0 | 1 | Şar et al. 2001 | 5 | 214 | 5.31 (1.00, 23.88) | 5.32, 16.17 | 2.90 (2.95) | 80.25 | 20.25 (<.001***) | <.001*** |
| Note. Outliers were identified using studentised residuals *Z*>\|3\|. Influential cases were identified using DFBETAS >\|1\| or Cook's distance >4/k. *p* < .05*, *p* < .01**, *p* < .001***.  ***Abbreviations.*** k = number of studies; N = total sample size. | | | | | | | | | | | | | |

| **Supplementary Table 7.**  *Sensitivity analyses of pooled prevalence estimates.* | | | | | | | |
| --- | --- | --- | --- | --- | --- | --- | --- |
| Disorder | *k* | *N* | Back-transformed pooled prevalence, % (95% CI) | *τ²* (SE) | *I²* (%) | *Q* (*p*) | Funnel plot asymmetry (*p*) |
| **Posttraumatic stress disorder** | | | | | | | |
| *Sample size ≥ 20* | 7 | 325 | 93.52 (85.21, 97.29) | 0.79 | 64.33 | 16.82 (.010**) | <.001*** |
| *Structured DID interview* | 11 | 327 | 88.08 (72.99, 95.27) | 1.8 | 78.40 | 46.30 (<.001***) | .26 |
| *Structured comorbidity interview* | 9 | 266 | 85.81 (65.34, 95.09) | 2.10 | 80.62 | 41.28 (<.001***) | .44 |
| **Major depressive disorder** | | | | | | | |
| *Sample size ≥ 20* | 8 | 462 | 72.73 (35.80, 92.73) | 4.54 | 95.84 | 168.25 (<.001***) | .032* |
| *Structured DID interview* | 10 | 483 | 56.59 (24.99, 83.59) | 4.14 | 94.73 | 170.72 (<.001***) | .16 |
| *Structured comorbidity interview* | 9 | 459 | 66.55 (29.90, 90.26) | 4.84 | 95.23 | 167.796 (<.001***) | .37 |
| **Borderline personality disorder** | | | | | | | |
| *Sample size ≥ 20* | 9 | 534 | 46.33 (31.32, 62.06) | 0.81 | 89.59 | 76.84 (<.001***) | .39 |
| *Structured DID interview* | 10 | 476 | 53.10 (40.71, 65.11) | 0.46 | 80.37 | 45.84 (<.001***) | .40 |
| *Structured comorbidity interview* | 10 | 435 | 58.25 (48.08, 67.77) | 0.25 | 66.69 | 27.02 (.001**) | .35 |
| **Panic disorder** | | | | | | | |
| *Sample size ≥ 20* | 3 | 183 | 49.08 (19.25, 79.58) | 1.36 | 91.88 | 24.620(<.001***) | .020* |
| *Structured comorbidity interview* | 4 | 180 | 36.38 (41.29, 73.83) | 0.34 | 71.46 | 10.51 (.015*) | .022* |
| **Somatic symptom disorder** | | | | | | | |
| *Sample size ≥ 20* | 8 | 485 | 36.38 (29.09, 44.35) | 0.12 | 58.56 | 18.89 (.018*) | .075 |
| *JBI quality score ≥ 7* | 3 | 90 | 36.98 (21.87, 55.13) | 0.26 | 60.62 | 5.08 (.079) | .30 |
| *Structured DID interview* | 11 | 445 | 29.42 (21.99, 61.87) | 0.22 | 64.17 | 27.91 (.002**) | <.001*** |
| *Structured comorbidity interview* | 10 | 453 | 37.50 (31.65, 43.73) | 0.05 | 33.83 | 13.60 (.14) | .080 |
| **Alcohol use disorder** | | | | | | | |
| *Structured DID interview* | 3 | 136 | 37.80 (16.99, 64.31) | 0.56 | 65.84 | 5.85 (.054) | .026* |
| *Structured comorbidity interview* | 3 | 136 | 37.80 (16.99, 64.31) | 0.56 | 65.84 | 5.85 (.054) | .026* |
| **Specific phobia** | | | | | | | |
| *Sample size ≥ 20* | 3 | 183 | 24.97 (23.60, 51.10) | 0.20 | 64.00 | 5.56 (.062) | .11 |
| *Structured comorbidity interview* | 4 | 180 | 30.68 (24.36, 62.15) | 0.00 | 0.00 | 1.66 (.65) | .49 |
| **Obsessive-compulsive disorder** | | | | | | | |
| *Sample size ≥ 20* | 3 | 183 | 23.33 (4.47, 66.28) | 2.39 | 93.80 | 32.27 (<.001***) | .009** |
| *Structured comorbidity interview* | 4 | 180 | 36.03 (15.37, 63.62) | 1.13 | 88.20 | 25.43 (<.001***) | .23 |
| **Substance use disorder** | | | | | | | |
| *Sample size ≥ 20* | 5 | 383 | 35.92 (21.20, 53.89) | 0.50 | 86.34 | 29.28 (<.001***) | <.001*** |
| *Structured DID interview* | 7 | 408 | 28.11 (15.54, 45.39) | 0.64 | 84.87 | 39.66 (<.001***) | <.001*** |
| *Structured comorbidity interview* | 8 | 428 | 23.87 (12.66, 40.23) | 0.75 | 85.33 | 47.73 (<.001***) | <.001*** |
| **Persistent depressive disorder** | | | | | | | |
| *Sample size ≥ 20* | 3 | 183 | 5.42 (0.44, 42.68) | 4.45 | 88.32 | 17.13 (<.001***) | <.001*** |
| *Structured comorbidity interview* | 4 | 180 | 14.43 (2.37, 53.88) | 3.17 | 88.19 | 25.41 (<.001***) | .12 |
| Note. *p* < .05*, *p* < .01**, *p* < .001***.  ***Abbreviations.*** k = number of studies; N = total sample size; CI = confidence interval; DID = dissociative identity disorder; JBI = Joanna Briggs Institute. | | | | | | | |

| **Supplementary Table 8.**  *Grading of Recommendations Assessment, Development, and Evaluation (GRADE) summary of findings table for the pooled prevalence estimates.* | | | | | | | | | |
| --- | --- | --- | --- | --- | --- | --- | --- | --- | --- |
| Psychiatric comorbidity | Number of participants (studies) | Study design | Risk of bias | Inconsistency | Indirectness | Imprecision | Publication bias | Pooled prevalence (95% CI) | Certainty of evidence (GRADE) |
| Posttraumatic stress disorder | 420 (13) | Cross-sectional designs | Not serious | Very serious^a^ | Not serious | Not serious | Not serious | 87.44 (71.79, 94.96) | ⊕⊕◯◯  Low |
| Major depressive disorder | 523 (12) | Cross-sectional designs | Not serious | Very serious^a^ | Not serious | Very serious^b^ | Not serious | 62.64 (33.45, 84.84) | ⊕◯◯◯  Very low |
| Borderline personality disorder | 593 (12) | Cross-sectional and non-randomised experimental designs | Not serious | Very serious^a^ | Not serious | Serious^c^ | Not serious | 47.05 (34.21, 60.32) | ⊕◯◯◯  Very low |
| Panic disorder | 212 (5) | Cross-sectional designs | Not serious | Very serious^a^ | Not serious | Very serious^b^ | –^e^ | 45.24 (23.42, 69.08) | ⊕◯◯◯  Very low |
| Schizoaffective disorder | 136 (3) | Cross-sectional designs | Not serious | Very serious^a^ | Not serious | Very serious^b^ | –^e^ | 44.40 (14.72, 78.70) | ⊕◯◯◯  Very low |
| Social anxiety disorder | 180 (4) | Cross-sectional designs | Not serious | Serious^d^ | Not serious | Not serious | –^e^ | 32.85 (20.41, 48.28) | ⊕⊕⊕◯ Moderate |
| Somatic symptom disorder | 577 (14) | Cross-sectional designs | Not serious | Serious^d^ | Not serious | Not serious | Serious^e^ | 31.17 (24.34, 38.96) | ⊕⊕◯◯  Low |
| Alcohol use disorder | 151 (4) | Cross-sectional designs | Not serious | Very serious^a^ | Not serious | Very serious^b^ | –^e^ | 29.15 (11.91, 55.63) | ⊕◯◯◯  Very low |
| Specific phobia | 229 (6) | Cross-sectional designs | Not serious | Not serious | Not serious | Not serious | –^e^ | 26.11 (18.09, 36.33) | ⊕⊕⊕⊕  High |
| Agoraphobia | 166 (3) | Cross-sectional designs | Not serious | Serious^d^ | Not serious | Very serious^b^ | –^e^ | 24.79 (8.20, 54.86) | ⊕◯◯◯  Very low |
| Bulimia nervosa | 180 (4) | Cross-sectional designs | Not serious | Not serious | Not serious | Not serious | –^e^ | 24.23 (18.33, 31.09) | ⊕⊕⊕⊕  High |
| Obsessive-compulsive disorder | 231 (6) | Cross-sectional designs | Not serious | Very serious^a^ | Not serious | Very serious^b^ | –^e^ | 22.10 (8.12, 47.88) | ⊕◯◯◯  Very low |
| Substance use disorder | 443 (9) | Cross-sectional designs | Not serious | Very serious^a^ | Not serious | Not serious | –^e^ | 22.10 (11.88, 37.40) | ⊕⊕◯◯  Low |
| Schizophrenia | 157 (4) | Cross-sectional designs | Not serious | Not serious | Not serious | Not serious | –^e^ | 16.11 (10.83, 23.34) | ⊕⊕⊕⊕  High |
| Unspecified schizophrenia spectrum and other psychotic disorder | 136 (3) | Cross-sectional designs | Not serious | Serious^d^ | Not serious | Very serious^b^ | –^e^ | 15.19 (1.73, 64.52) | ⊕◯◯◯  Very low |
| Functional neurological symptom disorder | 95 (3) | Cross-sectional designs | Not serious | Serious^d^ | Not serious | Very serious^b^ | –^e^ | 11.01 (1.41, 51.43) | ⊕◯◯◯  Very low |
| Anorexia nervosa | 180 (4) | Cross-sectional designs | Not serious | Serious^d^ | Not serious | Not serious | –^e^ | 10.16 (3.14, 28.46) | ⊕⊕⊕◯ Moderate |
| Bipolar disorder | 180 (4) | Cross-sectional designs | Not serious | Not serious | Not serious | Not serious | –^e^ | 10.07 (3.75, 24.36) | ⊕⊕⊕⊕  High |
| Persistent depressive disorder | 229 (6) | Cross-sectional designs | Not serious | Very serious^a^ | Not serious | Serious^c^ | –^e^ | 10.07 (2.28, 34.89) | ⊕◯◯◯  Very low |
| Generalised anxiety disorder | 185 (4) | Cross-sectional designs | Not serious | Serious^d^ | Not serious | Not serious | –^e^ | 8.71 (3.32, 20.10) | ⊕⊕⊕◯ Moderate |
| Unspecified personality disorder | 152 (3) | Cross-sectional designs | Not serious | Not serious | Not serious | Not serious | –^e^ | 8.71 (5.10, 14.37) | ⊕⊕⊕⊕  High |
| Illness anxiety disorder | 166 (3) | Cross-sectional designs | Not serious | Not serious | Not serious | Not serious | –^e^ | 2.53 (0.95, 6.55) | ⊕⊕⊕⊕  High |
| Brief psychotic disorder | 141 (3) | Cross-sectional designs | Not serious | Not serious | Not serious | Not serious | –^e^ | 1.52 (0.31, 7.25) | ⊕⊕⊕⊕  High |
| *Abbreviations.* CI = confidence interval.  ^a^Very serious inconsistency. Substantial between-study heterogeneity (I² >50%) and limited overlap between individual study confidence intervals.  ^b^Very serious imprecision. Pooled 95% confidence interval encompassed substantially different interpretations of the prevalence estimate.  ^c^Serious imprecision. Pooled 95% confidence interval was sufficiently wide to introduce meaningful uncertainty regarding the magnitude of prevalence.  ^d^Serious inconsistency. Substantial between-study heterogeneity observed (I² >50%), but substantial overlap between individual study confidence intervals.  ^e^Publication bias not assessed due to limited statistical power (*k* < 10).  ^f^Serious publication bias. Statistically significant funnel plot asymmetry indicated potential small-study effects. | | | | | | | | | |
